# External Validation of a Brain Imaging Model for Predicting Psychosis in Clinical High-Risk Cohorts

**DOI:** 10.64898/2026.10.17.26365769

**Authors:** Clara S. Vetter, Alessa Grund, Lisa Hahn, Sergio Mena, Charlotte Fraza, Julia Fietz, Nikos F. Diederichs, Dominic B. Dwyer, Paris Lalousis, Nora Penzel, David Popovic, Maria Fernanda Urquijo, Frauke Schultze-Lutter, Scott Woods, Tyrone D. Cannon, Shalaila S. Haas, Lana Kambeitz-Ilankovic, Joseph Kambeitz, Stephan Ruhrmann, Theresa Lichtenstein, Katharine Chisholm, Georg Romer, Linda A. Antonucci, Anita Riecher-Rössler, Christos Pantelis, Jarmo Hietala, Raimo K. R. Salokangas, Rebekka Lencer, Udo Dannlowski, Alexandra Korda, Alessandro Bertolino, Stefan Borgwardt, Paolo Brambilla, Stephen J. Wood, Eva Meisenzahl, Rachel Upthegrove, Peter Falkai, Karsten Heekeren, Wulf Rössler, Anastasia Theodoridou, Daniel Rueckert, Nikolaos Koutsouleris, the PRONIA Consortium

## Abstract

**Importance:** Structural magnetic resonance imaging (MRI) models for predicting psychosis in people at clinical high risk (CHR-P) are rarely tested in new cohorts, so whether and why they lose accuracy is unknown.

**Objective:** To test whether a structural MRI model predicts psychosis in external cohorts and adds to symptom ratings, whether its accuracy varies with participant characteristics, and what explains differences between cohorts.

**Design:** This prognostic study applied the PRONIA structural MRI model, re-estimated in its training sample (329 participants; 26 transitions), without refitting to 4 external cohorts. Data were analyzed from July 2025 to October 2026.

**Setting:** Early-recognition services in North America and Europe.

**Participants:** People at CHR-P in 4 external cohorts.

**Exposure:** Baseline MRI risk score.

**Main Outcomes and Measures:** Transition to psychosis by study-specific criteria (symptom duration, 1 week to 1 month); discrimination (area under the curve [AUC]), calibration, net benefit, added value beyond symptoms, and discrimination by participant characteristics, site, and outcome group.

**Results:** Among 685 external participants (mean [SD] age, 20.0 [5.1] years; 319 [46.6%] female; 106 transitions), the AUC was 0.53 in NAPLS-3, 0.56 in MUC-FRUE, 0.65 in ZInEP, and 0.69 in FePsy (pooled, 0.56; 95% CI, 0.51-0.62), compared with 0.74 in PRONIA. The calibration slope was 0.17 (95% CI, 0.01-0.34), no probabilities gave clear net benefit, and adding the MRI score to symptom ratings changed AUC by 0.01 (95% CI, −0.02 to 0.05). Only negative symptoms hinted at better discrimination, and not robustly (*q* = .15). Participants without transition scored like those in PRONIA, whereas those with transition scored lower, so sensitivity fell from 0.92 to 0.57 with similar specificity (0.52 vs 0.55). Within-site comparisons, reweighting for age, sex, and symptoms, and alternative classifiers did not restore discrimination.

**Conclusions and Relevance:** Outside its training cohort, this MRI model predicted psychosis poorly, a loss confined to people who developed psychosis and unexplained by scanners or participant characteristics. People may reach psychosis by routes that differ between cohorts, only some of them visible in brain structure. Identifying those for whom an MRI risk score works, if any, requires far more transitions, counted by shared criteria.

**Key Points:** *Question:* Does a structural brain imaging model predicting psychosis keep its accuracy in other clinical high-risk cohorts, and if not, why?

*Findings:* In this prognostic study of 685 participants across 4 external cohorts, discrimination ranged from chance to moderate (area under the curve, 0.53-0.69 vs 0.74 in the training cohort), and the score added little to symptom ratings. Participants without transition scored as in the training cohort despite different scanners, whereas those with transition scored lower.

*Meaning:* Brain imaging may predict psychosis in only some people at risk, so it could help only if symptoms or other tests first identify these people.

## Introduction

About 3% of people develop a psychotic disorder during their lifetime, most often in early adulthood.^1,2^ Outcomes are worse the longer psychosis goes untreated,^3^ so predicting who will develop it could change care. Transition to psychosis occurs within 3 years in about 1 in 4 people at clinical high risk for psychosis (CHR-P), with wide variation between cohorts,^4^ so identifying those at highest risk would let clinicians focus monitoring and care.^5^

Neuroimaging-based models for individualized prognosis in psychiatry have mostly been evaluated only within their training cohort, and whether their predicted risks match observed risks is rarely assessed.^6–10^ With a rare outcome, accuracy estimated in the training cohort is imprecise even with cross-validation, so performance elsewhere is easily overstated.^6,11,12^ A model can also lose accuracy in a study that recruits differently, uses other scanners, or defines the outcome differently, even if its brain signal carries real prognostic information.^13^ Lower accuracy in a new cohort can therefore mean that the training estimate was too high or that the cohorts differ.

Studies apply different criteria to recruit people at CHR-P and to define transition,^14–16^ so cohorts differ in who is classified as at risk and in who counts as having transitioned. Even within a cohort, people reach psychosis by different routes, to which early neurodevelopmental risk and later exposures such as cannabis use contribute to varying degrees.^17^ Structural brain alterations may precede onset in some people at risk but not in others.^18,19^ A brain-based predictor would then identify only people who develop psychosis by such a route, and clinicians would need to recognize in advance who is on it. Candidates are cognitive basic symptoms, which predict transition^14^ and have been linked to gray matter volume^20^ and a neurodevelopmental brain signature,^18^ and negative symptoms, which predict transition^21^ and have been linked to gray matter changes^22^ and a larger brain age gap.^23^ Because recruitment differs, a brain-based predictor trained in one cohort may capture mainly the routes common in that cohort.

The Personalized Prognostic Tools for Early Psychosis Management (PRONIA) consortium^24^ developed a neuroimaging-based model to predict transition to psychosis in people at CHR-P or with recent-onset depression, as part of a multimodal workflow.^25^ PRONIA recruited people at CHR-P by ultra-high-risk criteria or by cognitive basic symptoms (COGDIS).^25^ Only 26 of its 334 patients developed psychosis.^25^ Its structural magnetic resonance imaging (MRI) model, which derives a risk score from a single baseline scan, reached an area under the curve (AUC) of 0.71 in each of 2 external validation cohorts.^25^ The workflow, including the MRI model, is being tested in the randomized Computer-Assisted Risk Evaluation (CARE) trial (ClinicalTrials.gov NCT05813080).^26^

Here we applied the MRI model, developed in our group, unchanged to 4 external CHR-P cohorts to test whether it predicts transition and whether its predicted risks match observed risks. Because symptoms themselves predict transition, we tested whether the model improves prediction beyond symptom ratings. We also explored whether the model predicts transition better in people with more severe cognitive basic symptoms or negative symptoms, because such symptoms could tell clinicians in advance in whom an MRI scan is informative. Finally, we tested whether differences in participants and scanners explain differences in accuracy between cohorts.

## Methods

### Study Design and Participants

This prognostic study followed the TRIPOD+AI reporting guideline^27^ (eTable 1), and eTable 2 assesses risk of bias. We pooled individual participant data from the PRONIA training cohort^25^ and 4 external early-recognition cohorts, the North American Prodrome Longitudinal Study (NAPLS-3),^28^ the Zurich Program for Sustainable Development of Mental Health Services (ZInEP),^29^ the Basel Early Detection of Psychosis Project (FePsy),^30^ and the Munich Early Recognition of Psychosis Database (MUC-FRUE)^31^ (eTable 3). PRONIA contributed 164 participants with CHR-P (23 transitions) and 165 with recent-onset depression (3 transitions). The external cohorts contributed 685 participants (106 transitions), NAPLS-3 458 (51), ZInEP 157 (23), FePsy 37 (16), and MUC-FRUE 33 (16). The external studies enrolled only people at CHR-P, each by its own criteria (eMethods 1). All participants had a baseline T1-weighted scan and a known transition status (eFigure 1). Data were analyzed from July 2025 to October 2026.

**Figure 1.**
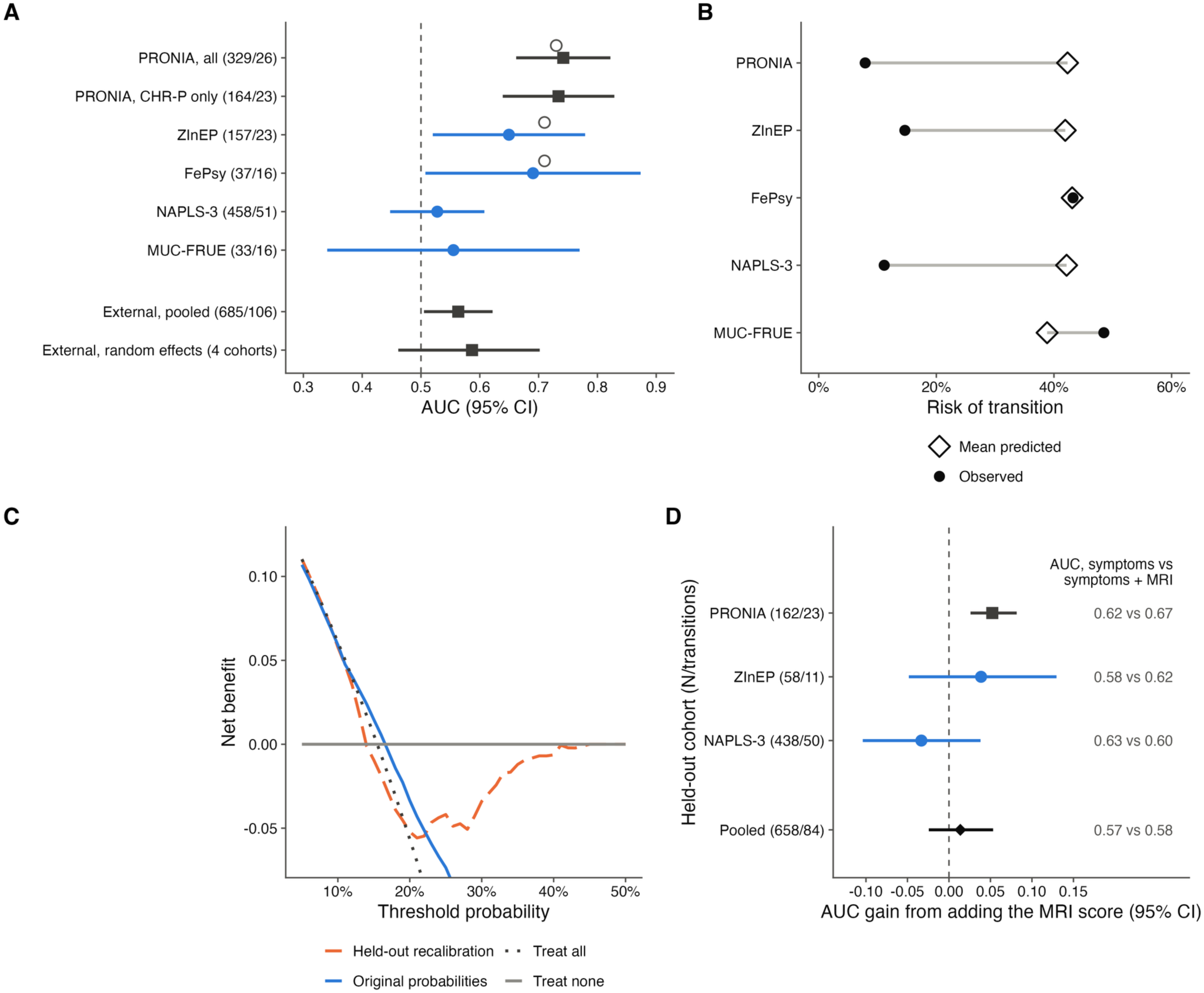
External Discrimination, Calibration, Net Benefit, and Added Value of the MRI Risk Score. (A) Area under the curve (AUC) with 95% CI by cohort (participants/transitions in parentheses). Blue circles indicate external cohorts, and squares indicate the training cohort and external summaries; these colors are used in all figures. Open circles show the AUC in the original report.^25^ (B) Mean predicted probability (open diamonds) and observed transition proportion (filled circles) by cohort. (C) Decision curves in the 4 external cohorts (685 participants) for the original probabilities and for probabilities recalibrated in the other external cohorts, with treat-all and treat-none strategies; the y-axis is truncated at −0.07. (D) Held-out AUC gain from adding the MRI score to a logistic regression of symptoms, age, and sex trained in the other 2 cohorts with symptom ratings (95% CIs from 2000 paired bootstrap replicates resampling within cohort and outcome). The diamond shows the pooled estimate. Values on the right are held-out AUCs without and with the MRI score.

Each study was approved by its ethics committee or institutional review board (eMethods 1). All participants gave written informed consent, with parental consent and participant assent for minors. Each study recorded sex as male or female. Race and ethnicity were not available for all cohorts and were not analyzed.

### Outcome and Symptom Ratings

The outcome was transition to psychosis as defined by each study, with criteria that differed in instrument, required symptom duration (1 week to 1 month), and diagnostic confirmation (eTable 3). Follow-up lasted 24 months in NAPLS-3, up to 36 months in PRONIA and ZInEP, and longer in FePsy and MUC-FRUE. Participants without a recorded transition were analyzed as nontransitioning. Baseline positive and negative symptoms were rated with the Structured Interview for Prodromal Syndromes (SIPS),^32^ cognitive disturbances (COGDIS) with the Schizophrenia Proneness Instrument,^33,34^ and functioning with the Global Assessment of Functioning (GAF; PRONIA and ZInEP)^35^ or the Social and Occupational Functioning Assessment Scale (SOFAS; NAPLS-3).^36^

### MRI Model

We re-estimated the published PRONIA structural MRI pipeline^25^ in the original training sample after reprocessing the scans with an updated version of CAT12.^37^ The pipeline combined smoothing, age adjustment, a mask of voxels reliable across scanners, principal component analysis, and a class-weighted linear support vector machine, trained with nested leave-one-site-out cross-validation so that each PRONIA participant was scored by models trained without their site (eMethods 2; eTables 4 and 5). The MRI risk score was the support vector machine decision value (higher values indicating higher risk), for external participants the median across all cross-validation models, without refitting or harmonization. Platt scaling with equal class weights, fitted to the PRONIA out-of-training scores, converted scores to probabilities. Nonlinear classifiers and classifiers without principal component analysis were retrained for comparison (eMethods 3).

### Statistical Analysis

**External validation.** Discrimination was measured by the AUC with DeLong 95% CIs and tested against chance by permuting outcomes within each cohort. We report the AUC per cohort, pooled across external participants, and as a random-effects summary (eMethods 4).

**Calibration and clinical utility.** Because Platt scaling weighted both outcome groups equally, we assessed the calibration slope rather than calibration-in-the-large (eFigure 2). We compared the Brier score with that of a constant prediction and computed net benefit^38^ for thresholds from 5% to 50%. Sensitivity, specificity, and balanced accuracy (BACC) were computed at the original threshold and, post hoc, at the BACC-maximizing threshold in each cohort (eMethods 4).

**Figure 2.**
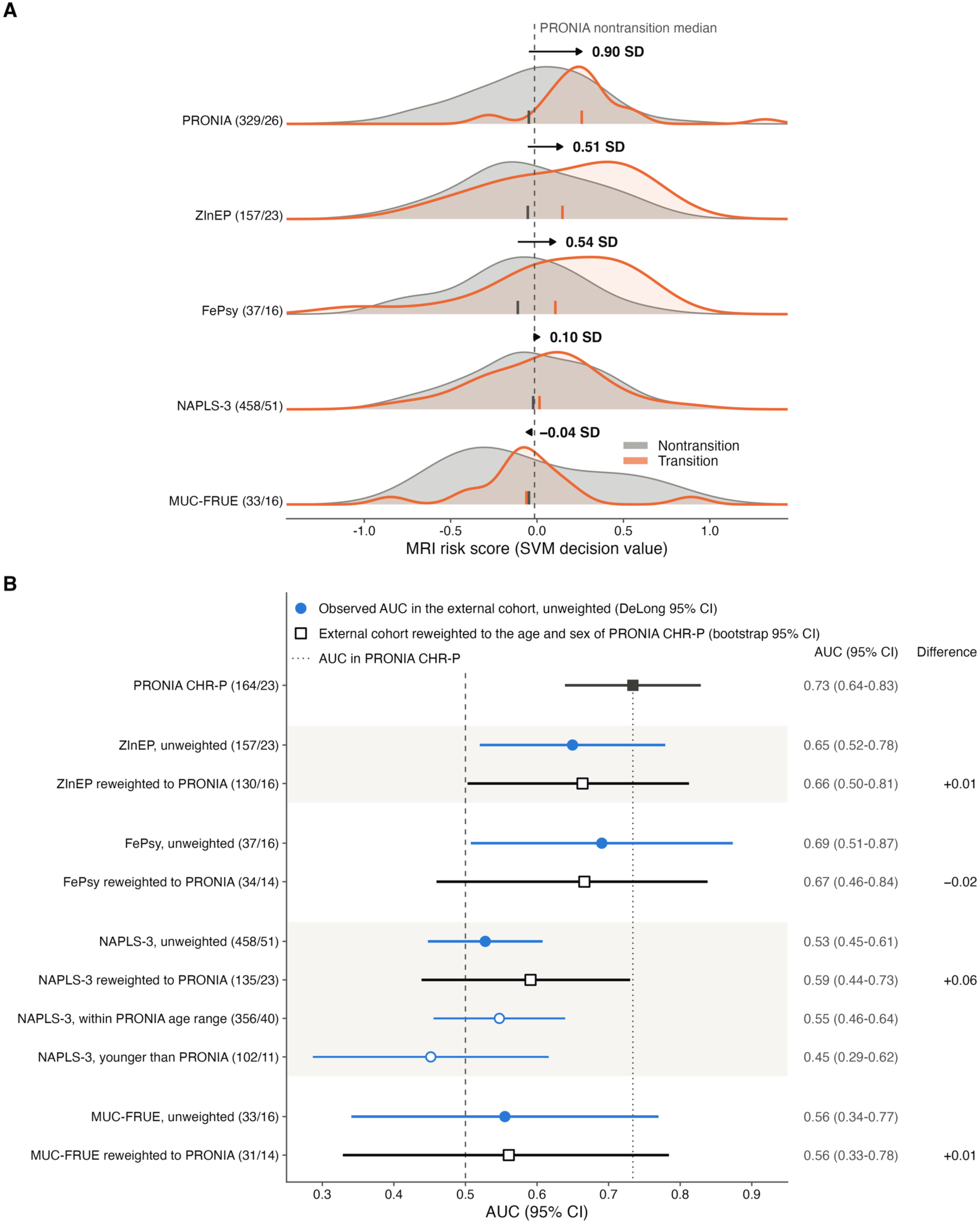
Pattern of the Loss of Discrimination and Reweighting for Participant Characteristics. (A) Density of the MRI risk score (support vector machine decision value) among participants without (gray) and with (orange) transition in each cohort (participants/transitions in parentheses); higher scores indicate higher predicted risk. Each curve is scaled to the same peak height; ticks mark group means, arrows and labels give the separation in SDs (standardized mean difference), and the dashed line marks the PRONIA nontransition median. FePsy and MUC-FRUE contributed 16 transitions each, so their curves are imprecise. (B) For each external cohort, the observed AUC (filled circles; DeLong 95% CI) and, in the row below, the AUC of the same cohort reweighted to the age and sex of PRONIA participants with CHR-P (open squares; bootstrap 95% CI; effective participants/transitions in parentheses). Open circles show NAPLS-3 participants within and below the PRONIA CHR-P age range, and the dotted line marks the PRONIA CHR-P AUC. Values on the right are AUCs and the reweighted minus unweighted difference. PRONIA reweighted to each external cohort is shown in eTable 19.

**Added value beyond symptoms.** In the 3 cohorts with symptom ratings, we trained logistic regressions of COGDIS, SIPS positive and negative scores, age, and sex, with and without the MRI score, in 2 cohorts and tested them in the third. Held-out AUC and Brier score with and without the MRI score were compared with paired bootstrap CIs and DeLong tests (eMethods 5).

**Exploratory moderation.** We estimated interactions of the MRI score with COGDIS and SIPS negative and positive scores in a logistic regression with cohort fixed effects and in a generalized additive model (eMethods 6 and 7; eTables 6 and 7). We also estimated local AUCs in overlapping windows along age, brain age gap, symptoms, and functioning and tested their range against permutations and simulations (eMethods 8 and 9; eTables 8-10).

**Differences between cohorts.** We examined whether age, sex, symptoms, or scanners explain the differences in discrimination between cohorts. We reweighted each external cohort to resemble PRONIA participants with CHR-P in age and sex, separately by outcome, and conversely reweighted PRONIA to each external cohort.^39–41^ In NAPLS-3 and ZInEP, the weights also matched symptom ratings (eMethods 10). Participants younger than any PRONIA participant with CHR-P (15.2 years), whom weighting cannot represent, are reported separately in NAPLS-3. Because the pipeline is linear, a scanner offset shifts all scores at a site without changing their ranking within it. We therefore compared nontransition scores with PRONIA and estimated a within-site AUC across the 9 NAPLS-3 sites (eMethods 11).

**Decomposition of discrimination.** Post hoc, we examined which outcome group drove the differences in discrimination. We ranked each external participant against PRONIA participants of the other outcome group. If a cohort ranks participants as PRONIA does, the mean of these placement values equals the PRONIA AUC on both sides (eMethods 12). We also pooled the external separations (standardized mean differences between outcome groups) by random-effects meta-analysis and regressed the placement values of participants who developed psychosis on age, sex, and symptoms (eMethods 12). We also computed the separation for each of the 4 principal components of the gray matter maps that carried significant weight in the score (eMethods 13; eFigures 3 and 4; eTable 11).

**Missing data, study size, and software.** Analyses used complete observations without imputation (eTable 12), and study size was set by the available cohorts. Two-sided *P* < .05 was considered nominally significant, and Benjamini-Hochberg *q* values are reported within the families listed in eTable 13. Analyses used NeuroMiner, version 1.4, in MATLAB, version R2023b, Python, version 3.14, and R, version 4.5.2 (eMethods 2, 3, and 14; eFigure 5).

## Results

### Participants

The 685 external participants had a mean (SD) age of 20.0 (5.1) years, 319 (46.6%) were female, and 106 (15.5%) developed psychosis (Table 1). Transition proportions ranged from 11.1% in NAPLS-3 to 48.5% in MUC-FRUE, compared with 14.0% among PRONIA participants with CHR-P. NAPLS-3 participants were younger than PRONIA participants and had more severe positive symptoms.

**Table 1.**
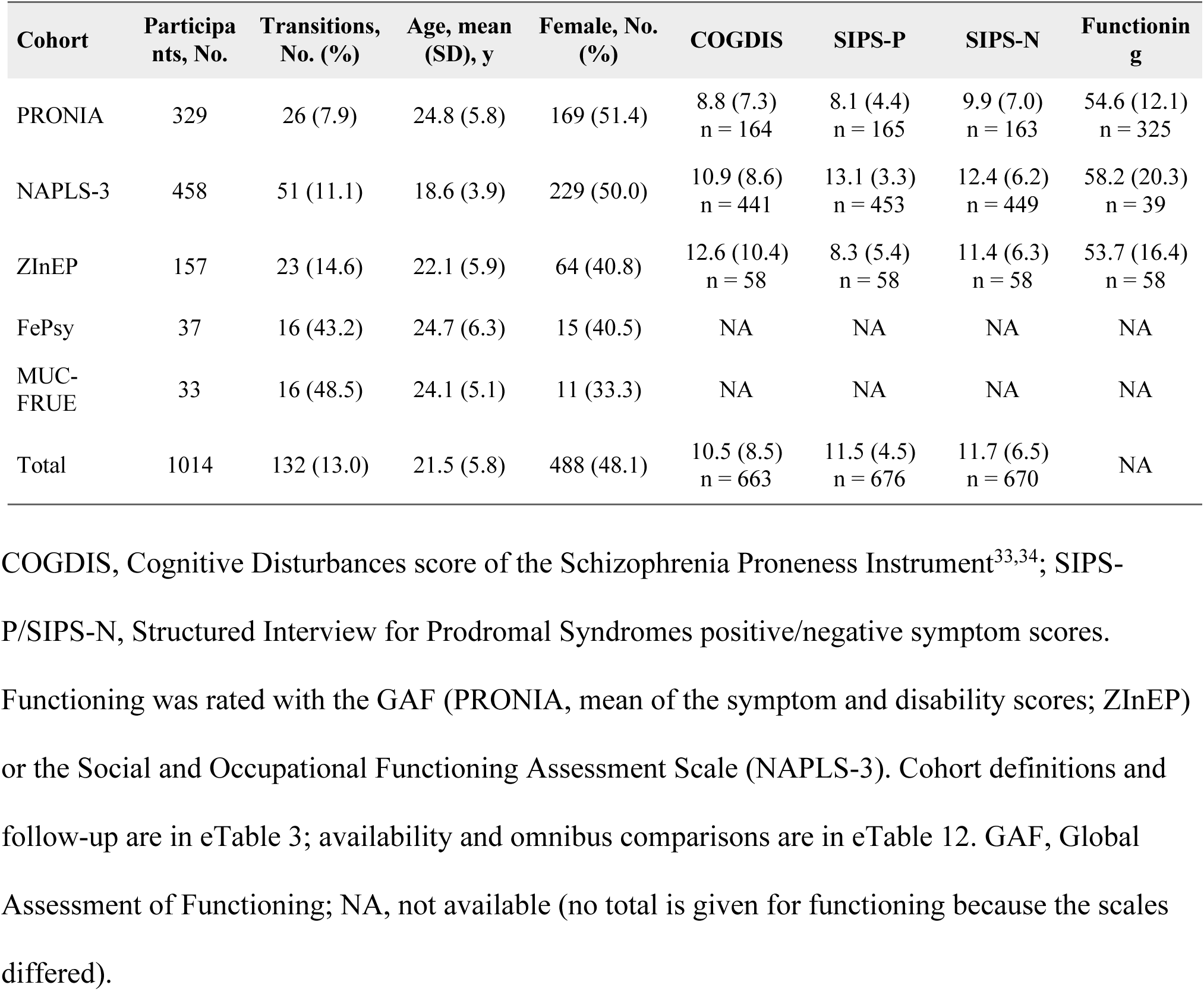
Baseline Demographic and Clinical Characteristics of the Study Cohorts. Clinical measures are mean (SD) with the number of participants with available ratings, which can be smaller than the cohort size.

| **Cohort** | **Participants, No.** | **Transitions, No. (%)** | **Age, mean (SD), y** | **Female, No. (%)** | **COGDIS** | **SIPS-P** | **SIPS-N** | **Functioning** |
|---|---|---|---|---|---|---|---|---|
| PRONIA | 329 | 26 (7.9) | 24.8 (5.8) | 169 (51.4) | 8.8 (7.3)<br>n = 164 | 8.1 (4.4)<br>n = 165 | 9.9 (7.0)<br>n = 163 | 54.6 (12.1)<br>n = 325 |
| NAPLS-3 | 458 | 51 (11.1) | 18.6 (3.9) | 229 (50.0) | 10.9 (8.6)<br>n = 441 | 13.1 (3.3)<br>n = 453 | 12.4 (6.2)<br>n = 449 | 58.2 (20.3)<br>n = 39 |
| ZInEP | 157 | 23 (14.6) | 22.1 (5.9) | 64 (40.8) | 12.6 (10.4)<br>n = 58 | 8.3 (5.4)<br>n = 58 | 11.4 (6.3)<br>n = 58 | 53.7 (16.4)<br>n = 58 |
| FePsy | 37 | 16 (43.2) | 24.7 (6.3) | 15 (40.5) | NA | NA | NA | NA |
| MUC-FRUE | 33 | 16 (48.5) | 24.1 (5.1) | 11 (33.3) | NA | NA | NA | NA |
| Total | 1014 | 132 (13.0) | 21.5 (5.8) | 488 (48.1) | 10.5 (8.5)<br>n = 663 | 11.5 (4.5)<br>n = 676 | 11.7 (6.5)<br>n = 670 | NA |

COGDIS, Cognitive Disturbances score of the Schizophrenia Proneness Instrument<sup>33,34</sup>; SIPS-P/SIPS-N, Structured Interview for Prodromal Syndromes positive/negative symptom scores. Functioning was rated with the GAF (PRONIA, mean of the symptom and disability scores; ZInEP) or the Social and Occupational Functioning Assessment Scale (NAPLS-3). Cohort definitions and follow-up are in eTable 3; availability and omnibus comparisons are in eTable 12. GAF, Global Assessment of Functioning; NA, not available (no total is given for functioning because the scales differed).

### External Discrimination

In PRONIA, the out-of-training AUC was 0.74 (95% CI, 0.66-0.82), and 0.73 (95% CI, 0.64-0.83) among participants with CHR-P (Table 2). Across the 685 external participants, the AUC was 0.56 (95% CI, 0.51-0.62; *P* = .03) (Figure 1A). Discrimination did not differ from chance in NAPLS-3 (AUC, 0.53) or MUC-FRUE (0.56) and was 0.65 in ZInEP and 0.69 in FePsy (Table 2). Alternative classifiers discriminated no better externally (AUC, 0.48-0.56; eTable 14). At the original threshold, BACC fell from 0.72 in PRONIA to 0.56 (95% CI, 0.51-0.61) externally, because sensitivity fell from 0.92 to 0.57, whereas specificity was similar (0.52 vs 0.55; Table 2; eTable 15).

**Table 2.** External Performance of the MRI Risk Score. AUC uses continuous scores and DeLong confidence intervals. Threshold metrics use the model’s original decision threshold. Brier scores assess the original probabilities. PRONIA estimates are out of training; the four external cohorts were evaluated without refitting the transition model.

| **Cohort** | **Participants/transitions** | **AUC (95% CI)** | **P value** | **BACC** | **Sens.** | **Spec.** | **Brier** |
|---|---|---|---|---|---|---|---|
| PRONIA (training) | 329/26 | 0.74 (0.66-0.82) | <.001 | 0.72 | 0.92 | 0.52 | 0.21 |
| PRONIA, CHR-P only | 164/23 | 0.73 (0.64-0.83) | <.001 | 0.72 | 0.91 | 0.52 | 0.22 |
| NAPLS-3 | 458/51 | 0.53 (0.45-0.61) | .51 | 0.56 | 0.59 | 0.53 | 0.25 |
| ZInEP | 157/23 | 0.65 (0.52-0.78) | .02 | 0.59 | 0.61 | 0.57 | 0.23 |
| FePsy | 37/16 | 0.69 (0.51-0.87) | .05 | 0.62 | 0.63 | 0.62 | 0.22 |
| MUC-FRUE | 33/16 | 0.56 (0.34-0.77) | .61 | 0.51 | 0.38 | 0.65 | 0.33 |
| External pooled | 685/106 | 0.56 (0.51-0.62) | .03 | 0.56 | 0.57 | 0.55 | 0.25 |
| Random-effects summary | 4 cohorts | 0.59 (0.46-0.70) | — | — | — | — | — |

PRONIA, CHR-P only: PRONIA participants meeting CHR-P criteria, excluding those with recent-onset depression. AUC P values use 5000 fixed-prediction label permutations, restricted within cohort for the pooled external analysis. The random-effects estimate synthesizes cohort AUCs on the logit scale (Hartung-Knapp CI; 95% prediction interval, 0.38-0.76) and is not the same estimand as the participant-pooled external AUC. Additional threshold metrics, calibration quantities, and uncertainty are in eTable 15. BACC, balanced accuracy; Sens., sensitivity; Spec., specificity.

### Calibration and Clinical Utility

As expected from the equal class weights, mean predicted risk was about 40% in every cohort, whatever the observed transition proportion (Figure 1B). The external calibration slope was 0.17 (95% CI, 0.01-0.34), compared with 0.74 in PRONIA, and the external Brier score was 0.25, compared with 0.13 for a constant prediction (eTable 15; eFigure 6). No probabilities offered a clear net benefit at thresholds from 10% to 30%, and at 20% and 30% they did worse than treating no one (Figure 1C; eTable 15).

### Added Value Beyond Symptoms

Among 658 participants with symptom ratings (84 transitions), regressions of symptoms, age, and sex reached held-out AUCs of 0.58 to 0.63. Adding the MRI score changed the pooled held-out AUC by 0.01 (95% CI, −0.02 to 0.05) and did not improve the Brier score (Figure 1D). It improved discrimination only when PRONIA was held out (difference, 0.05; 95% CI, 0.03-0.08; eTable 16).

### Exploratory Moderation

The MRI score did not predict transition better at higher COGDIS or SIPS positive scores (both *q* = .82) but did at higher SIPS negative scores (*P* = .049), without surviving correction (*q* = .15; eFigures 7 and 8; eTable 7). Within NAPLS-3, this interaction was nominally significant (*P* = .03), with an AUC of 0.58 at or above and 0.44 below the median SIPS negative score, as in ZInEP (0.72 vs 0.34) but not PRONIA (0.74 vs 0.72; eTable 6). Participants who developed psychosis had, if anything, more negative symptoms in NAPLS-3 than in PRONIA (*P* = .17; eTable 6). Local discrimination did not vary along age, brain age gap, symptoms, or functioning beyond chance (eResults; eFigures 9 and 10; eTables 17 and 18).

### Differences Between Cohorts

Reweighting to the age and sex of PRONIA participants with CHR-P changed the external AUCs little, from 0.53 to 0.59 (95% CI, 0.44-0.73) in NAPLS-3 and by at most 0.02 in the other cohorts (Figure 2B; eTable 19). Adding symptom ratings to the weights in NAPLS-3 and ZInEP gave the same AUCs with wider CIs. In NAPLS-3, the AUC was 0.45 (95% CI, 0.29-0.62) in the 102 participants younger than any PRONIA participant with CHR-P and 0.55 in the others (Figure 2B).

Scores of nontransition participants were distributed similarly in every cohort (standardized mean difference vs PRONIA, −0.17 to 0.06; eTable 20). Within NAPLS-3, score offsets among the 9 sites were small (≤0.15 SD; *P* = .99), and the within-site AUC was 0.54 (95% CI, 0.46-0.61).

### Where Discrimination Was Lost

In post hoc analyses, the separation between outcome groups ranged from −0.04 SD in MUC-FRUE and 0.10 SD in NAPLS-3 to 0.54 SD in FePsy, compared with 0.90 SD in PRONIA (Figure 2A; eFigure 11). Combined, the external separation was 0.64 SD smaller than in PRONIA (95% CI, 0.16-1.12; *P* = .009). External participants without transition ranked as in PRONIA (mean placement value, 0.71-0.81 vs 0.74), whereas those who developed psychosis ranked lower (0.45-0.65), clearly so in NAPLS-3 (difference, −0.20; 95% CI, −0.31 to −0.10) and MUC-FRUE (−0.29; eFigure 12). In NAPLS-3, all 4 components lost their separation to a similar degree (eTable 6), so the loss was not confined to one brain pattern. Participants who developed psychosis ranked more as in PRONIA with more negative symptoms in NAPLS-3 and at older age, but not after correction (eTable 21).

## Discussion

In this prognostic study, the PRONIA structural MRI model predicted transition to psychosis worse in all 4 external cohorts than in its training cohort, gave no net benefit, and added little to symptom ratings. Discrimination ranged from moderate to chance, mirroring how far people who developed psychosis scored above people who did not. The scores of people who did not develop psychosis, by contrast, were similar in all 5 cohorts. Any explanation must therefore account for a difference between PRONIA and the other cohorts that was confined to people who developed psychosis.

Such a difference argues against scanner and site effects, which are well documented in multisite imaging and usually removed by harmonization,^42,43^ because a scanner offset would also have shifted the scores of people who did not develop psychosis. In NAPLS-3, discrimination neither improved within sites nor depended on image quality (eTable 20). Site effects would explain our results only if they masked the features that set apart people who develop psychosis.

Three other explanations fit a difference confined to people who developed psychosis. First, the 26 people who developed psychosis in PRONIA may have scored high partly by chance, a known risk when accuracy rests on so few transitions, even with sites held out.^11,12^

Second, the score may capture a real signal that is present in only a subgroup of people who develop psychosis. Psychosis can develop along neurodevelopmental and other routes,^17–19^ and marked deviations in brain structure occur in only a minority of people at risk.^44^ The score would then detect people on a neurodevelopmental route but miss the others, so its discrimination in a cohort would depend on how large this subgroup is among those who develop psychosis. Because studies recruit people at CHR-P by different criteria,^14–16^ the size of this subgroup can differ between cohorts. A characteristic marking this subgroup would therefore predict better discrimination in people who have it and lower discrimination in cohorts with fewer such people. Within PRONIA’s age range, age and sex did not explain the gap, because reweighting in either direction left it in place (eTable 19). Of the symptoms we examined, only negative symptoms hinted at such a route, in line with their links to brain changes and a larger brain age gap,^22,23^ but the interaction did not survive correction, perhaps for lack of power (eDiscussion). Cognitive basic symptoms, despite their link to a neurodevelopmental brain signature,^18,20^ and positive symptoms showed no such pattern. Negative symptoms are unlikely to explain the gap between cohorts, however, because people who developed psychosis had, if anything, more of them in NAPLS-3 than in PRONIA.

Third, studies differed in how they defined and followed up transition, which changes who counts as having developed psychosis. NAPLS-3, like PRONIA, used the SIPS but required psychotic symptoms for 1 month rather than 7 days, so some PRONIA transitions would not have counted in NAPLS-3. NAPLS-3 also followed participants for 24 rather than up to 36 months, and transition risk keeps rising after 2 years.^4,45,46^ Missed late transitions would have been counted among people without transition, whereas the loss of discrimination lay with people who developed psychosis.

Whichever explanation holds, this MRI score should not be used to communicate individual risk or to guide care, least of all in people younger than anyone in its training cohort, among whom it did not discriminate in NAPLS-3. Because the score added little to symptom ratings, the CARE trial^26^ should examine whether the MRI step of the multimodal workflow still reduces false-positive predictions. Holding out sites within one consortium did not anticipate accuracy in other cohorts, so it cannot replace external validation.^47^ Telling the explanations apart requires training such models in cohorts with far more than 26 transitions, such as those of AMP SCZ^48^ and the ENIGMA CHR-P working group,^49^ and a common definition of transition and minimum follow-up. If the signal is confined to some people, imaging could follow a first step that identifies them,^50^ possibly based on neurocognition^51^ or polygenic risk,^52^ which also predict transition.

### Limitations

We tested a re-estimated model, which came close to the published AUCs in PRONIA and FePsy but not in ZInEP (0.65 vs 0.71).^25^ We could not test the MRI score within the multimodal workflow, whose other modalities were not available in comparable form across cohorts. ZInEP and FePsy overlap with the original validation, although people who developed psychosis scored lower than in PRONIA there as well. FePsy and MUC-FRUE were small, and their transition proportions above 40% suggest imaging subsamples selected by outcome, which limits their use for calibration and net benefit. Symptom ratings were comparable in only 3 cohorts. Transition was analyzed without a common time horizon, and treatment during follow-up was not recorded uniformly.

## Conclusions

The structural MRI model for psychosis trained in PRONIA predicted transition worse in 4 external CHR-P cohorts, a loss confined to people who developed psychosis, and should not inform individual care. Whether the training estimate was too high, the signal holds in only some people, or differing transition criteria drove the loss can be resolved only in cohorts with far more transitions and a common definition of transition.

## Data Availability

All data produced in the present study are available upon reasonable request to the authors

## Article Information

### Additional Contributions

We thank the study participants and the members of the PRONIA Consortium (eAppendix in Supplement 1), who performed the screening, recruitment, rating, examination and follow-up of participants, implemented the examination protocols and IT infrastructure, and organized data flow and quality control between the study sites and the central database. Data and/or research tools used in the preparation of this manuscript were obtained from the National Institute of Mental Health (NIMH) Data Archive (NDA). NDA is a collaborative informatics system created by the National Institutes of Health to provide a national resource to support and accelerate research in mental health. Dataset identifier: NDA collection 2275. This manuscript reflects the views of the authors and may not reflect the opinions or views of the NIH or of the submitters of original data to NDA. Large language models (OpenAI GPT-6 Astra and Anthropic Claude Opus 5.5; October 2026) were used for language editing, formatting, manuscript review, and assistance with analysis and plotting code; the authors reviewed all output and take responsibility for the content.

### Author Contributions

Ms Vetter had full access to all of the data in the study and takes responsibility for the integrity of the data and the accuracy of the data analysis. *Concept and design:* Vetter, Koutsouleris. *Acquisition, analysis, or interpretation of data:* All authors. *Drafting of the manuscript:* Vetter, Koutsouleris. *Critical review of the manuscript for important intellectual content:* All authors. *Statistical analysis:* Vetter, Koutsouleris. *Obtained funding:* Koutsouleris, Ruhrmann, Riecher-Rössler, Pantelis, Salokangas, Borgwardt, Brambilla, Wood, Meisenzahl, Cannon, Woods, Lichtenstein. *Administrative, technical, or material support:* Vetter, Koutsouleris. *Supervision:* Koutsouleris.

### Group Information

The PRONIA Consortium members appear in the eAppendix in Supplement 1.

### Data Sharing Statement

Aggregate results are in the article and Supplement 1. Participant-level clinical and imaging data are not publicly available because of cohort-specific consent, ethics approvals, and data-use agreements. NAPLS-3 data are available through controlled access to the NIMH Data Archive (collection 2275). PRONIA data are available from the PRONIA Consortium under a data-sharing agreement, and ZInEP, FePsy, and MUC-FRUE data from the respective study centers subject to their approvals; requests can be directed to the corresponding author. The analysis and figure-generation code (https://github.com/claravetter/pronia-mri-transition-performance-landscapes) and the performance-landscape application (https://github.com/claravetter/prediction-landscape-explorer) are publicly available and will be archived with DOIs upon acceptance. The fitted model evaluated here can be requested from the corresponding author; the PRONIA model family is described previously^25^ and at https://www.proniapredictors.eu/.

