## Supplement for "External Validation of a Brain Imaging Model for Predicting Psychosis in Clinical High-Risk Cohorts"

Supplementary Online Content

### Contents

eMethods 1. Cohorts, CHR-P definitions and baseline comparisons

eMethods 2. MRI preprocessing and model

eMethods 3. Alternative classifiers

eMethods 4. Fixed-model validation, calibration, probability updating and decision curves

eMethods 5. Added value beyond symptoms

eMethods 6. Hierarchical logistic moderation regressions

eMethods 7. GAM specification and diagnostics

eMethods 8. Performance windows, surfaces, sensitivity analyses and simulation

eMethods 9. Brain-age prediction and BrainAGE estimation

eMethods 10. Reweighting for participant characteristics and follow-up

eMethods 11. Technical-shift analyses

eMethods 12. Transition and nontransition sides of discrimination

eMethods 13. Component-wise back-projection and anatomical characterization

eMethods 14. Reusable performance-landscape application

eResults

eDiscussion

eAppendix. PRONIA Consortium members

eFigure 1. Participant flow and analysis populations

eFigure 2. Decision curves and calibration by external cohort

eFigure 3. Structural components of the MRI risk score

eFigure 4. AAL3 regional summaries of the structural components

eFigure 5. Synthetic application example

eFigure 6. Original external probabilities and apparent pooled updating

eFigure 7. MRI-score association with transition at different negative-symptom levels

eFigure 8. GAM predictions, uncertainty, and observed support

eFigure 9. Exploratory discrimination across developmental, clinical, and MRI-derived contexts

eFigure 10. Exploratory local discrimination across BrainAGE and negative symptoms

eFigure 11. MRI score of each participant by cohort and outcome

eFigure 12. Transition and nontransition sides of discrimination

eTable 1. TRIPOD+AI reporting checklist

eTable 2. Narrative assessment of bias and applicability

eTable 3. Cohort criteria, outcomes, and follow-up

eTable 4. Structural MRI acquisition parameters

eTable 5. Model card for the PRONIA-derived structural MRI transition-risk score

eTable 6. Moderation regressions without functioning and follow-up analyses

eTable 7. Functioning-complete regressions and GAM sensitivity analyses

eTable 8. Pairwise associations among the seven moderators

eTable 9. MRI-score associations with the seven moderators

eTable 10. Normative brain-age training sample and clinical-cohort application diagnostics

eTable 11. Subject-level associations with structural components

eTable 12. Clinical availability, cohort composition and omnibus comparisons

eTable 13. Statistical families and uncertainty methods

eTable 14. Discrimination of alternative classifiers

eTable 15. Calibration and additional validation metrics

eTable 16. Added value of the MRI score beyond symptoms

eTable 17. Window definitions, global tests and local estimates

eTable 18. Sensitivity of performance landscapes

eTable 19. Reweighting for participant characteristics

eTable 20. Technical-shift analyses

eTable 21. Placement-value regressions

eReferences

This supplementary material has been provided by the authors to give readers additional information about their work.

### eMethods

#### eMethods 1. Cohorts, CHR-P definitions and baseline comparisons

PRONIA included participants with CHR-P or recent-onset depression. The analysis export coded the latter group as “ROP”; this was verified as recent-onset depression against the PRONIA database for 162 of 165 records. Of the 3 exceptions, all nontransitioners, 1 was recorded as CHR, 1 as recent-onset psychosis, and 1 had no group recorded; 1 participant in the CHR-P group also had no group recorded. CHR-P operationalizations across cohorts were Structured Interview for Prodromal Syndromes (SIPS)/Criteria of Psychosis-Risk Syndromes criteria (NAPLS-3; PRONIA, together with cognitive basic symptom criteria); ultra-high-risk criteria (ZInEP); Personal Assessment and Crisis Evaluation (PACE)–derived at-risk mental state criteria (MUC-FRUE, FePsy); and cognitive basic symptom criteria (PRONIA, ZInEP, MUC-FRUE).

Transition was defined in PRONIA as a SIPS positive symptom at psychotic intensity daily for at least 7 days, in NAPLS-3 by the SIPS Presence of Psychotic Symptoms criteria (psychotic intensity for at least 1 hour a day on an average of 4 days per week for 1 month, or a seriously disorganizing or dangerous symptom), in MUC-FRUE by Positive and Negative Syndrome Scale–based Yung/PACE criteria with later ICD-10 confirmation, in FePsy by Brief Psychiatric Rating Scale–based Yung/PACE criteria, and in ZInEP by ICD-10 criteria. The original report^1^ had already validated the published model in FePsy, with the same participants, and in ZInEP, with 146 participants and 16 transitions; ZInEP now contributed 157 participants with 23 transitions because follow-up was longer. Recruitment settings, follow-up schedules and treatment context are given in eTable 3. Some MUC-FRUE participants may also have been included in the 2009 classification study of this cohort,^2^ which developed a different classifier; the present model had not been evaluated in MUC-FRUE.

Written informed consent was obtained in accordance with each cohort’s approved procedures, including parental or guardian consent and participant assent where applicable. PRONIA was approved by the local research ethics committees at participating sites, with lead approval from the Ethics Committee of the Medical Faculty of LMU Munich (351/13), and was registered in the German Clinical Trials Register (DRKS00005042). ZInEP was approved by the regional ethics committee of the Canton of Zurich (KEK-ZH-Nr. E-63/2009). FePsy was approved by the Ethics Committee of Both Cantons of Basel (Ethikkommission beider Basel, EKBB). MUC-FRUE was approved by the Ethics Committee of the Medical Faculty of LMU Munich.^2^ NAPLS-3 protocols were approved by the institutional review boards at all participating sites.

Continuous variables were compared using Kruskal-Wallis tests; BH correction was applied across the seven continuous-variable omnibus comparisons. Recorded sex and transition frequencies were compared using Fisher tests with 20,000 simulated tables and a separate two-test BH family. Pairwise-complete, two-sided Pearson correlations described the seven moderators (21 pairs) and their associations with the MRI score (seven pairs); two-sided Spearman correlations were reported as a separately corrected sensitivity analysis. MRI-score/component correlations and continuous-moderator/component correlations used Spearman coefficients, with separate four-test and 24-test families (eTables 8, 9, and 11). We did not infer demographic invariance from similar sex-stratified performance estimates.

#### eMethods 2. MRI preprocessing and model

Gray-matter volume maps were derived from baseline T1-weighted scans with CAT12 r1207^3^ in SPM12^4^ and resliced to 3-mm isotropic resolution (acquisition parameters in eTable 4). The published PRONIA pipeline^1^ was re-estimated in the original training sample after reprocessing with CAT12 r1207 instead of r1155; architecture, hyperparameter search and training procedure were retained. A model from the same pipeline is used in the CARE project.^5^

Training used NeuroMiner, version 1.4 (https://github.com/neurominer-git/NeuroMiner), with repeated nested cross-validation (outer leave-one-site-out; inner 5 repetitions of 10-fold cross-validation). Within training partitions, the pipeline applied, in order, Gaussian smoothing with a 4-, 6-, or 8-mm kernel; adjustment for age by partial correlation analysis; exclusion of voxels below the 25th, 50th, or 75th percentile of an external between-scanner reliability (G-coefficient) map, estimated from 6 traveling healthy volunteers; principal component analysis retaining 15, 20, or 25 components; standardization by the median with winsorization at 4; and a class-weighted, L2-regularized, L1-loss linear support vector machine (LIBLINEAR; C, 1/16 to 16). Models were trained in the inner training folds and evaluated by balanced accuracy in the whole inner partition (training and test folds) across 135 combinations of smoothing kernel, reliability threshold, number of components, and C. As in the original report, the best 3 combinations of each inner partition were refined by greedy forward selection of principal components, which stopped when the gain in balanced accuracy leveled off, as detected by knee-point detection, or when half of the components had been selected, and the 3 combinations with the highest mean balanced accuracy were retrained on the whole inner partition and combined into the ensemble.

All learned transformations were confined to training partitions, and held-out participants were projected through the corresponding training-derived mappings. PRONIA scores were the median out-of-training score across the applicable base learners, and class labels were assigned by majority vote. External participants were scored by the median decision value of all cross-validation models. The fitted pipeline, including the reliability mask, was applied unchanged to the external cohorts, with no refitting or cohort-specific harmonization. Probability scaling is described in eMethods 4; the model card is in eTable 5.

#### eMethods 3. Alternative classifiers

To test whether the linear support vector machine (SVM) or the unsupervised principal component analysis (PCA) limited transfer, we retrained alternative classifiers in Python, version 3.14, with scikit-learn, version 1.9.1. We used this open-source library because it provides widely used nonlinear classifiers, including gradient-boosted trees, with a common interface, and because a reimplementation outside NeuroMiner tests the result independently of the original software. Inputs were read from the NeuroMiner structure of the MRI model, namely the gray matter maps within the 3-mm mask (71 276 voxels), age, the between-scanner reliability map, and the stored outer leave-one-site-out partitions with 5 repetitions of 10-fold cross-validation within each outer training set. Of the 329 analyzed PRONIA participants, 328 had image data in this structure.

Within each training partition, preprocessing followed eMethods 2, with Gaussian smoothing (4, 6, or 8 mm full width at half maximum), adjustment for age, exclusion of voxels below the 25th, 50th, or 75th percentile of the reliability map, and standardization by the median with winsorization at 4. Six pipelines then differed in dimensionality reduction and classifier. (1) A linear reference used PCA (15, 20, or 25 components), greedy forward selection of components as in eMethods 2, and a class-weighted linear SVM (C, 1/16 to 16). (2) The same components, again with forward selection, entered a class-weighted SVM with a radial basis function (RBF) kernel and (3) class-weighted gradient-boosted trees (histogram-based, 100 iterations, depth 2 or 3, learning rate 0.05 or 0.1). (4 and 5) Linear and RBF SVMs used all standardized voxels without PCA. (6) Partial least squares (PLS) components (2, 5, or 10), which unlike principal components are chosen to covary with the outcome, entered an RBF SVM. RBF kernel widths were 0.25, 1, or 4 times the inverse number of features.

As in NeuroMiner, the 3 hyperparameter combinations with the highest mean inner balanced accuracy, including the smoothing kernel and the reliability threshold, were retrained on the whole inner partition, and each outer fold combined the resulting 150 models by their median decision value. Unlike NeuroMiner, which evaluates balanced accuracy in the whole inner partition, all 6 pipelines were selected by balanced accuracy in the inner test folds, because flexible classifiers fit their training data almost perfectly and would otherwise always be chosen. Each PRONIA participant was scored by the ensemble of the outer fold that held out the participant's site, and external participants by the median over all 1050 models, without refitting. Discrimination was evaluated as in eMethods 4, with DeLong 95% CIs, the participant-level AUC across the 4 external cohorts reported in the main text, the random-effects summary, and paired DeLong differences from the original model and from the linear reference. Because the reimplementation differed from NeuroMiner in software, its internal AUC in PRONIA was lower than that of the original model, so the alternative classifiers were compared with a linear reference trained in the same implementation. Comparisons with the linear reference therefore isolate the classifier and the dimensionality reduction, whereas comparisons with the original model show whether any alternative exceeded the published pipeline.

#### eMethods 4. Fixed-model validation, calibration, probability updating and decision curves

Continuous MRI scores were evaluated with AUC and DeLong confidence intervals. The inherited ensemble class predictions were used for the primary confusion counts and threshold metrics; they were not replaced with a new probability threshold or the sign of the aggregated score. PRONIA out-of-training predictions served as a training-cohort reference, and external predictions evaluated the unchanged fitted pipeline. ZInEP and FePsy had previously been used to benchmark the model family^1^; NAPLS-3 and MUC-FRUE had not been used to evaluate the model.

SVM decision values were converted to probabilities using logistic (Platt) calibration in NeuroMiner 1.4, with leave-one-out cross-validation and inverse class-frequency weighting. The sigmoid was fitted to PRONIA leave-one-site-out, out-of-training decision values and outcomes, and its parameters were applied unchanged to the external cohorts. The weighting and the population on which calibration was fitted are part of the evaluated probability-scaling procedure.

Predicted probabilities were clipped to [10⁻⁶, 1-10⁻⁶] before logit transformation. The calibration-in-the-large intercept was estimated from a logistic regression with the probability logit as an offset. A separate logistic regression estimated an unrestricted intercept and calibration slope. eTable 15 distinguishes these assessment quantities from coefficients used to update probabilities. Original-probability Brier scores are accompanied by p(1-p), where p is the evaluation-sample transition frequency; this is a descriptive, in-sample reference, not an independently trained prediction rule.

Pooled intercept-only and intercept-and-slope updating were fitted and evaluated on the same 685 external observations. Their results are apparent performance. The pooled intervals use 2000 participant-bootstrap replicates; the update was refitted in each resample. PRONIA, individual-cohort and held-out-update calibration intervals use the separately reported 500-replicate percentile-bootstrap procedure. In the held-out-update analysis, coefficients were learned in the other external cohorts; a constant probability based only on those training cohorts provides an additional comparator. These analyses update probability scaling, not the structural MRI classifier.

Fixed-prediction AUC p-values used 5000 outcome-label permutations and the statistic |AUC-0.5|. Pooled external permutations were restricted within cohort. Thus, the null is within-cohort exchangeability of outcomes with respect to fixed scores; it is not equality of cohort AUCs, and the pooled null distribution need not be centered on 0.5. Balanced-accuracy tests used the upper tail. Add-one p-values were (b+1)/(B+1), with b the exceedance count and B the number of valid resamples. Reported permutation tests had 5000 valid statistics. Recorded-sex stratification was descriptive in the combined PRONIA and external population, not an external-only fairness validation.

**Random-effects synthesis.** Cohort AUCs were combined on the logit scale with DeLong variances (restricted maximum likelihood), with Hartung-Knapp-Sidik-Jonkman CIs and a 95% prediction interval. Never-used and previously used cohorts were pooled separately by inverse-variance weighting and compared with a *z* test on the logit scale; a meta-regression with Hartung-Knapp adjustment is reported as a sensitivity analysis (eTable 15). With 4 cohorts, these estimates are imprecise.

**Calibration by cohort.** Because the class-weighted Platt scaling fixes mean predicted risk near 0.4, calibration-in-the-large is poor by design wherever the transition proportion differs. We therefore updated the intercept within each external cohort (logistic regression with the probability logit as offset) and plotted observed proportions against the updated predictions, grouped into fifths (NAPLS-3, ZInEP) or thirds (FePsy, MUC-FRUE), with exact binomial CIs and a loess curve (eFigure 2C-F). Intercept updating does not change the calibration slope.

**Decision curves.** Net benefit was computed as true positives/*n* − false positives/*n* × *t*/(1 − *t*) at threshold probabilities *t* from 5% to 50% in steps of 1 percentage point.^6^ Curves were computed for the original probabilities and the held-out recalibrated probabilities in the external cohorts, for the leave-one-cohort-out clinical-only and clinical-plus-MRI predictions (eMethods 5), and for treat all and treat none. Differences from the better default strategy at 10%, 15%, 20% and 30% had 95% CIs from 2000 bootstrap replicates resampling within cohort and outcome (eTable 15; eFigure 2).

**Threshold metrics.** Sensitivity, specificity and balanced accuracy were also computed at the threshold of the MRI score that maximized balanced accuracy within each cohort (eTable 15). Because this threshold is chosen in hindsight with the outcomes on which it is evaluated, it is optimistic, so each maximum was compared with the maximum reached in 5000 permutations of outcome. Sensitivity and specificity have exact binomial CIs, and balanced accuracy has percentile CIs from 2000 bootstrap replicates resampling within cohort and outcome.

#### eMethods 5. Added value beyond symptoms

Clinical-only and clinical-plus-MRI second-stage regressions were compared by leaving one cohort out. Standardization and clinical-regression coefficients were learned in the training cohorts and applied to identical held-out observations. MRI scores were held fixed; this was not nested retraining or validation of the entire MRI classifier. Pooled differences in AUC and Brier score used paired 2000-replicate bootstrap intervals by resampling held-out predictions within cohort and outcome without refitting. The resulting uncertainty is conditional on those fitted predictions (eTable 16). Predictors were COGDIS, SIPS positive and negative scores, age and sex (658 participants; 84 transitions); a second variant added functioning (254 participants; 38 transitions). In NAPLS-3, functioning was rated with the Social and Occupational Functioning Assessment Scale (SOFAS) rather than the Global Assessment of Functioning (GAF) and was available for 39 participants, 37 with complete ratings (4 transitions), so the functioning variant is uninformative for this cohort. The comparison was repeated for each held-out cohort, with paired DeLong tests. In NAPLS-3, the held-out symptom regression was also compared with the MRI score alone on the same participants (paired DeLong test).

#### eMethods 6. Hierarchical logistic moderation regressions

The symptom-complete population comprised 658 participants and 84 transitions. Continuous predictors were standardized within the complete-case analysis population. Cohort was entered as a fixed effect, with NAPLS-3 as reference; recorded sex used male as reference. Let s denote standardized MRI score and c, p and n denote standardized COGDIS, SIPS-P and SIPS-N. The clinical regression was:

logit[P(transition)] = β0 + βs s + βc c + βp p + βn n + βsc sc + βsp sp + βsn sn + cohort effects.

Model 2 added age and sex. Model 2b added MRI-score × age and MRI-score × sex to Model 2. Model 3 instead added BrainAGE (eMethods 9) and MRI-score × BrainAGE to Model 2; it did not include the demographic interactions of Model 2b. The functioning-complete hierarchy additionally included functioning and its MRI-score interaction. Its population comprised 254 participants and 38 transitions. A no-functioning clinical regression was fitted on exactly those same 254 participants, using the same means and standard deviations for shared variables. This matched comparison separates a change in included observations from the addition of functioning as a covariate.

All regressions retained the main effects underlying interactions. Coefficient p-values and 95% intervals used Wald normal approximations. BH adjustment was applied only to the declared score-by-moderator interactions within each model; main effects, likelihood-ratio tests and simple slopes were unadjusted. Nested likelihood comparisons used identical participants. AIC values in eTables 6 and 7 refer to in-sample fit, not externally validated predictive improvement. Simple slopes at mean and ±1 SD SIPS-N were calculated as linear combinations of coefficients with their covariance-based standard errors.

To check whether the direction of the MRI score × SIPS-N interaction was consistent across cohorts, the interaction was also estimated within PRONIA, NAPLS-3 and ZInEP separately (logistic regression with standardized MRI score, standardized SIPS-N and their product), and discrimination was compared below vs at or above each cohort’s median SIPS-N (eTable 6).

To examine whether participants who developed psychosis differed clinically between cohorts, age, sex, SIPS positive and negative scores and COGDIS were summarized by cohort and outcome among participants with SIPS ratings, and SIPS-N of participants who developed psychosis was compared between cohorts with Welch *t* tests (eTable 6). In PRONIA participants with CHR-P, premorbid adjustment was summarized as the mean of the Premorbid Adjustment Scale^7^ items for childhood and early adolescence (range, 0-6; higher scores indicate poorer adjustment). Its interaction with the MRI score was estimated as for SIPS-N, and discrimination was compared at or below vs above the median (eTable 6).

#### eMethods 7. GAM specification and diagnostics

The GAM used the same 658 observations and analysis-set standardization as the symptom-complete logistic regression. It contained cohort effects, a smooth of MRI score, separate smooths of COGDIS, SIPS-P and SIPS-N, and three tensor-product score-by-symptom interactions. The primary fit used restricted maximum likelihood, k=10 marginal bases and additional term-selection penalties. Approximate tensor-term p-values were BH-adjusted across the three interactions within that model. The adjustment did not cover the search across linear and smooth models. A significant tensor term was not taken as a direct test that a nonlinear interaction was required instead of a linear one.

Bounded diagnostic comparisons used k=6, a cubic-regression-spline basis, or omitted the additional term-selection penalty. We recorded convergence, model rank, effective degrees of freedom, k-index diagnostics and concurvity. Neighbor-based k diagnostics were repeated with the original, reversed and shuffled row ordering to examine tie sensitivity. The diagnostic p-values used 400 resamples; a simulated value reported as zero means no exceedance in those simulations, not an exact probability of zero. Bernoulli simulations under fixed fitted probabilities (1000 replicates, without refitting) summarized event counts, Brier score and Pearson residual statistics. These were conditional diagnostic comparisons, not an independent validation of the model.

GAM displays use the NAPLS-3 reference cohort with COGDIS and SIPS-P held at their standardized means. The support criterion for eFigure 8 is at least ten pooled participants within ±0.25 SD on both the MRI-score and SIPS-N axes. It does not require ten transitions or ten participants specifically from the reference cohort. Supported contiguous segments are shown with confidence bands, without connecting them across unsupported gaps.

#### eMethods 8. Performance windows, surfaces, sensitivity analyses and simulation

Primary windows used 20% of each moderator's observed analysis-set range and advanced by 5%. Intervals were left-closed and right-open except for the final candidate interval, which included the observed maximum; an additional final start was used if needed to reach that maximum. Supported windows required at least seven transitions and seven nontransitions. Two-dimensional cells were intersections of the same moderator intervals. Figure coordinates are interval midpoints. Participant means, which need not coincide with midpoints, are reported separately in eTable 17.

The global statistic Dmax was the largest minus smallest supported AUC, requiring at least three supported units. Its primary permutation test shuffled moderator values within cohort, moving the two coordinates jointly for a surface, and recomputed the full supported landscape. BH correction was applied separately to six one-dimensional global tests and six primary surface tests. Unrestricted moderator permutations are reported as secondary comparisons. Local bootstrap intervals resampled outcome groups separately with 2000 replicates; they are pointwise, not simultaneous intervals, and overlapping windows are not independent observations. Figure annotations refer to the global tests, not local tests of above-chance performance.

Sensitivity analyses jointly varied width and step (15%/3.75%, 20%/5%, 25%/6.25%, and 30%/7.5%). The six-moderator correction was repeated within each setting. The selected SIPS-N × BrainAGE surface was tested across the four settings as a separate, limited family; this did not correct its selection from the six primary surfaces. Descriptive comparisons used minimum support of ten in each outcome group, external-only and CHR-P-only populations, never-used external cohorts, and omission of each cohort. Ranges were recalculated in the population being analyzed. Therefore support and interval locations could both change; these comparisons do not test the invariance of a fixed subgroup (eTable 18).

**Simulation.** To decide whether the window maps could support inference at our sample sizes, we simulated MRI scores with a known local AUC while keeping the observed moderator values, cohort membership and outcomes of the SIPS-N population (670 participants; 84 transitions) and the age population (1014; 132). Scores of nontransitioners were drawn from N(0, 1) and those of transitioners from N(√2 Φ^−1^[AUC(*u*)], 1), where *u* is the moderator’s position in its range. The true local AUC was constant at 0.60 (null), linear from 0.50 to 0.70, linear from 0.45 to 0.80, a step from 0.55 to 0.75 at the midpoint, or the observed trajectory (loess-smoothed and taken as the truth).

Each scenario had 1000 replicates. Dmax was tested with 199 within-cohort permutations of the moderator, which is equivalent to permuting outcome and score within cohort against fixed moderator values. The comparison test was the MRI score × moderator interaction in a logistic regression with cohort effects, fitted to the same simulated data (eTable 18).

#### eMethods 9. Brain-age prediction and BrainAGE estimation

Brain-age estimates were obtained using a normative prediction model trained on structural MRI data from 1105 healthy controls aged 12-65 years (mean [SD], 27.70 [9.3] years; 46.43% female) scanned at 22 sites (eTable 10). Input features comprised GM-RAVENS gray-matter maps (mwp1, smoothing setting s0), resliced to 3-mm isotropic resolution and corrected for gray-matter volume. Chronological age was scaled to [0,1] during training. The pipeline used NeuroMiner 1.4 with five outer cross-validation folds for evaluation and five inner folds for hyperparameter optimization.

Within each training partition, features were initially ranked using an external age-related ranking, and the highest-ranked 50% were retained. Non-informative features, including zero-variance and non-finite columns, were removed. Scanner/site effects were adjusted using ComBat, with study site as the batch variable. Features were subsequently ranked by Pearson correlation with chronological age, followed by soft feature selection, principal component analysis, and extraction of age-predictive subspaces.

Prediction used linear support vector regression implemented in LIBLINEAR, specifically an L2-regularized, L1-loss epsilon-support vector regression model in its dual formulation. Hyperparameter optimization covered 48 parameter combinations varying the soft feature-selection multiplier and the number of extracted subspace dimensions, and 11 epsilon-support vector regression settings.

The healthy-control outer-cross-validation result was R²=0.69. In the clinical cohorts, BrainAGE was the corrected MRI-predicted age minus chronological age. We assessed MAE, RMSE, mean signed error, predicted-age correlation, predictive R² relative to the cohort mean-age reference, regression of predicted age on chronological age, and correlation between signed error and age (eTable 10). Predictive R² is not the square of the age correlation and can be negative. The clinical-cohort diagnostics and the historical healthy-control cross-validation result address different populations and were kept separate.

#### eMethods 10. Reweighting for participant characteristics and follow-up

We followed the benchmark approach of Vergouwe et al^8^ and the relatedness assessment of Debray et al.^9^ For each external cohort we fitted a logistic membership regression distinguishing it (coded 1) from PRONIA participants with CHR-P (coded 0). Covariates were age and sex in all cohorts, with SIPS positive, SIPS negative and COGDIS scores added for NAPLS-3 and ZInEP, the cohorts with symptom ratings. Each covariate interacted with outcome, so that covariate distributions were matched within the transition and nontransition groups. The AUC of the membership regression’s linear predictor quantified how different the cohorts were in these characteristics.

External participants were weighted by the predicted odds of PRONIA membership. Weights were trimmed at the 1st and 99th percentiles and rescaled to a mean of 1 within each outcome group, and the effective sample sizes are reported for all participants and for transitions. The weighted external AUC (a weighted Mann-Whitney statistic) estimates the discrimination expected in the external cohort under the measured characteristics of PRONIA. Figure 2B shows this analysis, because it uses the outcomes of the external cohorts and does not depend on the PRONIA estimate. Conversely, PRONIA participants were weighted by the predicted odds of external membership from the same regression, trimmed and rescaled in the same way, so that the weighted PRONIA AUC estimates the discrimination expected in the training cohort under the measured characteristics of the external cohort^10^; it inherits any optimism of the training estimate. CIs came from 2000 bootstrap replicates resampling within cohort and outcome, refitting the membership regression in each replicate. Age overlap was summarized as the number of external participants younger than the youngest PRONIA participant with CHR-P (15.2 years), with AUC reported inside and outside the overlapping range. Because the MRI pipeline regressed age out of the image data, the reweighting tests whether discrimination depends on the age and sex composition of the sample, not whether age shifts scores, and it cannot extrapolate below the PRONIA age range, which was therefore examined separately.

To examine whether the shorter NAPLS-3 follow-up (24 months) could explain its lower discrimination, transitions in PRONIA participants with CHR-P were dated from the transition criteria recorded at the 9- and 18-month assessments (onset date minus baseline assessment). Discrimination was recomputed counting only transitions with a documented onset as transitions and all other participants as nontransitioning, and after excluding transitions without a documented onset (eTable 6).

#### eMethods 11. Technical-shift analyses

**Why only between-site comparisons are affected.** All steps from gray-matter features to the support vector machine score are linear and use training-derived parameters (smoothing, age adjustment, masking, projection, scaling and the linear classifier). A site- or cohort-specific additive offset in the features therefore shifts every score at that site by the same constant. This leaves discrimination within the site unchanged and alters only comparisons between sites and calibration. Only site-specific differences in feature scale can change rankings within a site.

**Between cohorts.** Among participants without transition, each cohort’s score distribution was compared with PRONIA by the standardized mean difference (pooled SD) and the ratio of SDs. Separation between participants with and without transition was summarized as a standardized mean difference within each cohort (Figure 2A); pooled SD was the square root of the mean of the 2 group variances. The pooled external AUC was recomputed after outcome-blind per-cohort median centering and z-scoring of the score.

**Between NAPLS-3 sites.** Site was taken from the participant identifier and confirmed against the recorded site for all 458 participants. Score offsets among participants without transition were compared across the 9 sites with a Kruskal-Wallis test. The within-site AUC was the Mann-Whitney statistic restricted to transition/nontransition pairs from the same site. AUC was also computed after per-site median centering and z-scoring. CIs came from 2000 bootstrap replicates resampling within site and outcome.

**Image quality.** In NAPLS-3, discrimination was computed within tertiles of the CAT12 image-quality rating and of global gray-matter volume, and the score × quality and score × volume interactions were tested in logistic regressions.

#### eMethods 12. Transition and nontransition sides of discrimination

To locate differences in discrimination, each participant was ranked against PRONIA participants of the other outcome group. The placement value of a participant who developed psychosis was the proportion of PRONIA nontransition participants with a lower MRI score, and that of a nontransition participant the proportion of PRONIA transition participants with a higher score, with ties counted as one half. The mean placement value of a cohort's transition participants and that of its nontransition participants each equal the PRONIA AUC if the cohort ranks participants as PRONIA does, so comparing them shows on which side discrimination differs. All 329 PRONIA participants served as the reference, as in Figure 2A, and PRONIA participants were ranked against their own other outcome group. Placement values were regressed on standardized covariates by linear regression, separately for transition and nontransition participants, in NAPLS-3 and in PRONIA participants with CHR-P (age, sex, SIPS positive and negative scores, and COGDIS) and across the external cohorts (age and sex with cohort indicators). Benjamini-Hochberg adjustment was applied within each regression, without counting cohort indicators. Intervals and P values came from 2000 bootstrap replicates resampling participants within cohort and outcome, with the PRONIA reference and the placement values recomputed in each replicate.

Separation, the standardized mean difference between transition and nontransition participants as in Figure 2A, was synthesized across the 4 external cohorts by random-effects meta-analysis (restricted maximum likelihood with Hartung-Knapp adjustment), and the PRONIA separation was compared with the external summary by a z test using the sum of both variances. To test whether external transition participants lay in a region of the structural components not represented in PRONIA, transition participants of PRONIA and of NAPLS-3 or ZInEP were distinguished by logistic regression on the 4 components (eMethods 13). The cross-validated AUC was averaged over 50 repetitions of 10-fold cross-validation, once with all components and once after projecting out the direction of the MRI score, estimated by regressing the score on the components in PRONIA (R2 = 0.64). Nontransition participants were compared in the same way as a reference for differences between cohorts in general. P values came from 1000 label permutations.

To examine whether the loss of separation outside PRONIA was shared by the structural components of the MRI score or concentrated in some of them, the separation between participants with and without transition was computed in each cohort for the MRI score and for the expression of each of the 4 components (eMethods 13), as the standardized mean difference (transition minus nontransition, pooled SD) used for the MRI score in Figure 2A. The loss of a variable was its separation in PRONIA minus its separation in an external cohort, and the relative loss of a component was its loss minus the mean loss of the 4 components. Relative losses were tested for NAPLS-3 and for an external summary (the participant-weighted mean of the 4 external cohort separations), with Benjamini-Hochberg adjustment across these 8 contrasts. CIs and two-sided *P* values came from 2000 bootstrap replicates resampling within cohort and outcome (eTable 6). Because the components were estimated in PRONIA, their separation there is expected to be optimistic; relative losses compare components with each other and are less affected by this.

#### eMethods 13. Component-wise back-projection and anatomical characterization

To characterize structural patterns contributing to the MRI risk score, we extracted the trained coefficient vector in reduced feature space and evaluated each latent component separately. Component-wise permutation testing used 5000 label permutations within each training fold. For each permutation, the model was retrained and the reduced-space coefficient vector was recomputed. For each component, the empirical p-value was calculated as (K+1)/(5000+1), where K was the number of permuted absolute coefficients exceeding the observed absolute coefficient. P-values were adjusted across components using the Benjamini-Hochberg false discovery rate procedure. Components meeting the predefined significance threshold of 0.05 were retained for visualization.

Each retained component was independently back-projected into the original structural MRI feature space using the stored reconstruction/loading matrix from dimensionality reduction. The mapping was applied to the component-specific coefficient rather than the complete reduced-space coefficient vector, producing one anatomical relevance map per component. Earlier preprocessing operations were reversed in inverse order where applicable, including restoration of features removed before dimensionality reduction and reversal of feature scaling.

Because component order and sign are not identifiable across cross-validation folds, fold-specific maps were aligned to a running reference component space using similarity-based Hungarian assignment, followed by sign correction. The reference space was expanded when previously unmatched components were encountered. For each aligned component, the procedure stored adjusted permutation p-values, similarity-to-reference statistics, and component magnitude measures for assessment across folds.

Anatomical relevance maps were summarized using Automated Anatomical Labelling atlas 3 (AAL3) parcels^11^ and regions of the Yeo 17-network parcellation.^12^ The Yeo parcellation is restricted to cortex; cerebellar and subcortical contributions were captured by the AAL3 summaries. For each region, occupancy was calculated as 100 times the component-overlap voxel count divided by the region's total voxel count. The displayed component maps retained voxels with absolute back-projected weight greater than 3 and a log-scaled sign-based stability metric greater than 1.3. Signed weights were displayed on the spm152 template in axial radiological orientation at z = -50, -35, -20, -5, 10, 25, 40, 55 and 70 mm. Positive weights are red and negative weights blue, with a shared color scale saturating at -15 and +15. The heat map uses 27 regions of the Yeo 17-network parcellation selected from the component-wise 10% occupancy summaries, in the established region order. For those regions, all four component values are taken from the complete recorded overlap data; an entry absent from a thresholded summary is not assumed to be zero. The color scale spans 0-45%. The 27 displayed region labels are not 27 separate canonical functional networks.

The four displayed components were labeled consecutively as Components 1-4, corresponding to COMP01, COMP02, COMP04, and COMP06 in the Hungarian-aligned reference space. Participant-level component expression was the dot product of each participant's smoothed gray-matter volume map and the component voxel weights.

#### eMethods 14. Reusable performance-landscape application

We developed an interactive R Shiny application, the Prediction Landscape Explorer, to facilitate reuse of the sliding-window and performance-surface framework. The application allows users to vary window width and step size, inspect local sample size and event counts, filter by grouping variables, and generate one- and two-dimensional performance maps using user-supplied data. The demonstration uses synthetic example data to illustrate the required input structure and analysis workflow. The application address is https://claravetter.shinyapps.io/prediction-landscape-explorer/; its source-code repository is https://github.com/claravetter/prediction-landscape-explorer.

eFigure 5 illustrates this application. The app illustrates local-performance mapping and permits application to user-supplied data; the bundled synthetic examples do not reproduce participant-level study results. The illustrated options include one-dimensional sliding windows and two-dimensional local-performance maps across pairs of moderators, in both cases computing AUC, balanced accuracy, sensitivity, and specificity per window. A fully synthetic example dataset (N = 1200, prevalence ≈ 19%) is bundled to demonstrate the interface without distributing patient-level records; users can additionally upload their own CSV / RDS files and map columns to outcome, score, optional probability, optional grouping variable, and one or more numeric moderators. Source code, a dependency-lock file (renv.lock) specifying dependency versions, and a README are available in the companion repository. The default settings shown in eFigure 5 follow the fixed-range configuration of the study analyses, reported in eMethods 8 and eTable 17. Fixed-subject-count windows, fixed-range windows and k-nearest-neighbor (KNN) neighborhoods are distinct procedures.

### eResults

#### Training-cohort reference

Restricted to participants with CHR-P, PRONIA discrimination was 0.734 (95% CI, 0.639-0.829; 164 participants, 23 transitions), close to the full-sample estimate of 0.742 (eTable 15). Participants with recent-onset depression contributed 3 of the 26 transitions. With the PRONIA database’s own group labels, the CHR-P-only AUC was 0.733.

#### Random-effects synthesis

The random-effects summary AUC was 0.59 (Hartung-Knapp 95% CI, 0.46-0.70; *I*^2^ = 31%), with a 95% prediction interval of 0.38 to 0.76. Pooled AUC was 0.53 (95% CI, 0.46-0.60) in the never-used and 0.66 (95% CI, 0.55-0.76) in the previously used cohorts (difference on the logit scale, 0.55; 95% CI, −0.01 to 1.11; *P* = .06); the Hartung-Knapp meta-regression gave *P* = .02 with an estimated between-cohort variance of 0 (eTable 15). The never-used estimate rests mainly on NAPLS-3 (51 of 67 transitions).

#### Technical shift

Among participants without transition, external score distributions were close to PRONIA’s (standardized mean differences, −0.17 to 0.06; SD ratios, 0.92-1.13), whereas the separation between participants with and without transition ranged from −0.04 in MUC-FRUE to 0.90 in PRONIA (eTable 20). Per-cohort centering or z-scoring changed the pooled external AUC from 0.563 to 0.573 and 0.566. In NAPLS-3, nontransition score offsets did not differ across the 9 sites (Kruskal-Wallis *P* = .99), and the within-site AUC (0.537; 95% CI, 0.461-0.609) was close to the cohort AUC (0.528; eTable 20). Discrimination did not vary significantly with image quality (AUC, 0.56 in the best-quality and 0.49 in the worst-quality tertile; interaction *P* = .59; eTable 20), and even the best-quality third was far below PRONIA.

#### Participant characteristics and follow-up

Membership regressions separated NAPLS-3 from PRONIA well (AUC, 0.81 with age and sex; 0.88 with symptoms added) and the other cohorts moderately (0.61-0.73). Reweighting NAPLS-3 to the measured characteristics of PRONIA raised its AUC from 0.53 to 0.59 (95% CI, 0.44-0.73; 22.9 effective transitions) with age and sex and to 0.59 (0.32-0.77; 12.9 effective transitions) with symptoms added, and changed the AUC of the other cohorts by −0.02 to 0.07 (0.56 to 0.67), so all reweighted external AUCs remained below the PRONIA AUC of 0.73 (eTable 19). Because these weights nearly exclude the NAPLS-3 participants younger than anyone in PRONIA, the gain in NAPLS-3 agrees with the lower discrimination outside the overlapping age range. Discrimination was lower in participants younger than any PRONIA participant with CHR-P (NAPLS-3, 0.45 with 11 transitions; ZInEP, 0.43 with 2 transitions) than inside the overlapping age range (0.55 and 0.67). Conversely, reweighted to the characteristics of each external cohort, PRONIA retained expected AUCs of 0.72 to 0.77 with age and sex, compared with observed external AUCs of 0.53 to 0.69. The age-and-sex weights for NAPLS-3 left 14 effective transitions and the symptom-adjusted weights only 5.6, so the symptom-adjusted estimate is imprecise (0.76; 95% CI, 0.53-0.91).

Of 23 transitions among PRONIA participants with CHR-P, 13 had an onset documented at the 9- or 18-month assessment (median, 7.0 months after baseline; IQR, 4.9-8.3; maximum, 20.1). The other 10 had no onset recorded at either assessment. Three of them attended the 18-month assessment and thus developed psychosis later, whereas the timing of the other 7 is unknown. Counting only the 13 documented transitions gave an AUC of 0.65 (95% CI, 0.52-0.79), and excluding the 10 others gave 0.68 (95% CI, 0.54-0.81), compared with 0.73 (95% CI, 0.64-0.83) for all transitions and 0.53 (95% CI, 0.45-0.61) in NAPLS-3. The 10 later or undocumented transitions were separated from nontransitions with an AUC of 0.81 (95% CI, 0.72-0.90) (eTable 6).

#### Transition and nontransition sides

Nontransition participants in every external cohort were ranked against PRONIA transition participants as in PRONIA (mean placement value, 0.74 in ZInEP, 0.81 in FePsy, 0.72 in NAPLS-3, and 0.71 in MUC-FRUE vs 0.74 in PRONIA; all differences within 0.07; P = .30 to .86), whereas transition participants were ranked lower against PRONIA nontransition participants (0.65, 0.64, 0.54, and 0.45; difference from PRONIA, −0.20 [95% CI, −0.31 to −0.10] in NAPLS-3 and −0.29 [−0.42 to −0.15] in MUC-FRUE) (eFigure 12). The loss of discrimination therefore lay almost entirely with the participants who developed psychosis, which agrees with the similar nontransition score distributions (eFigure 11). Separation was 0.51 SD in ZInEP, 0.54 in FePsy, 0.10 in NAPLS-3, and −0.04 in MUC-FRUE, consistent with a common external value of 0.25 SD (Hartung-Knapp 95% CI, −0.17 to 0.67; 95% prediction interval, −0.36 to 0.87; I2 = 25%; P = .29 for heterogeneity) and smaller than the 0.90 SD in PRONIA (difference, 0.64; 95% CI, 0.16-1.12; P = .009; 0.59 [0.07-1.11] for PRONIA participants with CHR-P). With PRONIA included, heterogeneity rose to I2 = 63% (P = .02). In the space of the 4 structural components, transition participants of NAPLS-3 were distinguished from those of PRONIA (cross-validated AUC, 0.84; P < .001) only along the direction of the MRI score (0.56 without it; P = .16), and nontransition participants were not distinguished (0.48 and 0.49). ZInEP showed the same pattern (transition participants, 0.73 and 0.33; nontransition participants, 0.39 and 0.38). In the placement-value regressions, transition participants with higher SIPS negative scores in NAPLS-3 (0.10 per SD; 95% CI, 0.02-0.17; P = .02; adjusted q = .08) and older transition participants across the external cohorts (0.07 per SD; 95% CI, 0.01-0.13; P = .03; q = .052) were ranked more as in PRONIA, and female nontransition participants were ranked less as in PRONIA in NAPLS-3 (q = .035), across the external cohorts (q = .046), and in PRONIA itself (q = .08), a property of the score rather than of transfer (eTable 21). No covariate on the transition side survived adjustment.

#### Alternative classifiers

The linear reference reached an AUC of 0.65 (95% CI, 0.54-0.75) in PRONIA and 0.54 (0.48-0.60) across the external cohorts, compared with 0.74 and 0.56 for the original model (eTable 14). No alternative classifier discriminated better than the original model in PRONIA or across the external cohorts (external AUC, 0.48 to 0.56; differences from the original model, −0.08 to 0.00). Relative to the linear reference, the RBF SVM on principal components gained 0.02 (95% CI, 0.01-0.04) externally and 0.02 in PRONIA, which brought it level with the original model externally, and gradient boosting and PLS changed the external AUC by less than 0.01. Without PCA, the voxel-level SVMs discriminated less well than the linear reference (external differences, −0.05 and −0.01; PRONIA AUCs, 0.58 and 0.52), and their inner balanced accuracy was 0.50 to 0.55 in every outer fold, so with 26 transitions these classifiers did not generalize from the voxel data. In NAPLS-3, every classifier discriminated close to chance (0.51 to 0.55). The only cohort-level AUC that exceeded that of the original model by more than 0.03 was that of gradient boosting in ZInEP (0.69 vs 0.65; difference, 0.04; 95% CI, −0.02 to 0.10).

#### Added value beyond symptoms

Adding the MRI score to the symptom regression improved held-out AUC in PRONIA (difference, 0.052; 95% CI, 0.026-0.082; paired DeLong *P* < .001) but not in NAPLS-3 (−0.033; 95% CI, −0.104 to 0.038) or ZInEP (0.039; 95% CI, −0.048 to 0.130); pooled across held-out cohorts, the gain was 0.014 (95% CI, −0.024 to 0.053) and the Brier score did not improve (difference, 0.0013; 95% CI, −0.0035 to 0.0060). With functioning added, the pooled gain was 0.058 (95% CI, 0.008-0.105), but PRONIA contributed 159 of the 254 participants and NAPLS-3 only 37 participants with 4 transitions (eTable 16). In the same 438 NAPLS-3 participants, the held-out symptom regression discriminated somewhat better than the MRI score alone (AUC, 0.631 vs 0.533; paired DeLong *P* = .08).

#### Decision curves and calibration by cohort

No model had a net benefit clearly above that of the better default strategy at the key thresholds (eTable 15; eFigure 2). The original probabilities were within 0.01 of the default strategies at 10% and 15% and fell below them at 20% (−0.034; 95% CI, −0.047 to −0.021) and 30% (−0.114). Held-out recalibration and the clinical regressions were at or below the default strategies at every key threshold; the clinical-plus-MRI regression was indistinguishable from them at 10% and 15% and below them at 20% and 30%. After cohort-specific intercept updating, calibration slopes were 0.08 (95% CI, −0.16 to 0.31) in NAPLS-3, −0.03 (−0.58 to 0.52) in MUC-FRUE, 0.42 (0.05 to 0.80) in ZInEP, and 0.47 (−0.11 to 1.04) in FePsy; in NAPLS-3 and MUC-FRUE, predicted risk carried almost no information about observed risk (eFigure 2C-F).

#### Calibration and recorded-sex strata

The additional calibration summaries (eTable 15) show that poor probability calibration was present in PRONIA as well as externally. Apparent updating improves fit in the same observations (eFigure 6), while held-out updating was not consistently better than the training-cohort constant comparator. The sex-stratified estimates describe the pooled PRONIA and external population; similar point estimates do not establish equality of performance or fairness.

#### Exploratory performance landscapes

We pooled PRONIA and the external cohorts for exploratory mapping of local discrimination using fixed-range sliding windows (width=20%, step=5% of each moderator range). Adjacent windows overlap and local sample sizes vary. We summarized variation as the difference between the highest and lowest local AUC (Dmax). For each participant characteristic, this range was compared with ranges obtained after shuffling its values among participants within the same cohort.^13^ Shuffling within cohorts preserves cohort composition but does not harmonize clinical measurements, follow-up, or outcome definitions.

Across the six moderators, local AUC estimates ranged from 0.451 to 0.843 (eFigure 9). Global tests were nominally significant for age (Dmax = 0.33; *P* = .045), BrainAGE (Dmax = 0.28; *P* = .03), COGDIS (Dmax = 0.34; *P* = .048), and SIPS-N (Dmax = 0.32; *P* = .04), but not SIPS-P (Dmax = 0.31; *P* = .19) or functioning (Dmax = 0.16; *P* = .60). None survived Benjamini-Hochberg correction across the six global tests (minimum *q* = .07; eTable 17).

The trajectories show where discrimination differed and where support was sparse. Across age, AUC ranged from 0.510 to 0.839, with the highest estimate in the 30-36-year window (midpoint 33 years; 77 participants, 8 transitions; 95% CI, 0.676-0.962). SIPS-N showed higher estimates at greater symptom burden, with a maximum AUC of 0.771 in a window containing 26 participants and 9 transitions. COGDIS followed a non-monotonic trajectory, and the highest supported window (AUC, 0.80) included only 40 participants and 7 transitions. BrainAGE showed a more consistent increase, reaching AUC=0.843 in a window with 27 participants and 7 transitions (95% CI, 0.671-0.979). Each of these maxima rests on few transitions and is not a validated subgroup estimate.

Because some moderators were correlated (eTable 8), we also examined all six bivariate maps among age, BrainAGE, COGDIS, and SIPS-N. Only the SIPS-N × BrainAGE surface was nominally supported (Dmax = 0.54; *P* = .03), but it did not survive correction across the six surfaces (*q* = .15; eFigure 10; eTable 17). The 670 participants with both variables included 84 transitions; 652 participants and 79 transitions were represented in the 76 supported cells. Correlations between moderators and the MRI risk score are reported in eTable 9. Sensitivity analyses showed how strongly these ranges depended on event support and cohort composition. Increasing the minimum to ten transitions and ten nontransitions reduced the age Dmax from 0.328 to 0.147 and the BrainAGE Dmax from 0.275 to 0.132. The BrainAGE range was 0.139 in the external-only population, compared with 0.275 in the pooled analysis. For the highlighted SIPS-N × BrainAGE surface, Dmax was 0.535 externally under the seven/seven rule, but 0.307 under the ten/ten rule; the corresponding pooled ten/ten range was 0.401. The external surface used 507 participants with 61 transitions who had both variables. Joint window-width/step sensitivities and cohort-omission analyses also changed the eligible windows and estimated ranges (eTable 18). Cohort-omission analyses distinguished robust from cohort-dependent patterns. Without FePsy, the age range fell from 0.328 to 0.148, and without ZInEP, the BrainAGE range fell from 0.275 to 0.146; in the never-used cohorts alone, these ranges were 0.111 and 0.068. The SIPS-N range was 0.409 in the external-only population and 0.312 in NAPLS-3 and MUC-FRUE, and it was at least 0.278 when any single cohort was omitted (eTable 18).

**Simulation.** Ranges close to those observed arose without any true variation in local AUC: under a constant AUC of 0.60, the mean range was 0.22 for SIPS-N (95th percentile, 0.34; observed, 0.32) and 0.18 for age (95th percentile, 0.30; observed, 0.33) (eTable 18). No null distribution was simulated for the other moderators. The window test detected a large true change (0.45 to 0.80) in 41.0% of SIPS-N data sets and 15.4% of age data sets, compared with 70.1% and 58.4% for the interaction test. When the observed SIPS-N trajectory was taken as the truth, the window test rejected in 28.3% and the interaction test in 23.4% of data sets. The maps are therefore descriptive at these sample sizes.

The complete landscapes allow the observed clinical and MRI-derived patterns to be examined alongside local sample support. None of the primary six-test one-dimensional or six-test surface families had *q* < .05. The nominal associations and complete estimates are retained in eTable 17. Sensitivities in eTable 18 quantify changes in both estimated AUC range and available support. The largest range alone does not identify a stable high-performing subgroup.

#### Structural components of the MRI score

To make the high-dimensional neuroanatomical pattern more interpretable, we summarized the MRI risk score in terms of its four significant model-derived principal components, using complementary spatial and signed-weight summaries. Spatial distribution was quantified as parcel occupancy in the Automated Anatomical Labelling atlas 3 (AAL3)^11^ and in regions of the Yeo 17-network parcellation,^12^ independently of weight sign. Signed voxel-weight maps, displayed as axial radiological slices, show the positive and negative weights that define each component’s spatial contrast (eFigure 3); AAL3 parcel-occupancy summaries are shown in eFigure 4. Comp 1, the posterior visuo-attentional/anterior midline axis, had positive loadings in visual and spatial-attention regions (lingual and fusiform gyri, inferior parietal lobule) contrasted against negative loadings in anterior midline regions (pregenual anterior cingulate, medial orbitofrontal cortex); Yeo overlap was dominated by precuneus (control and default), dorsal attention (frontal eye fields, precentral ventral, posterior), and visual parcels, with additional contributions from bilateral cerebellar lobule VI in AAL3. Comp 2, the posterior midline–cerebellar/motor-cingulate axis, showed positive loadings in posterior structures (lingual gyri, precuneus, cerebellar vermis) contrasted against negative loadings in the supplementary motor area and mid-cingulate cortex; Yeo overlap was dominated by precuneus control and default-mode parcels. Comp 3, the temporoparietal association/primary visual axis, had positive loadings in multisensory and social-cognition hubs (supramarginal and inferior parietal gyri) contrasted against negative loadings in primary visual regions (calcarine sulcus, cuneus); Yeo overlap spanned lateral default-mode (parietal), salience/ventral attention (temporo-occipital, parietal opercular), and dorsal-attention parcels, with cerebellar lobule VI as a major AAL3 contributor. Comp 4, the motor–executive/posterior sensory-integrative axis, had positive loadings in frontal motor and executive regions (precentral gyrus, supplementary motor area) contrasted against negative loadings in posterior sensory-integrative areas (Heschl's gyrus, supramarginal gyrus); Yeo overlap was dominated by medial prefrontal control and medial salience/ventral-attention parcels.

For each participant, component expression was calculated as the dot product of the smoothed gray-matter volume map and the component voxel weights. These scores quantify the expression of each structural pattern. The MRI risk score was positively correlated with all four component scores (ρ = 0.44 for Comp 1 and Comp 2 and 0.46 for Comp 3 and Comp 4; all *q* < .001). Participant-level associations with the six continuous moderators did not survive correction across the 24 moderator-component comparisons (eTable 11).

In PRONIA, all 4 components separated participants with and without transition (standardized mean difference, 0.56-0.91; MRI score, 0.90), and in NAPLS-3 all 4 lost most of this separation (−0.09 to 0.19; MRI score, 0.10) (eTable 6). Losses ranged from 0.61 to 0.72 SD across components in NAPLS-3 and from 0.59 to 0.71 SD in the external summary, and no component lost more or less than the others (relative loss, −0.06 to 0.08; all *q* = .93; eTable 6). In PRONIA and NAPLS-3, the components were nearly uncorrelated with each other (Spearman ρ, −0.07 to 0.22), so the loss affected spatially distinct patterns alike. In the smaller external cohorts, separation varied more between components (for example, 0.68 for Comp 3 and −0.40 for Comp 4 in FePsy), in line with their wide CIs.

#### Brain-age model in the clinical cohorts

To interpret the BrainAGE-related performance patterns, we examined how closely the normative model’s age predictions matched chronological age in the clinical cohorts (eTable 10). Mean absolute error ranged from 5.80 to 6.87 years and root-mean-square error from 7.71 to 9.64 years. Predictive R², relative to each cohort's mean-age reference, was negative in every cohort (−2.95 to −0.71), although predicted and chronological age were positively correlated (Pearson *r* = 0.29-0.49). Residual error was negatively associated with age (r=-0.30 to -0.18). Because the observed BrainAGE range spanned approximately 78 years, each primary 20% window was 15.6 years wide (eTable 17), so the windows with the highest local AUC were populated by participants with large positive BrainAGE values. Clinical-cohort prediction error and residual age dependence therefore need to be considered when interpreting the BrainAGE-related performance patterns.

#### Moderation regressions

We explored whether phenotypic variables modified the association between the MRI risk score and transition using logistic interaction regressions and generalized additive models (GAMs). Because functioning was missing for many participants, we fitted regressions without functioning (658 participants; 84 transitions) and in the functioning-complete subset (254 participants; 38 transitions). In the larger sample, the clinical interaction regression had the lowest AIC among its hierarchical extensions, although the differences were small (483.26, compared with 485.18 for demographic main effects, 486.24 for demographic interactions, and 486.77 for the BrainAGE model; eTable 6). The MRI risk score was positively associated with transition at mean symptom levels (β = 0.26; SE, 0.13; *P* = .04), as were SIPS-P (β = 0.49; *P* < .001) and SIPS-N (β = 0.28; *P* = .045).

The MRI risk score × SIPS-N coefficient was positive (β = 0.25; 95% CI, 0.001-0.50; *P* = .049) but did not survive correction across the three score-by-symptom interactions (*q* = .15; eFigure 7). Estimated MRI-score slopes were 0.011 at -1 SD SIPS-N, 0.262 at the mean, and 0.513 at +1 SD. Adding age and sex main effects did not improve fit (likelihood ratio *P* = .35), nor did adding demographic interactions (*P* = .23) or BrainAGE and its score interaction (*P* = .30; eTable 6).

In the functioning-complete population, the estimated score-by-SIPS-N interaction was attenuated both with functioning (β = 0.04; *P* = .87) and without functioning on the same participants (β = 0.16; *P* = .44; eTable 7). The two populations also differed in cohort composition, because PRONIA contributed 159 of the 254 functioning-complete participants, whereas NAPLS-3 contributed 438 of the 658 symptom-complete participants. The contrast between the larger and smaller regressions therefore cannot be attributed to adjustment for functioning alone. When fitted on all but one cohort and tested on the left-out cohort, a clinical-only logistic regression benchmark also transferred poorly (held-out AUC, 0.568; 658 participants, 84 transitions), and adding the fixed MRI score changed AUC by 0.014 (95% CI, −0.024 to 0.053; eTable 16). These comparisons were conditional on the existing MRI scores and did not re-evaluate the complete MRI training procedure.

In the GAM fitted to the same 658 participants, the MRI-score × SIPS-N interaction met the correction threshold across the three tested interactions (*P* = .02; *q* = .047). The result was sensitive to model specification (without the additional term-selection penalty, *P* = .10 and *q* = .30). These within-model corrections did not account for comparing different functional forms, so the findings are exploratory. Full sensitivity analyses and model diagnostics are reported in eTable 7 and eFigure 8.

Within cohorts, the MRI score × SIPS-N interaction was positive in all 3 cohorts with symptom ratings: NAPLS-3 (β = 0.29; SE, 0.14; *P* = .03), PRONIA (0.13; SE, 0.28; *P* = .65) and ZInEP (0.51; SE, 0.39; *P* = .19). Discrimination was higher at or above than below the cohort median SIPS-N in NAPLS-3 (0.58 vs 0.44) and ZInEP (0.72 vs 0.34) and similar in PRONIA (0.75 vs 0.72) (eTable 6).

Participants who developed psychosis had a mean (SD) SIPS-N of 15.1 (6.2) in NAPLS-3, 13.7 (6.9) in ZInEP and 11.9 (9.8) in PRONIA (NAPLS-3 vs PRONIA, difference 3.1; 95% CI, −1.4 to 7.7; *P* = .17). In PRONIA, SIPS-P separated participants with and without transition more than SIPS-N (standardized difference, 0.76 vs 0.33); in NAPLS-3, SIPS-N separated them at least as well as SIPS-P (0.49 vs 0.36), and the MRI score separated them little (0.09 vs 0.80 in PRONIA) (eTable 6). In PRONIA participants with CHR-P, premorbid adjustment did not differ between participants with and without transition (mean [SD], 1.68 [0.83] vs 1.65 [0.78]; *P* = .73) and did not modify the association between the MRI score and transition (β = 0.16; SE, 0.29; *P* = .57). The AUC was 0.70 at or below and 0.80 above the median (eTable 6).

The score-by-SIPS-N coefficient was nominally positive in the symptom-complete linear regression but did not survive its three-interaction family. In the matched functioning population, the no-functioning interaction was not nominally supported even before adding functioning. The primary GAM tensor met its within-model adjustment, but the model without additional term-selection penalties did not. Shuffling tied-score rows substantially changed several k-index diagnostics with negligible change in fitted likelihood. The combined results support exploratory characterization, not independent confirmation of nonlinear moderation (eTables 6 and 7).

### eDiscussion

Target-population heterogeneity can make model performance vary within a cohort as well as between cohorts. CHR-P populations span adolescence and early adulthood, when normative changes in brain structure overlap spatially with regions implicated in psychosis risk,^14,15^ which may make an MRI-derived risk pattern more distinguishable at some developmental stages than at others. Alongside chronological age, BrainAGE, defined as MRI-predicted age minus chronological age, may reflect additional variation in maturation,^16,17^ although it can also be affected by scanner, preprocessing, and population mismatch, and is not a direct marker of disease. Clinical presentation is similarly heterogeneous, and negative and attenuated positive symptoms, cognitive disturbances (COGDIS), and functioning vary widely among individuals meeting CHR-P criteria.^18,19^ Thus, even among individuals recruited under the same broad risk label, a structural MRI risk score may carry different amounts of prognostic information across developmental and clinical contexts.

Sliding-window maps show where local discrimination appears to change along a measured characteristic, but at the sample sizes available here they are descriptive. In design-preserving simulations, ranges close to those observed arose without true variation, and the window test detected even a large true change in fewer than half of data sets (eTable 18). The apparent variation along age and BrainAGE depended on single cohorts, and requiring more transitions per window narrowed several ranges. The maps therefore show where more data are needed, not where the model works.

The developmental and clinical hypotheses are still relevant in light of prior work on cortical maturation and distributed structural alterations in psychosis risk.^14,20-23^ Negative symptoms are clinically prognostic in CHR-P^24^ and have been associated with brain alterations in CHR-P and early psychosis.^25,26^ Cognitive basic symptoms have likewise been linked to gray-matter alterations.^18,27^ BrainAGE, however, is an MRI-derived age-deviation score, not direct evidence of atypical maturation. It can reflect regression to the mean, scanner effects, preprocessing, and mismatch with the age-model training population as well as age-related structural variation.^16,17,28^ The clinical-cohort age-prediction errors and residual age dependence reinforce the need to distinguish an MRI-derived age gap from a direct measurement of biological maturation.

The regression analyses provided limited convergence with the landscape findings. In the larger sample, the estimated MRI score-transition association strengthened with negative-symptom burden, but the linear interaction did not survive correction for multiple comparisons. Support for the negative-symptom interaction in the GAM depended on model specification, and the correction did not account for the comparison of different model forms. In the smaller subset with functioning data, the interaction was weaker even when functioning was not included in the model. Its attenuation therefore cannot be attributed to adjustment for functioning alone. Together, these results motivate further study of clinical context but do not establish a subgroup in which the score should be used. BrainAGE did not show comparable evidence of independent moderation in the hierarchical logistic regressions.

The exploratory analyses have additional limitations. They included PRONIA and were not independent external validation. Clinical measures were available in different subsets, and moderation regressions used complete cases. Windows and surface cells overlapped, and the minimum of seven transitions and seven nontransitions was chosen after inspecting estimate stability. Corrections were applied within stated families, not across the entire sequence of landscape, component, linear-model, GAM, and sensitivity analyses. Component expressions are mathematically related to the classifier. GAM diagnostics were affected by tied symptom scores and row ordering, while high concurvity and penalty sensitivity limit term-level interpretation. The maps are therefore descriptive and support neither causal interpretation nor claims of validated subgroups.

### eAppendix. PRONIA Consortium members

The authors listed here performed the screening, recruitment, rating, examination, and follow-up of the study participants. They were involved in implementing the examination protocols of the study, setting up its IT infrastructure, and organizing the flow and quality control of the data analyzed in this manuscript between the local study sites and the central study database.

**Department of Psychiatry and Psychotherapy, Ludwig-Maximilian-University, Munich, Germany:** Shalaila Haas, Claudius Hoff, Ifrah Khanyaree, Aylin Melo, Susanna Muckenhuber-Sternbauer, Yanis Köhler, Ömer Öztürk, Nora Penzel, David Popovic, Adrian Rangnick, Sebastian von Saldern, Rachele Sanfelici, Moritz Spangemacher, Ana Tupac, Maria Fernanda Urquijo, Johanna Weiske, Antonia Wosgien, Camilla Krämer

**Department of Psychiatry and Psychotherapy, University of Cologne, Cologne, Germany:** Karsten Blume, Dennis Hedderich, Dominika Julkowski, Nathalie Kaiser, Thorsten Lichtenstein, Ruth Milz, Alexandra Nikolaides, Tanja Pilgram, Mauro Seves, Martina Wassen

**Department of Psychiatry (Psychiatric University Hospital, UPK), University of Basel, Switzerland:** Christina Andreou, Laura Egloff, Fabienne Harrisberger, Ulrike Heitz, Claudia Lenz, Letizia Leanza, Amatya Mackintosh, Renata Smieskova, Erich Studerus, Anna Walter, Sonja Widmayer

**Institute of Mental Health & School of Psychology, University of Birmingham, United Kingdom:** Sian Lowri Griffiths, Mariam Iqbal, Mirabel Pelton, Pavan Mallikarjun, Alexandra Stainton

**Department of Psychiatry, University of Turku, Finland:** Alexander Denissoff, Anu Ellilä, Tiina From, Markus Heinimaa, Tuula Ilonen, Päivi Jalo, Heikki Laurikainen, Antti Luutonen, Akseli Mäkela, Janina Paju, Henri Pesonen, Reetta-Liina Säilä, Anna Toivonen, Otto Turtonen

**Department of Psychiatry (Psychiatric University Hospital LVR/HHU Düsseldorf), University of Düsseldorf, Germany:** Sonja Botterweck, Norman Kluthausen, Gerald Antoch, Julian Caspers, Hans-Jörg Wittsack

**Department of Basic Medical Science, Neuroscience and Sense Organs - University of Bari Aldo Moro:** Giuseppe Blasi, Giulio Pergola, Grazia Caforio, Leonardo Fazio, Tiziana Quarto, Barbara Gelao, Raffaella Romano, Ileana Andriola, Andrea Falsetti, Marina Barone, Roberta Passiatore, Marina Sangiuliano

**Institute for Translational Psychiatry of the University of Münster, Germany:** Marian Surmann

**Department of Child Adolescent Psychiatry of the University of Münster, Germany:** Olga Bienek

*Workgroup of Paolo Brambilla, University of Milan, Italy:*

• **Department of Neuroscience and Mental Health, Fondazione IRCCS Ca' Granda Ospedale Maggiore Policlinico, University of Milan, Milan, Italy:** Carlo Altamura, Marika Belleri, Francesca Bottinelli, Adele Ferro, Marta Re

• **Programma2000, Niguarda Hospital, Milan:** Emiliano Monzani, Maurizio Sberna

• **San Paolo Hospital, Milan:** Armando D’Agostino, Lorenzo Del Fabro

• **Villa San Benedetto Menni, Albese con Cassano (CO):** Giampaolo Perna, Maria Nobile, Alessandra Alciati

*Workgroup of Paolo Brambilla at the University of Udine, Italy:*

• **Department of Medical Area, University of Udine, Udine, Italy:** Matteo Balestrieri, Carolina Bonivento, Giuseppe Cabras, Franco Fabbro

• **IRCCS Scientific Institute “E. Medea”, Polo FVG, Udine:** Marco Garzitto, Sara Piccin

### eFigures

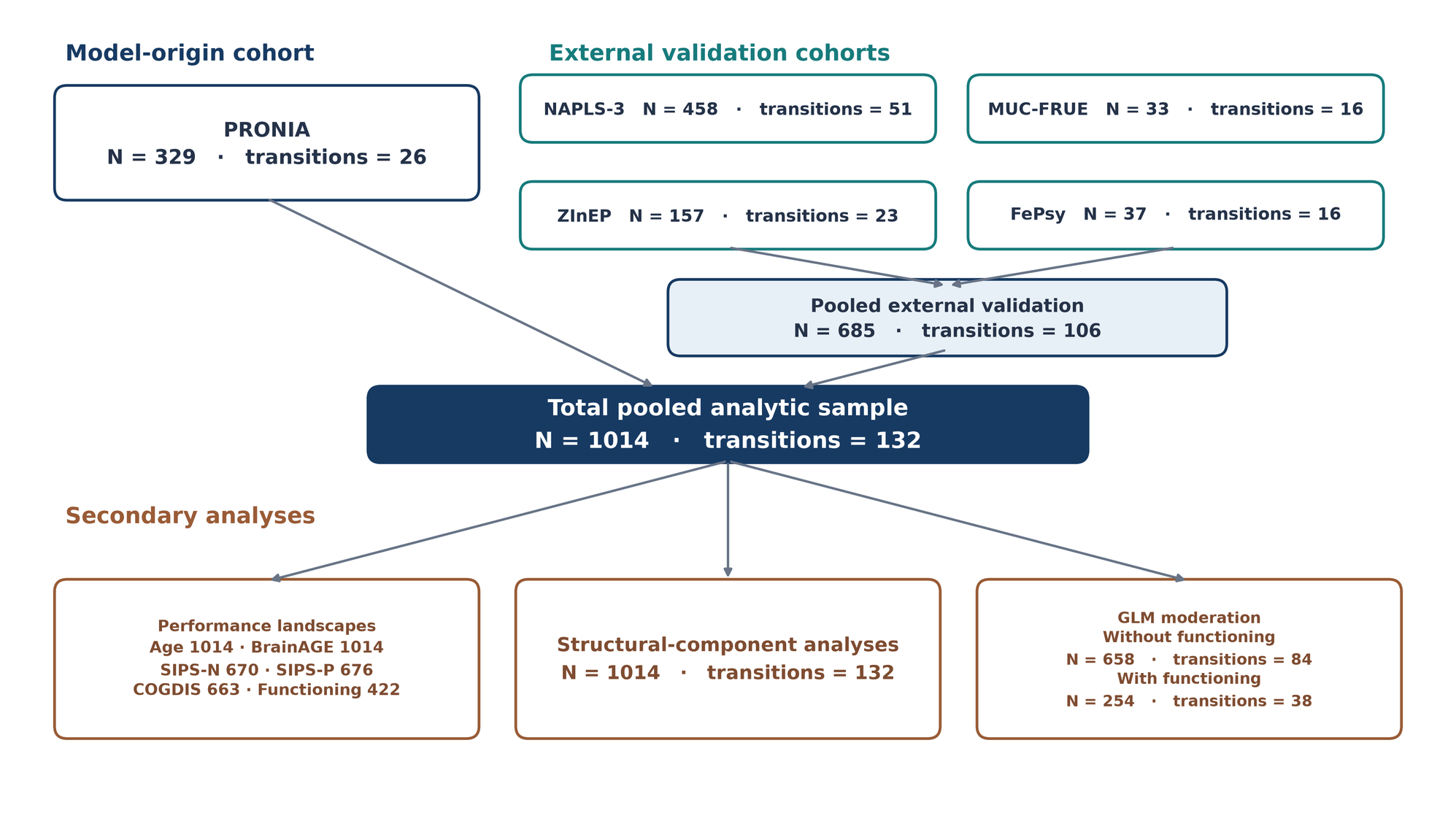

eFigure 1. Participant flow and analysis populations. PRONIA provided 329 training-cohort participants (26 transitions); four external cohorts provided 685 participants (106 transitions). The pooled sample contained 1014 participants and 132 transitions. Clinical-measure availability determined the exploratory populations (age and BrainAGE, 1014 each; SIPS-N, 670; SIPS-P, 676; COGDIS, 663; functioning, 422). Symptom-complete moderation used 658 participants with 84 transitions, and functioning-complete moderation used 254 with 38 transitions. These are available analysis populations, not the union of participants represented by supported windows or cells, which is reported separately in eTable 17. COGDIS, cognitive disturbances; SIPS-P and SIPS-N, Structured Interview for Prodromal Syndromes positive and negative symptom scores; GLM, generalized linear model; BrainAGE, corrected MRI-predicted age minus chronological age.

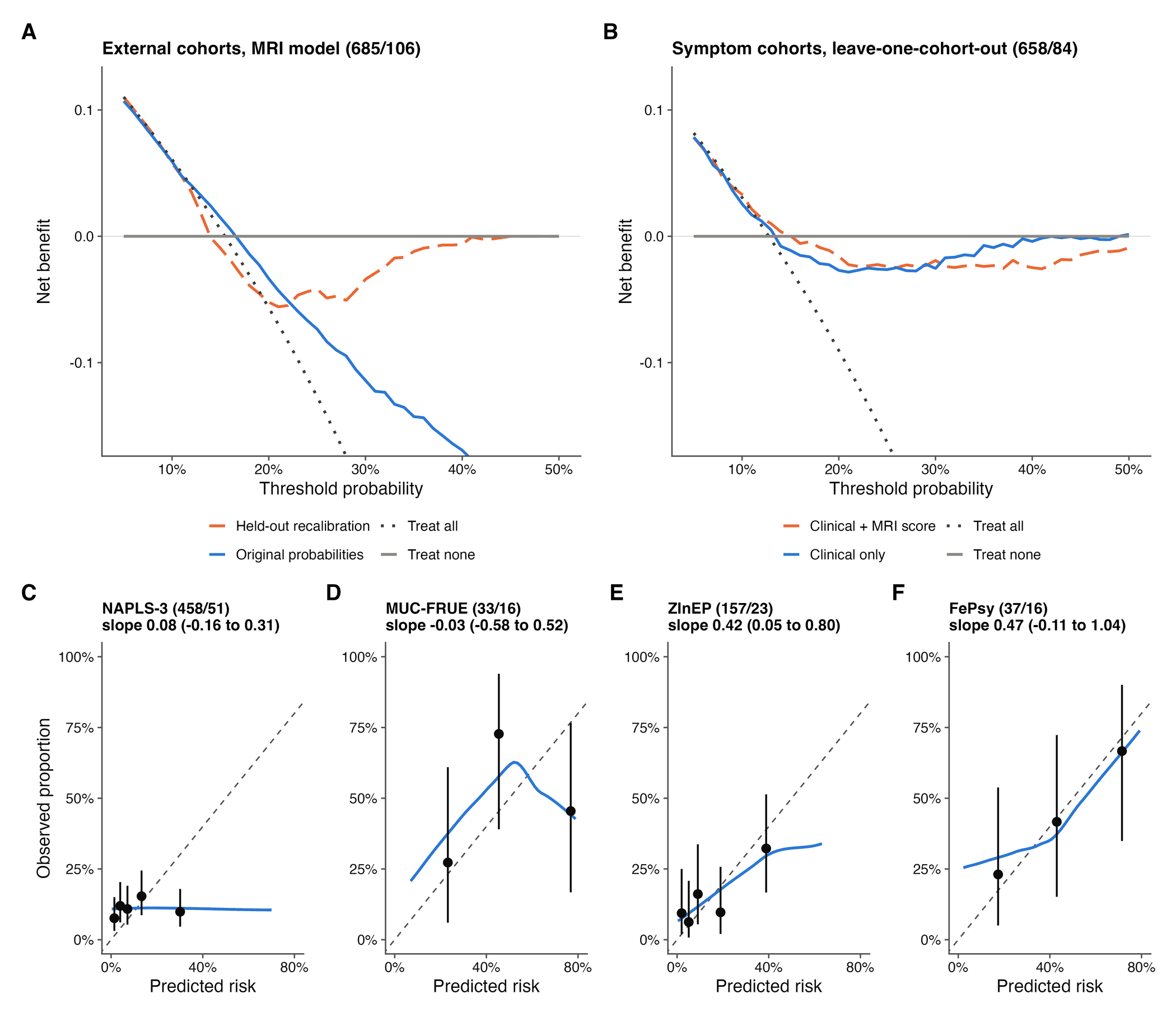

eFigure 2. Decision curves and calibration by external cohort. (A) Net benefit of the original MRI-model probabilities and of probabilities recalibrated in the other external cohorts, in the 4 external cohorts (685 participants; 106 transitions). (B) Net benefit of leave-one-cohort-out logistic regressions with symptoms, age and sex, with and without the fixed MRI score, in PRONIA, NAPLS-3 and ZInEP (658 participants; 84 transitions). Treat all and treat none are the default strategies; the y-axis is truncated at −0.16. Differences from the better default strategy at 10%, 15%, 20% and 30% are given in eTable 15. (C-F) Calibration in each external cohort after cohort-specific intercept updating (participants/transitions and calibration slope with 95% CI in each title). Points are observed proportions with exact 95% CIs in fifths (NAPLS-3, ZInEP) or thirds (MUC-FRUE, FePsy) of predicted risk; the blue line is a loess curve and the dashed line indicates perfect calibration.

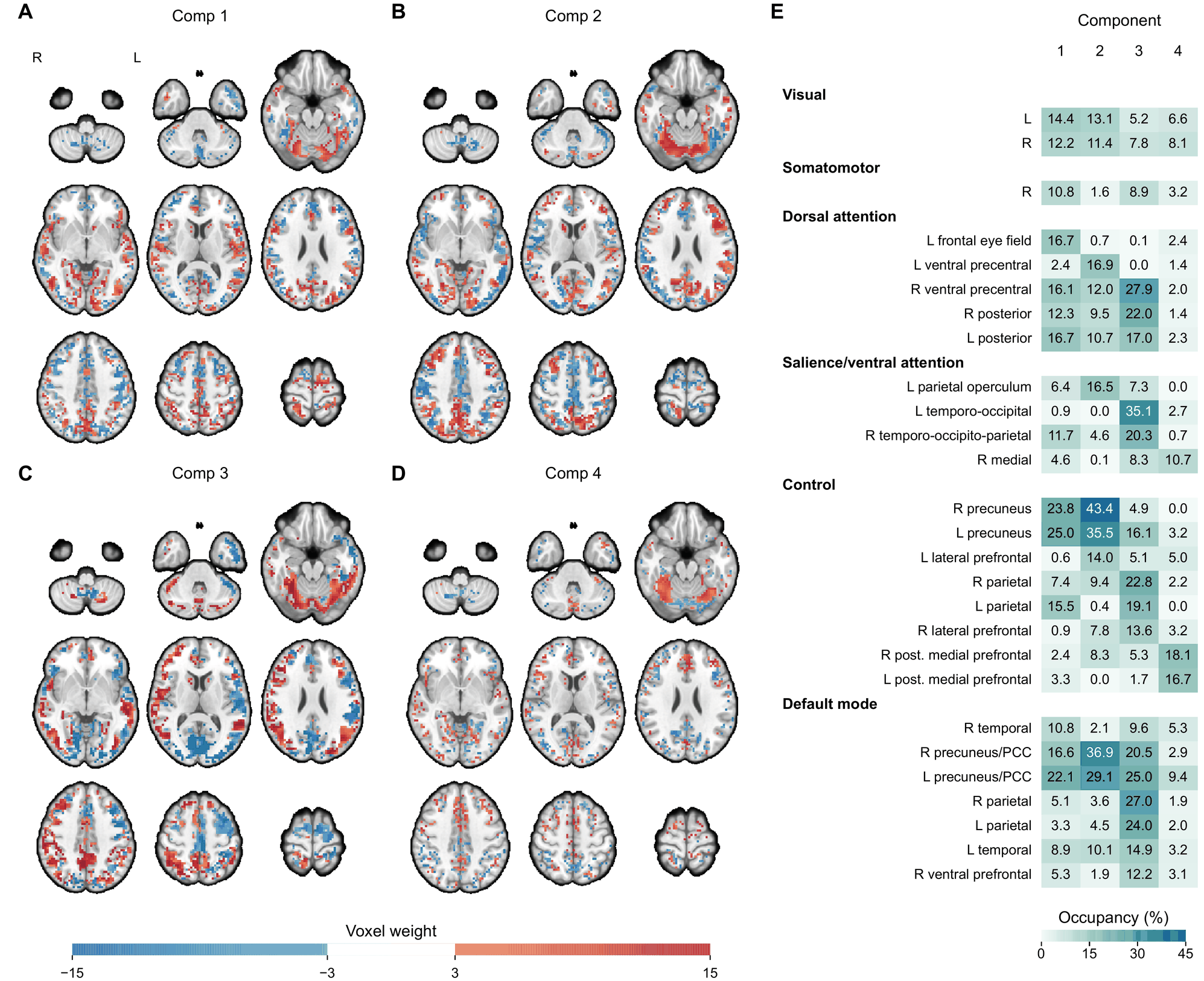

eFigure 3. Structural components of the MRI risk score. (A to D) Signed voxel-weight maps for Comp 1-4 on the spm152 template in axial radiological orientation; R and L on the first slice of A mark the right and left hemispheres. Slices are at z = -50, -35, -20, -5, 10, 25, 40, 55 and 70 mm (left to right, top to bottom). Retained voxels have absolute weight greater than 3 and a log-scaled sign-based stability metric greater than 1.3. Red and blue indicate positive and negative weights, shaded from medium at an absolute weight of 3 to full saturation at 15; the color bar is blank between -3 and 3 because sub-threshold weights are not displayed. The components are described by their dominant spatial and signed-loading patterns (Comp 1, posterior visuo-attentional/anterior midline; Comp 2, posterior midline-cerebellar/motor-cingulate; Comp 3, temporoparietal association/primary visual; Comp 4, motor-executive/posterior sensory-integrative). (E) Heatmap of occupancy in 27 selected regions of the Yeo 17-network parcellation for each component, grouped by network (L, left; R, right; PCC, posterior cingulate cortex; post., posterior). Occupancy (cell values, %) is 100 times the component-overlap voxel count divided by the network-region voxel count; the color scale spans 0-45%. AAL3 summaries are in eFigure 4.

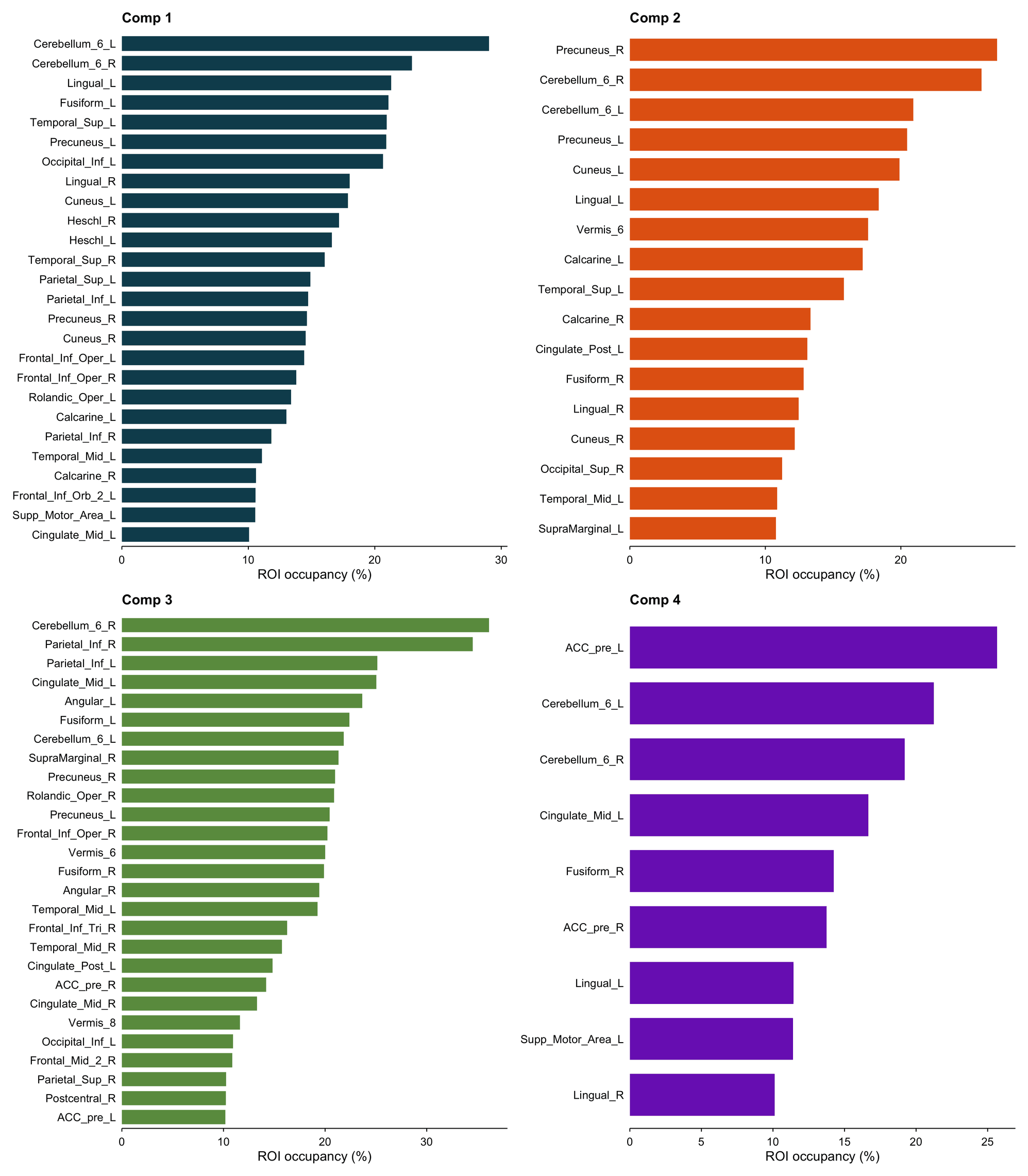

eFigure 4. AAL3 regional summaries of the structural components. Bars show parcel occupancy by suprathreshold absolute component weights and therefore summarize spatial distribution independently of sign. Comp 1: posterior visuo-attentional/anterior midline; Comp 2: posterior midline-cerebellar/motor-cingulate; Comp 3: temporoparietal association/primary visual; Comp 4: motor-executive/posterior sensory-integrative. Labels are descriptive; the signed maps are shown in eFigure 3. AAL3, Automated Anatomical Labelling atlas 3; Comp, component; ROI, region of interest. Occupancy is the percentage of a parcel's voxels overlapped by the retained component, irrespective of sign. L and R denote left and right hemispheres.

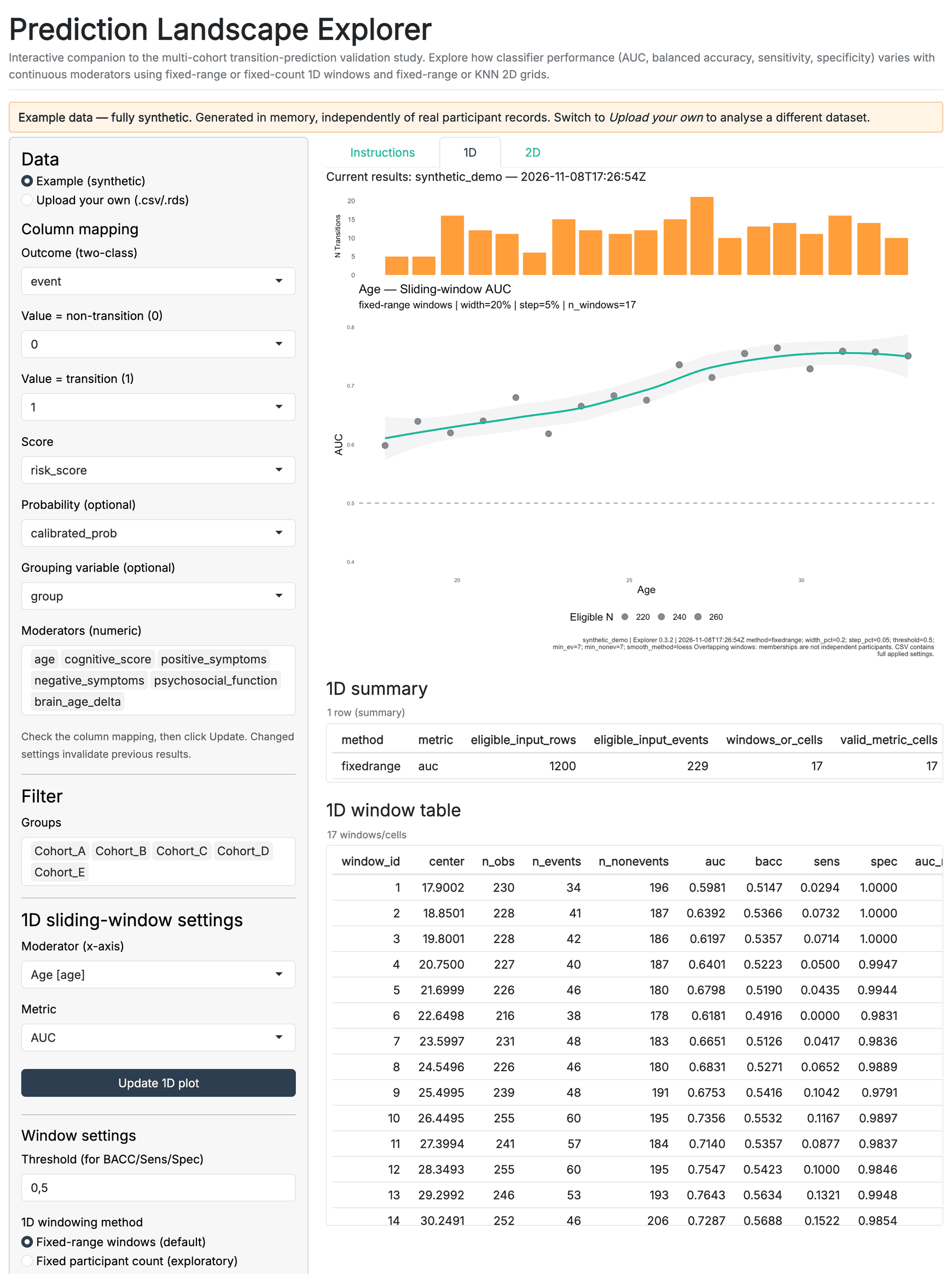

eFigure 5. Synthetic application example. AUC is shown in overlapping windows that each span 20% of the age range and advance by 5% of it, with a descriptive smoother. The example data comprise 1200 observations with approximately 19% event prevalence. This demonstrates the interface and an encoded age-related pattern, not an analysis of the clinical cohorts, an independent validation, or evidence of false-positive-rate control. These default settings follow the study's primary fixed-range definition (eTable 17), but the data are synthetic. AUC, area under the receiver operating characteristic curve.

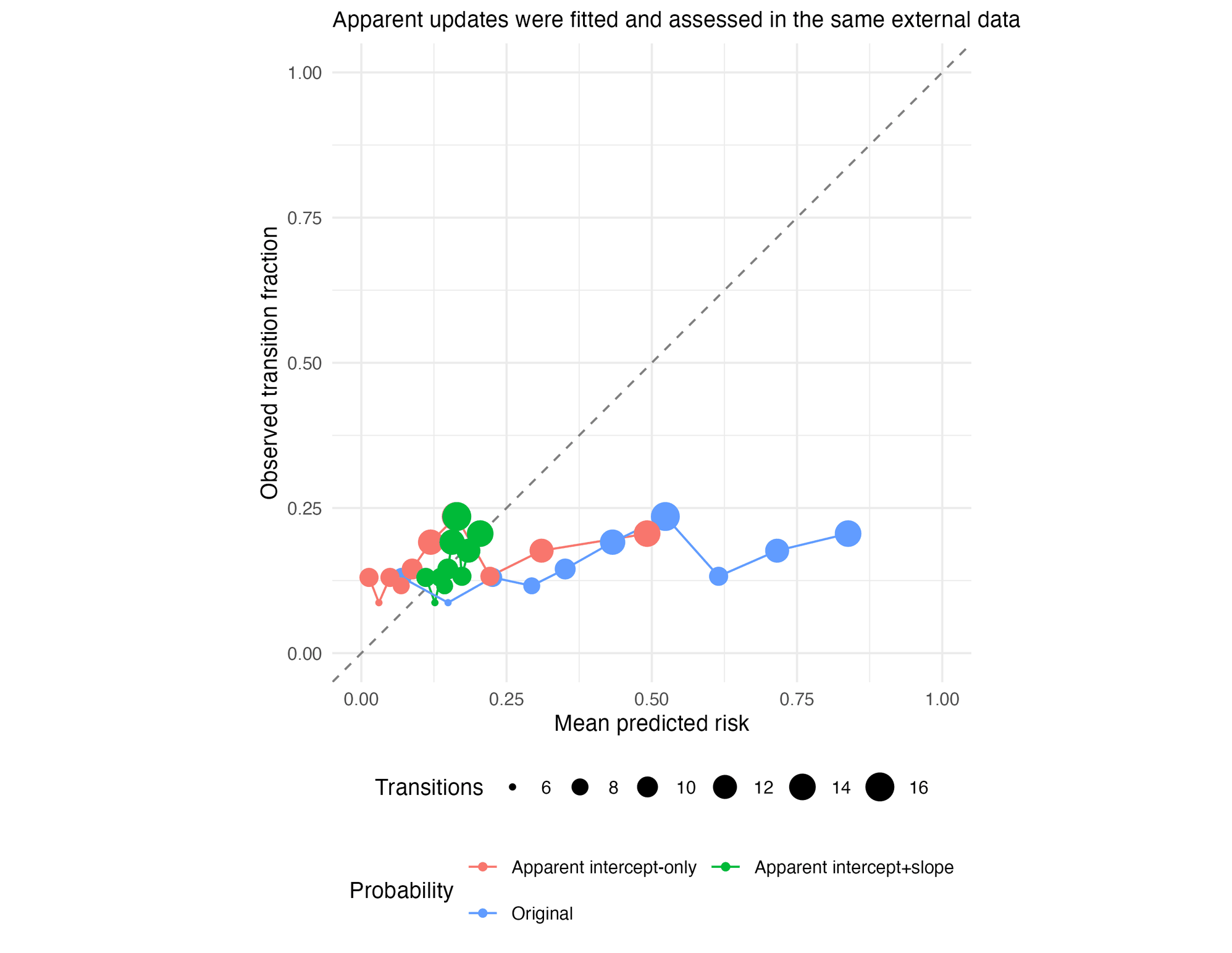

eFigure 6. Original external probabilities and apparent pooled updating. Original probabilities were assessed in 685 external participants (106 transitions). Intercept-only and intercept-and-slope updates were fitted and evaluated in those same observations. The pooled Brier scores were 0.245, 0.145 and 0.130, respectively; the descriptive pooled-prevalence reference was 0.131. Confidence intervals in eTable 15 use 2000 participant-bootstrap replicates, refitting the update in each replicate. These apparent results are distinct from held-out updating and do not establish improved external ranking or clinical utility.

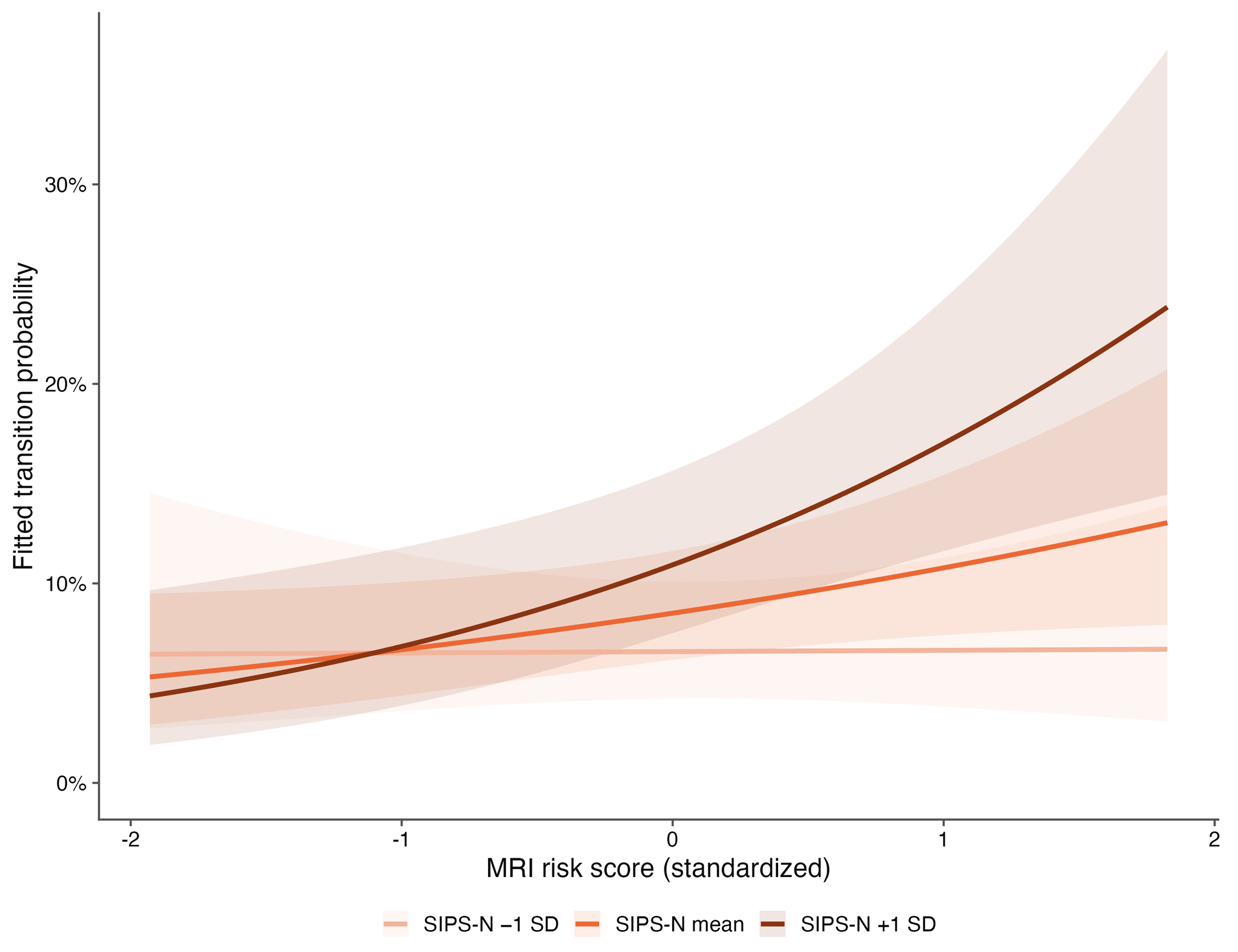

eFigure 7. MRI-score association with transition at different negative-symptom levels. Curves and 95% pointwise model-based confidence bands are from the symptom-complete logistic regression (658 participants; 84 transitions), evaluated for the NAPLS-3 reference cohort with COGDIS and SIPS-P at their analysis-set means. SIPS-N is shown at −1 SD, its mean, and +1 SD. The MRI-score × SIPS-N coefficient is 0.25 (95% CI, 0.001-0.50; *P* = .049; Benjamini-Hochberg *q* = .15 across 3 interactions). Darker lines indicate higher SIPS-N. These are fitted probabilities from an explanatory regression, not the original classifier probabilities or validated individualized predictions.

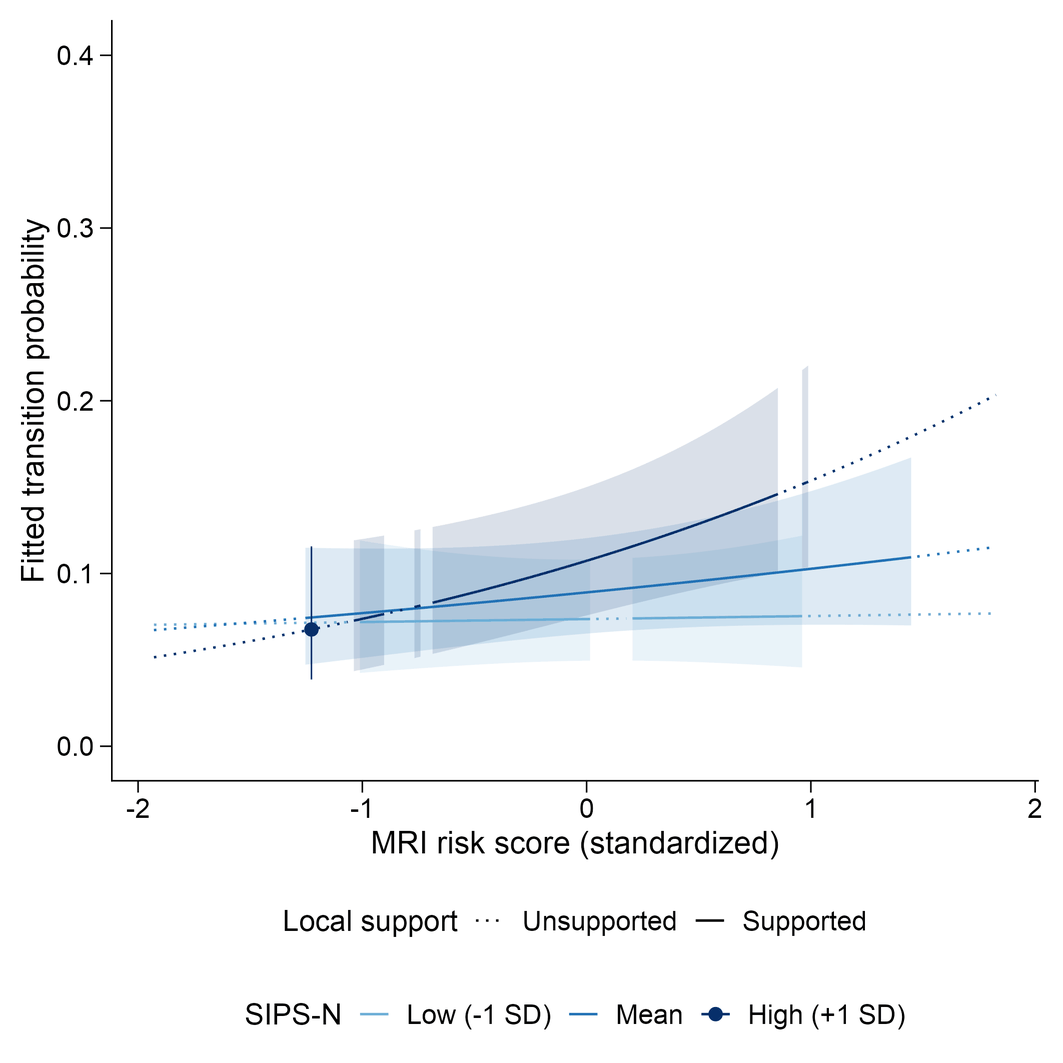

eFigure 8. GAM predictions, uncertainty, and observed support. Slices show modeled probability at SIPS-N values of -1 SD, the mean and +1 SD for the NAPLS-3 reference cohort with COGDIS and SIPS-P held at their analysis-set means. Solid lines and confidence bands are restricted to contiguous supported segments; unsupported segments are distinguished rather than joined by a ribbon. Support requires at least ten pooled participants within ±0.25 SD on both MRI score and SIPS-N, not ten transitions and not reference-cohort-specific support. Bands are pointwise model-based 95% intervals. The primary tensor *P* = .02 and within-GAM *q* = .047 do not cover selection across model forms; the penalty-sensitivity results are in eTable 7. GAM, generalized additive model; COGDIS, cognitive disturbances; SIPS-P and SIPS-N, Structured Interview for Prodromal Syndromes positive and negative symptom scores; SD, standard deviation.

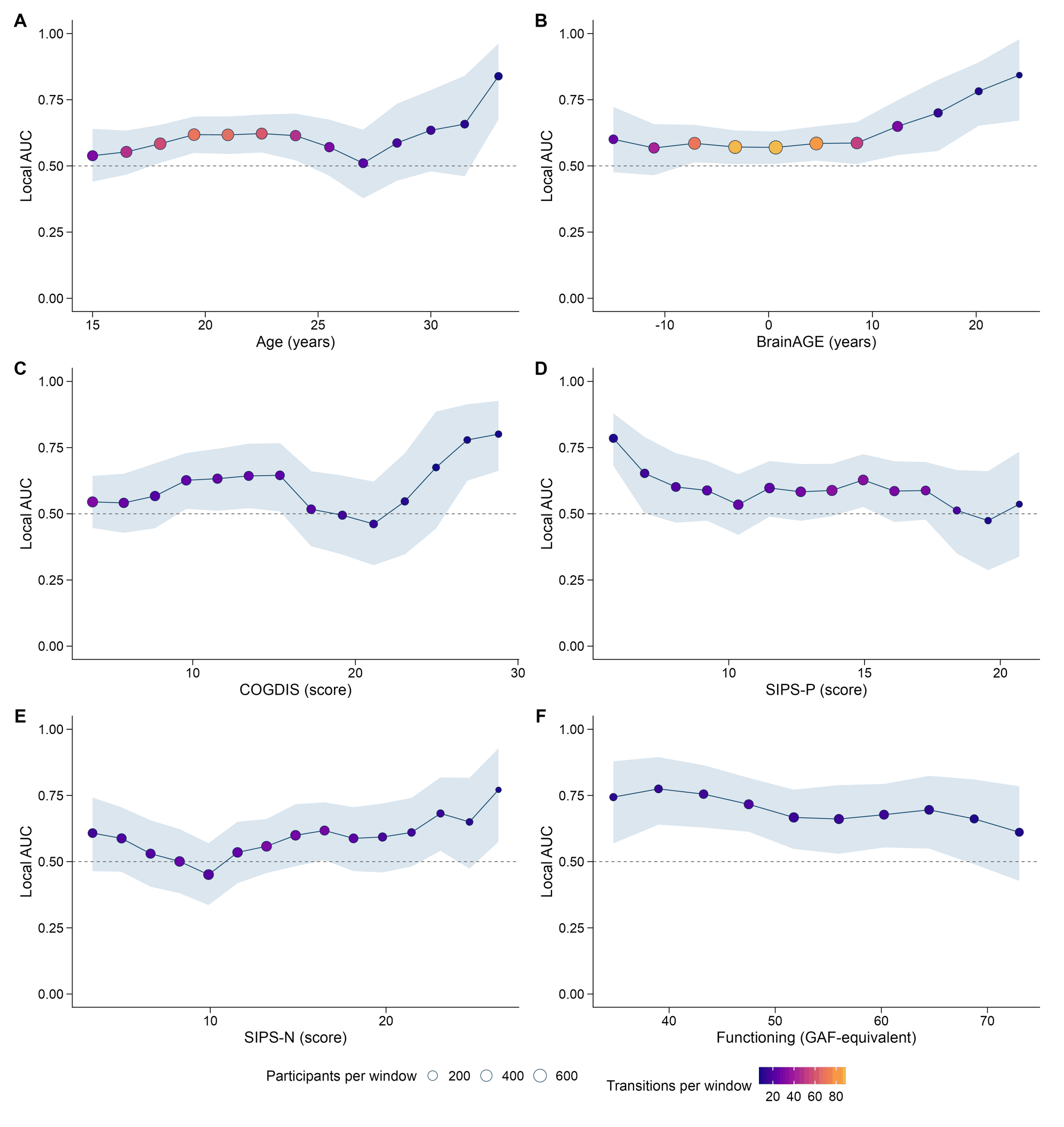

eFigure 9. Exploratory discrimination across developmental, clinical, and MRI-derived contexts. Windows span 20% of the observed moderator range, advancing by 5%, in the pooled PRONIA and external sample. Points show AUC at each window midpoint; ribbons are 95% percentile intervals from 2000 outcome-stratified bootstrap replicates. Each window contains at least seven transitions and seven nontransitions. Point size indicates the number of participants in a window and color indicates the number of transitions. Panel p-values are global within-cohort moderator-permutation tests of Dmax, not individual-window tests; q-values correct across the six global tests. Adjacent windows overlap. Age and BrainAGE are in years, symptom axes in score points, and functioning in GAF-equivalent points. None of the global tests survived the six-test correction.

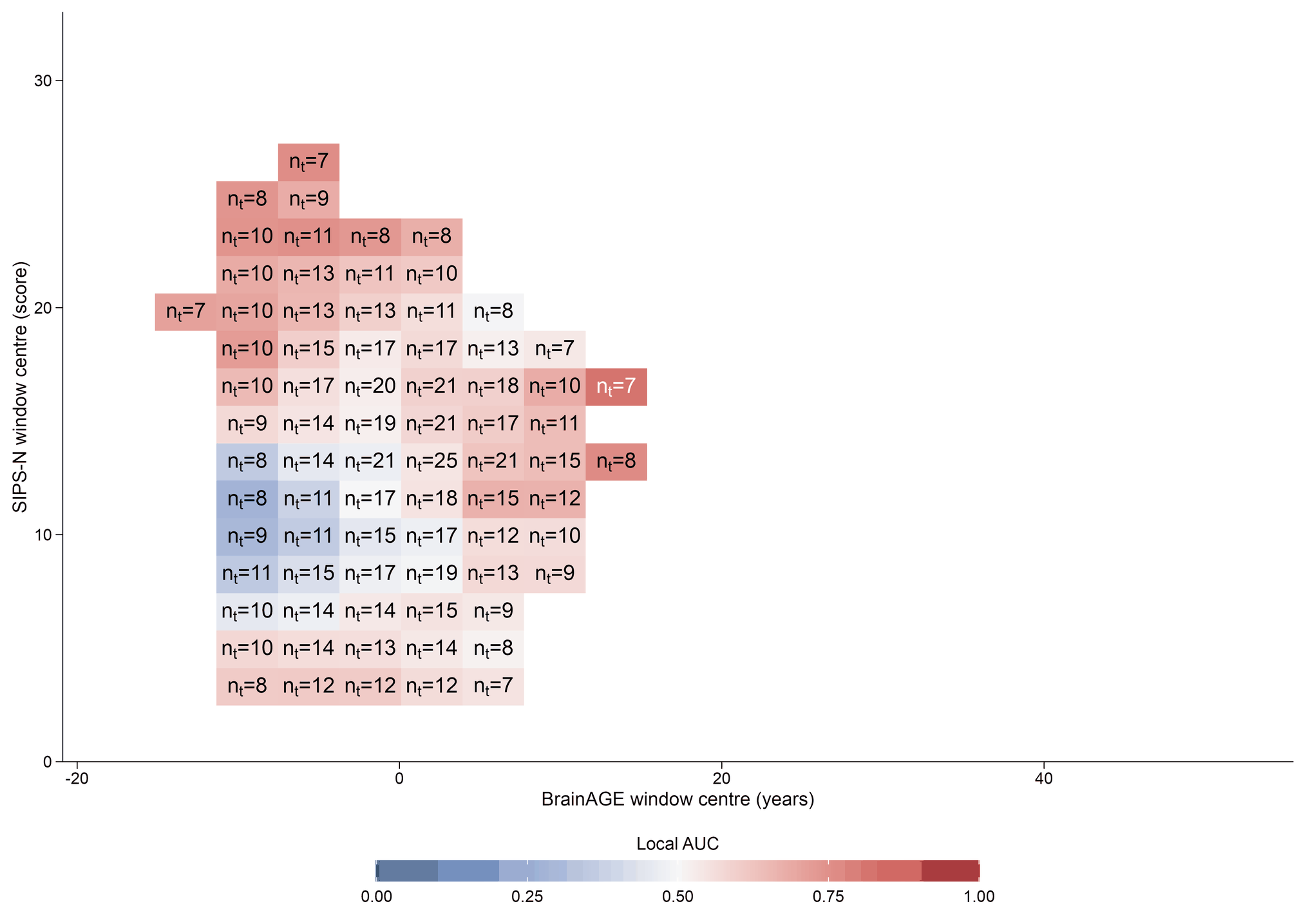

eFigure 10. Exploratory local discrimination across BrainAGE and negative symptoms. Colors show local AUC in intersections of 20%-of-range windows with 5%-of-range steps. The x-axis is BrainAGE (years); the y-axis is SIPS-N. Cell labels give the number of transitions (nt) in each supported cell; unlabeled areas fail the minimum of seven in each outcome group. Colors are centered on chance-level discrimination (AUC 0.5, white). The analysis population contains 670 participants and 84 transitions, with 652 participants and 79 transitions represented by the 76 eligible cells. Dmax = 0.54; within-cohort paired-moderator permutation *P* = .03; Benjamini-Hochberg *q* = .15 across six surfaces.

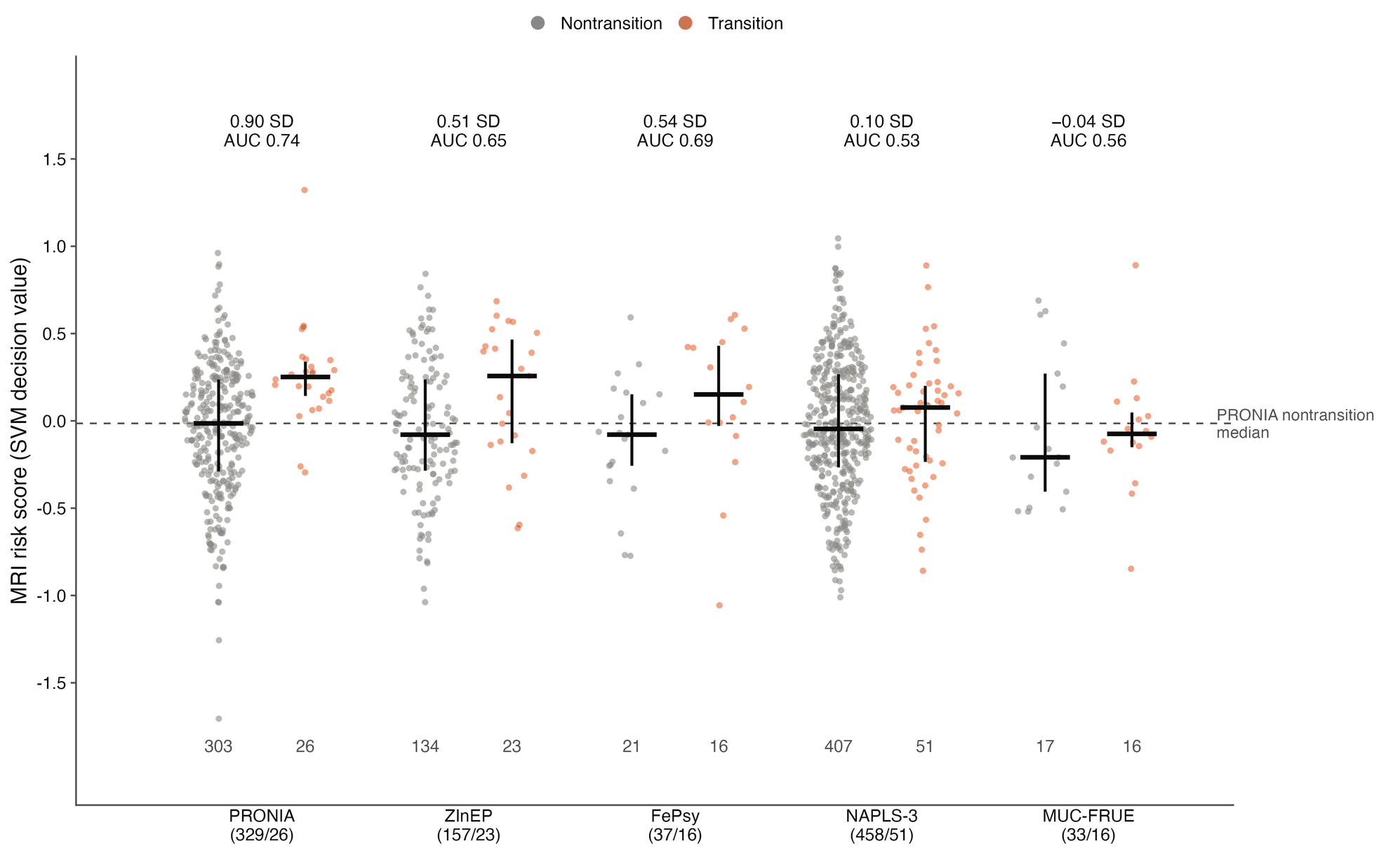

eFigure 11. MRI score of each participant by cohort and outcome. Points show individual participants without (gray) and with (orange) transition, spread horizontally in proportion to the local density. Black lines show medians and interquartile ranges, and numbers below give group sizes. Values above give the separation in SDs, as in Figure 2A, and the AUC. The dashed line marks the PRONIA nontransition median.

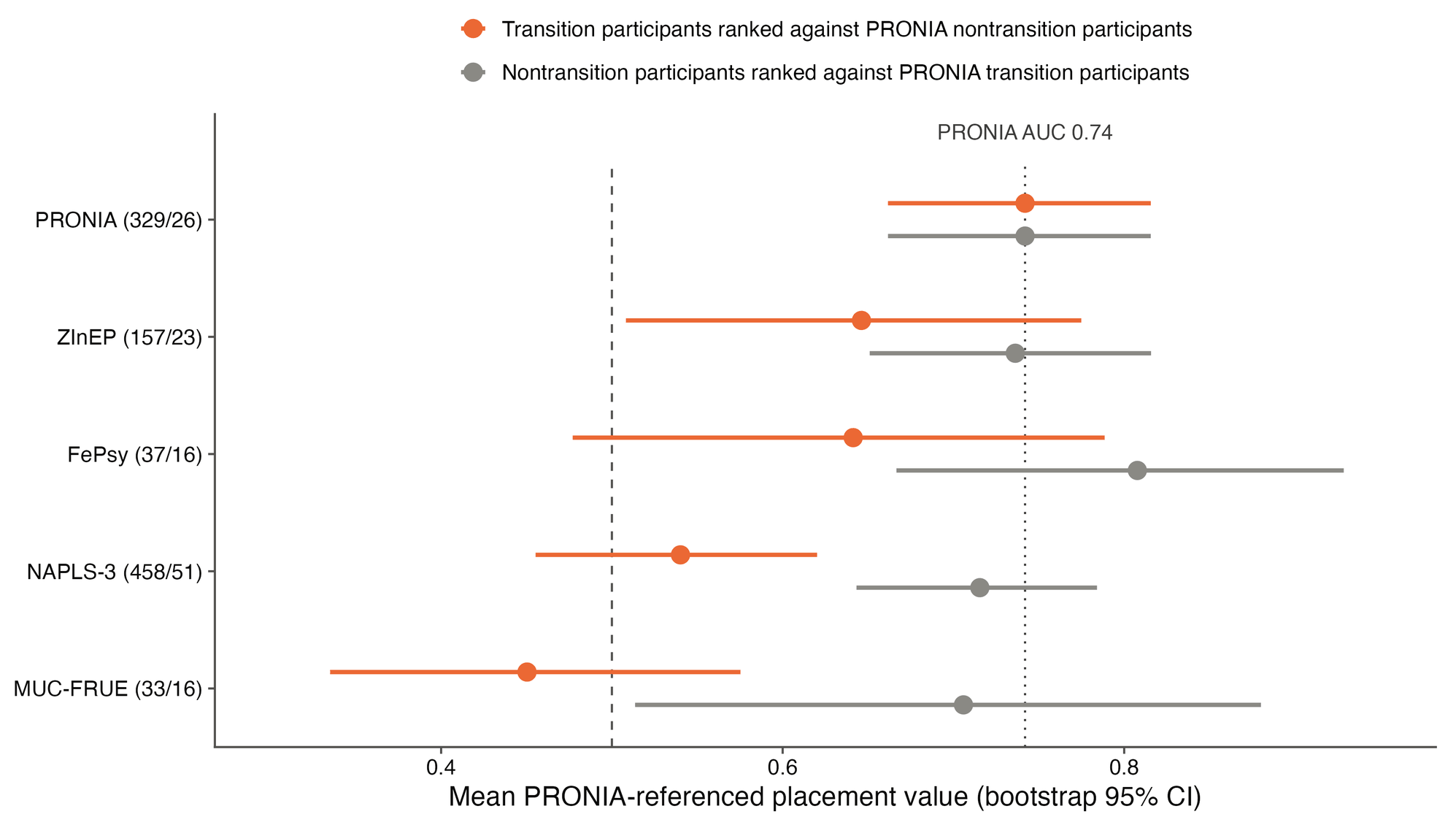

eFigure 12. Transition and nontransition sides of discrimination. Mean PRONIA-referenced placement values with bootstrap 95% CIs (2000 replicates within cohort and outcome; eMethods 12). Orange shows participants who developed psychosis ranked against PRONIA nontransition participants, and gray shows nontransition participants ranked against PRONIA transition participants. Both equal the PRONIA AUC (dotted line) if a cohort ranks participants as PRONIA does, and the dashed line marks chance. Participants/transitions are given in parentheses.

### eTables

eTable 1. TRIPOD+AI reporting checklist.^29^ The checklist items are mapped to the main text and this supplement. D/E indicates development/evaluation. A reporting location does not by itself establish low risk of bias; incomplete items are identified explicitly.

| **Section / item** | **D/E** | **Checklist recommendation** | **Reporting location / limitations** |
| --- | --- | --- | --- |
| Title / 1 | D;E | Identify the study as developing or evaluating the performance of a multivariable prediction model, the target population, and the outcome to be predicted | Title: external validation of a brain imaging model for predicting psychosis in clinical high-risk cohorts. |
| Abstract / 2 | D;E | See TRIPOD+AI for Abstracts checklist | Structured Abstract: cohorts, participants and transitions, AUC with 95% CIs, calibration slope, net benefit, added value beyond symptoms, scores by outcome group and conclusions; threshold metrics in Results and Table 2. |
| Background / 3a | D;E | Explain the healthcare context (including whether diagnostic or prognostic) and rationale for developing or evaluating the prediction model, including references to existing models | Introduction: CHR-P setting, transportability of imaging models and the original PRONIA model. |
| Background / 3b | D;E | Describe the target population and the intended purpose of the prediction model in the context of the care pathway, including its intended users (e.g., healthcare professionals, patients, public) | Introduction; Discussion; eTable 5: research evaluation in CHR-P services, intended users and out-of-scope clinical uses. |
| Background / 3c | D;E | Describe any known health inequalities between sociodemographic groups | No dedicated health-inequality analysis. Sex strata are descriptive (eTable 15), not fairness or invariance validation; race and ethnicity were not available. |
| Objectives / 4 | D;E | Specify the study objectives, including whether the study describes the development or validation of a prediction model (or both) | Introduction, final paragraph: transfer of discrimination and calibration, added value beyond symptoms, and explanations for any loss; exploratory analyses in Methods, Statistical Analysis. |
| Data / 5a | D;E | Describe the sources of data separately for the development and evaluation datasets (e.g., randomized trial, cohort, routine care or registry data), the rationale for using these data, and representativeness of the data | Methods, Study Design and Participants; Table 1; eTable 3: 5 longitudinal cohorts (training cohort, 2 cohorts used for external validation in the original report, 2 cohorts not used before). |
| Data / 5b | D;E | Specify the dates of the collected participant data, including start and end of participant accrual; and, if applicable, end of follow-up | Abstract; Methods, Study Design and Participants: recruitment period of PRONIA and analysis dates; eTable 3: published recruitment periods and follow-up schedules. |
| Participants / 6a | D;E | Specify key elements of the study setting (e.g., primary care, secondary care, general population) including the number and location of centers | Methods, Study Design and Participants; eTables 3 and 4: recruitment settings and MRI acquisition sites. eTable 3 distinguishes source-study settings from the analyzed populations. |
| Participants / 6b | D;E | Describe the eligibility criteria for study participants | Methods, Study Design and Participants; eMethods 1; eTable 3. PRONIA is transdiagnostic; the external cohorts used cohort-specific CHR-P criteria. |
| Participants / 6c | D;E | Give details of any treatments received, and how they were handled during model development or evaluation, if relevant | eTable 3 describes the available treatment context. Treatment was not harmonized or analyzed (Limitations). |
| Data preparation / 7 | D;E | Describe any data pre-processing and quality checking, including whether this was similar across relevant sociodemographic groups | Methods, MRI Model; eMethods 2 and 9; eTable 4: CAT12 r1207, SPM12 and NeuroMiner 1.4. Image quality: eMethods 11; eTable 20. |
| Outcome / 8a | D;E | Clearly define the outcome that is being predicted and the time horizon, including how and when assessed, the rationale for choosing this outcome, and whether the method of outcome assessment is consistent across sociodemographic groups | Methods, Outcome and Symptom Ratings; eTable 3: cohort-defined transition to psychosis during cohort-specific follow-up. No common time horizon; time to transition was not modeled (Limitations). |
| Outcome / 8b | D;E | If outcome assessment requires subjective interpretation, describe the qualifications and demographic characteristics of the outcome assessors | Methods, Outcome and Symptom Ratings; eMethods 1; eTable 3: instruments and criteria. Assessor qualifications and demographic characteristics are not reported. |
| Outcome / 8c | D;E | Report any actions to blind assessment of the outcome to be predicted | Not reported by the cohorts. The MRI score was computed retrospectively by a fixed pipeline and was not available to outcome assessors. |
| Predictors / 9a | D | Describe the choice of initial predictors (e.g., literature, previous models, all available predictors) and any pre-selection of predictors before model building | Methods, MRI Model; eMethods 2; original report. Architecture and training procedure were retained after CAT12 reprocessing. |
| Predictors / 9b | D;E | Clearly define all predictors, including how and when they were measured (and any actions to blind assessment of predictors for the outcome and other predictors) | Methods, Outcome and Symptom Ratings, MRI Model, and Statistical Analysis; eMethods 2 and 9 define the baseline MRI score, BrainAGE and clinical moderators; eTables 3, 4 and 12 describe instruments, acquisition and availability. |
| Predictors / 9c | D;E | If predictor measurement requires subjective interpretation, describe the qualifications and demographic characteristics of the predictor assessors | MRI scoring is computational. Clinical instruments are identified; assessor characteristics and training details are not complete. |
| Sample size / 10 | D;E | Explain how the study size was arrived at (separately for development and evaluation), and justify that the study size was sufficient to answer the research question. Include details of any sample size calculation | Reported in this checklist; Table 1: all available participants, with participants and transitions per cohort. No sample-size calculation was performed for this secondary analysis; precision is addressed in Limitations. |
| Missing data / 11 | D;E | Describe how missing data were handled. Provide reasons for omitting any data | Methods, Statistical Analysis (Missing data and software); eFigure 1; eTable 12: complete observations without imputation and availability of clinical ratings by cohort. |
| Analytical methods / 12a | D | Describe how the data were used (e.g., for development and evaluation of model performance) in the analysis, including whether the data were partitioned, considering any sample size requirements | Methods, MRI Model and Statistical Analysis: PRONIA leave-one-site-out out-of-training scores; unchanged application to the external cohorts; exploratory analyses reported separately. |
| Analytical methods / 12b | D | Depending on the type of model, describe how predictors were handled in the analyses (functional form, rescaling, transformation, or any standardization). | Methods, MRI Model; eMethods 2 and 6: imaging transformations, score outputs and standardization within analysis populations. |
| Analytical methods / 12c | D | Specify the type of model, rationale, all model-building steps, including any hyperparameter tuning, and method for internal validation | Methods, MRI Model; eMethods 2; original report: nested leave-one-site-out cross-validation and class-weighted linear support vector machine. |
| Analytical methods / 12d | D;E | Describe if and how any heterogeneity in estimates of model parameter values and model performance was handled and quantified across clusters (e.g., hospitals, countries). See TRIPOD-Cluster for additional considerations | Methods, Statistical Analysis; eMethods 4 and 11; eTables 15 and 20: random-effects meta-analysis, comparison of never-used and previously used cohorts, and within-site analyses in NAPLS-3. |
| Analytical methods / 12e | D;E | Specify all measures and plots used (and their rationale) to evaluate model performance (e.g., discrimination, calibration, clinical utility) and, if relevant, to compare multiple models | Methods, Statistical Analysis; Table 2; Figure 1; eTables 13 and 15: discrimination, threshold metrics, calibration, Brier score, net benefit and uncertainty methods. |
| Analytical methods / 12f | E | Describe any model updating (e.g., recalibration) arising from the model evaluation, either overall or for particular sociodemographic groups or settings | Methods, Statistical Analysis; eMethods 4; eTable 15; eFigures 2 and 6: cohort-specific intercept updating, held-out recalibration and apparent pooled updating, reported separately. |
| Analytical methods / 12g | E | For model evaluation, describe how the model predictions were calculated (e.g., formula, code, object, application programming interface) | Methods, MRI Model; eMethods 2 and 4: fitted transformations, score aggregation, class voting and Platt probabilities. The fitted model can be requested from the corresponding author (Data Sharing Statement). |
| Class imbalance / 13 | D;E | If class imbalance methods were used, state why and how this was done, and any subsequent methods to recalibrate the model or the model predictions | Methods, MRI Model; eMethods 4: class-weighted support vector machine and Platt scaling with inverse class-frequency weighting, fitted to PRONIA out-of-training scores. |
| Fairness / 14 | D;E | Describe any approaches that were used to address model fairness and their rationale | No dedicated fairness method was used. Sex-stratified estimates are descriptive (eTable 15). |
| Model output / 15 | D | Specify the output of the prediction model (e.g., probabilities, classification). Provide details and rationale for any classification and how the thresholds were identified | Methods, MRI Model; Table 2; eTable 5: continuous score, original class prediction and Platt probability; the decision threshold was not retuned. |
| Training versus evaluation / 16 | D;E | Identify any differences between the development and evaluation data in healthcare setting, eligibility criteria, outcome, and predictors | Results, Participants; Discussion; Table 1; eTables 3, 4 and 12: participant characteristics, outcome definition, follow-up, acquisition and availability of clinical ratings. |
| Ethical approval / 17 | D;E | Name the institutional research board or ethics committee that approved the study and describe the participant-informed consent or the ethics committee waiver of informed consent | Methods, Study Design and Participants; eMethods 1: consent procedures and approving ethics committees of the 5 cohorts. |
| Funding / 18a | D;E | Give the source of funding and the role of the funders for the present study | Article Information: Funding/Support; Role of the Funder/Sponsor. |
| Conflicts of interest / 18b | D;E | Declare any conflicts of interest and financial disclosures for all authors | Article Information: Conflict of Interest Disclosures. |
| Protocol / 18c | D;E | Indicate where the study protocol can be accessed or state that a protocol was not prepared | Reported in this checklist: no separate protocol was prepared for this secondary analysis. |
| Registration / 18d | D;E | Provide registration information for the study, including register name and registration number, or state that the study was not registered | Reported in this checklist: the study was not preregistered. eMethods 1: DRKS00005042 identifies the source PRONIA study, not this analysis. |
| Data sharing / 18e | D;E | Provide details of the availability of the study data | Article Information: Data Sharing Statement, with access routes for each cohort's data. |
| Code sharing / 18f | D;E | Provide details of the availability of the analytical code | Methods, Statistical Analysis (Missing data and software); Article Information: Data Sharing Statement; eTable 5: analysis code and application repositories. |
| Patient & Public Involvement / 19 | D;E | Provide details of any patient and public involvement during the design, conduct, reporting, interpretation, or dissemination of the study or state no involvement. | Reported in this checklist: patients and members of the public were not involved in the design, analysis or reporting of this secondary analysis. |
| Participants / 20a | D;E | Describe the flow of participants through the study, including the number of participants with and without the outcome and, if applicable, a summary of the follow-up time. A diagram may be helpful. | eFigure 1; Results, Participants; Table 1: analyzed populations by cohort; published follow-up schedules are summarized in eTable 3. |
| Participants / 20b | D;E | Report the characteristics overall and, where applicable, for each data source or setting, including the key dates, key predictors (including demographics), treatments received, sample size, number of outcome events, follow-up time, and amount of missing data. A table may be helpful. Report any differences across key demographic groups. | Table 1; eTables 3, 4 and 12: cohort characteristics, participants and transitions, published accrual and follow-up information, and availability of clinical ratings. Treatment was not harmonized; race and ethnicity were not available. |
| Participants / 20c | E | For model evaluation, show a comparison with the development data of the distribution of important predictors (demographics, predictors, and outcome). | Results, Participants; Table 1; eTable 12: cohort characteristics compared with PRONIA; Table 2 compares performance. |
| Model development / 21 | D;E | Specify the number of participants and outcome events in each analysis (e.g., for model development, hyperparameter tuning, model evaluation) | Tables 1 and 2; eTables 6, 7, 12, 16, 18 and 19: participants and transitions in each analysis. |
| Model specification / 22 | D | Provide details of the full prediction model (e.g., formula, code, object, application programming interface) to allow predictions in new individuals and to enable third-party evaluation and implementation, including any restrictions to access or re-use (e.g., freely available, proprietary) | Methods, MRI Model; eMethods 2; eTable 5; Article Information: Data Sharing Statement (fitted model available on request). |
| Model performance / 23a | D;E | Report model performance estimates with confidence intervals, including for any key subgroups (e.g.,sociodemographic). Consider plots to aid presentation. | Table 2; Figure 1; eTable 15: AUC, calibration and net benefit with 95% CIs, and descriptive sex strata (eTable 15); local estimates in eFigures 9 and 10 and eTable 17. |
| Model performance / 23b | D;E | If examined, report results of any heterogeneity in model performance across clusters. See TRIPOD Cluster for additional details. | Results, External Discrimination; Table 2; eTables 15, 17 and 18: random-effects summary, prediction interval and exploratory performance heterogeneity. |
| Model updating / 24 | E | Report the results from any model updating, including the updated model and subsequent performance | Results, Calibration and Clinical Utility; eTable 15; eFigures 2 and 6: apparent and held-out probability updating; the classifier itself was not updated. |
| Interpretation / 25 | D;E | Give an overall interpretation of the main results, including issues of fairness in the context of the objectives and previous studies | Discussion: lower discrimination in all external cohorts, close to chance in NAPLS-3; loss confined to participants who developed psychosis; scanner effects, classifiers, age and sex considered; 3 explanations (overstated training estimate, signal in only some people, transition definition and follow-up); fairness was not assessed. |
| Limitations / 26 | D;E | Discuss any limitations of the study (such as a non-representative sample, sample size, overfitting, missing data) and their effects on any biases, statistical uncertainty, and generalizability | Discussion, Limitations; eDiscussion; eTable 2. |
| Usability of the model in the context of current care / 27a | D | Describe how poor quality or unavailable input data (e.g., predictor values) should be assessed and handled when implementing the prediction model | eTable 5: scan-quality and compatibility requirements; image-quality analyses in eTable 20. |
| Usability of the model in the context of current care / 27b | D | Specify whether users will be required to interact in the handling of the input data or use of the model, and what level of expertise is required of users | eTable 5: specialist MRI processing and CHR-P assessment expertise; the score is not suitable for stand-alone clinical use. |
| Usability of the model in the context of current care / 27c | D;E | Discuss any next steps for future research, with a specific view to applicability and generalizability of the model | Discussion; eTable 5: prospective evaluation of the MRI component, validation in several cohorts, and shared transition definitions with a common minimum follow-up. |

eTable 2. Narrative assessment of bias and applicability. This is a descriptive assessment informed by PROBAST+AI,^30^ not a completed standardized signalling-question assessment or an assigned overall risk-of-bias rating.

| **Domain** | **Assessment focus / concerns** | **Applicability and interpretation** |
| --- | --- | --- |
| Participants | Recruitment, eligibility, analytic selection and exclusions. Case selection and incomplete source-flow reporting. | Population differences across settings. Five longitudinal cohorts were included. The PRONIA sample also contained recent-onset depression participants. The analyzed sample depended on MRI, outcome and covariate availability; these selections may influence performance estimates. |
| Predictors | Baseline MRI, processing, scoring and clinical measurement. Implementation and measurement differences; incomplete documentation of some clinical measures. | Scanner/protocol and preprocessing compatibility. The re-estimated classifier was applied without validation-cohort-specific refitting. Reprocessing version, image reliability filtering, PCA/SVM scoring, Platt scaling and BrainAGE need reproducible specification. Clinical moderators were not uniformly available. |
| Outcome | Transition definition, timing, observation period, treatment and blinding. Heterogeneous endpoints and follow-up; unharmonized treatment. | No single common prediction horizon. Binary transition during cohort-specific follow-up combines different observation periods and assessment procedures. Outcome coding should be distinguished from the broader study protocols. Blinding information is incomplete. |
| Analysis | Event counts, model version, missingness, calibration, inference and multiplicity. Small event counts, complete-case selection, overlapping local samples and multiple exploratory tests. | Exploratory findings do not establish clinical subgroups. Primary external validation and pooled exploratory analyses serve different roles. ZInEP and FePsy had been benchmarked previously. One-dimensional global tests did not survive post hoc correction; surface and component results are nominal. Apparent recalibration is distinct from the separately reported held-out updating analysis (eTable 15). |
| Overall | Synthesis across domains. Narrative summary only; no standardized overall PROBAST+AI judgment. | Research evaluation; not ready for individual clinical use. The multi-cohort study documents limited transportability. It does not isolate a cause of model failure, establish a neurobiological mechanism, or demonstrate that local subgroup selection would improve care. |

Explicit statistical corrections do not remove concerns arising from selection, missing follow-up or measurement differences. Conversely, heterogeneity is not by itself proof of an implementation error. The clinical conclusion is limited external validity without an established reproducible subgroup.

eTable 3. Cohort criteria, outcomes, and follow-up. The analyzed endpoint in every cohort is transition to psychosis during its cohort-specific observation period. Broader source-study eligibility does not redefine the analyzed endpoint. Cohort protocols, calendar dates, recruitment settings, source-study flow and treatment context are summarized from the cited study descriptions.

| **Cohort** | **CHR-P criteria / instrument** | **Source-study risk syndromes** | **Analyzed transition endpoint** | **Follow-up schedule** | **Treatment context** | **Recruitment dates** |
| --- | --- | --- | --- | --- | --- | --- |
| PRONIA^1^ | SIPS for UHR criteria and SPI-A for basic symptoms; CHR subset included participants meeting UHR criteria and/or cognitive basic symptom criteria. | APS, BLIPS, GRFD, and COGDIS. | Transition to psychosis during follow-up. Defined as a SIPS positive symptom item reaching psychotic intensity daily for >= 7 days. | Published follow-up ranged from 9 to 36 months, with visits every 3 months through month 18 and every 9 months thereafter.^1^ | Baseline antipsychotic exposure above guideline threshold excluded. | Recruitment: 1 February 2014 to 31 May 2017.^1^ |
| NAPLS-3^31^ | Criteria of Psychosis-Risk Syndromes (COPS) assessed with SIPS; North American consortium. Parent-study eligibility was age 12-35 years; the analyzed NAPLS-3 participants were aged 12-30 years. | COPS/SIPS psychosis-risk syndromes. | Transition to psychosis during NAPLS-3 follow-up. Defined by the SIPS Presence of Psychotic Symptoms criteria, psychotic intensity for at least 1 hour a day on an average of 4 days per week for 1 month, or a seriously disorganizing or dangerous symptom. | Clinical and biomarker assessments every 2 months for first 8 months; clinical assessments at 12, 18, and 24 months; 2-year follow-up. | Psychotropic medication use was systematically recorded at baseline and during follow-up. | Recruitment: February 2015 to November 2018.^32^ |
| MUC-FRUE^2^ | Munich LMU early-detection cohort, recruited at the Early Detection and Intervention Center for Mental Crises; two-stage ARMS framework. | Early ARMS/basic-symptom states and late ARMS/PACE-defined states, including attenuated psychotic symptoms and/or brief limited intermittent psychotic symptoms. | Transition defined using Yung/PACE-like PANSS thresholds: hallucinations, unusual thought content, suspiciousness, or conceptual disorganization reaching psychotic intensity and occurring daily for more than one week, with later ICD-10 schizophrenia-spectrum confirmation. | Four-year longitudinal follow-up; visits weekly in the first month, monthly in the first year, quarterly in the second year, and annually thereafter. | Participants reported as antipsychotic-naive before baseline testing. | Not reported; recruitment was completed before July 2008, when the 2009 classification study was submitted.^2^ |
| ZInEP^33,34^ | Zurich early-recognition cohort; SIPS for UHR criteria and SPI-A/SPI-CY for basic symptoms; participants aged 13-35 from Canton Zurich study centers. | APSS, BLIPS, state-trait/GRFD-like criteria, COPER/basic symptoms, COGDIS/cognitive disturbances, with a bipolar-risk pathway in the broader protocol (not a combined outcome in this analysis). | Transition to psychosis during follow-up. Defined according to ICD-10 criteria. | Follow-up assessments at 6, 12, 24, and 36 months, or earlier if transition occurred. | Naturalistic treatment setting. Psychotropic medication and psychotherapeutic interventions were permitted and tracked as part of the multi-level study approach. | Not reported; screening took 28 months and began after ethics approval in 2009 (KEK-ZH-Nr. E-63/2009).^34^ |
| FePsy^35^ | Basel FePsy cohort; BSIP/BIP-based at-risk framework with stepwise screening and clinical interviews. | FePsy-specific ARMS criteria / BSIP-defined at-risk mental state. | Transition defined using BPRS according to Yung/PACE criteria. | Monthly in year 1, every 3 months in years 2-3, then yearly until transition or the published follow-up endpoint in 2007.^36^ | Needs-based treatment, case management, and supportive psychotherapy during follow-up; antipsychotic treatment only after transition. | Study inclusion: 1 March 2000 to 29 February 2004.^36^ |

CHR-P, clinical high risk for psychosis; SIPS, Structured Interview for Prodromal Syndromes; SPI-A/SPI-CY, Schizophrenia Proneness Instrument, adult/child-youth versions; APS/APSS, attenuated psychotic symptoms (syndrome); BLIPS, brief limited intermittent psychotic symptoms; GRFD, genetic risk and functioning decline; COGDIS, cognitive disturbances; COPS, Criteria of Psychosis-Risk Syndromes; ARMS, at-risk mental state; PACE, Personal Assessment and Crisis Evaluation; PANSS, Positive and Negative Syndrome Scale; BSIP/BIP, Basel screening/interview instruments; BPRS, Brief Psychiatric Rating Scale. PRONIA additionally included recent-onset depression participants. For orientation, the external AUCs were 0.53 in NAPLS-3, 0.56 in MUC-FRUE, 0.65 in ZInEP, and 0.69 in FePsy, compared with 0.74 in PRONIA (Figure 1A). Because the cohorts differ in criteria, follow-up, recruitment, and scanners at the same time, this comparison is descriptive.

eTable 4. Structural MRI acquisition parameters. Published baseline T1-weighted protocols are summarized with their source citations.

| **Cohort** | **Site / group** | **Scanner model** | **Field** | **Flip (°)** | **Coil** | **Voxel size (mm)** | **TR (ms)** | **TE (ms)** | **FOV (mm)** | **Slices** |
| --- | --- | --- | --- | --- | --- | --- | --- | --- | --- | --- |
| PRONIA^1^ | Munich | Philips Ingenia | 3T | 8 | 32 | 0.97 x 0.97 x 1 | 9.5 | 5.5 | 250 x 250 | 190 |
| PRONIA | Milan Niguarda | Philips Achieva Intera | 1.5T | 12 | 8 | 0.93 x 0.93 x 1 | Shortest (8.1) | Shortest (3.7) | 240 x 240 | 170 |
| PRONIA | Cologne | Philips Achieva | 3T | 8 | 8 | 0.97 x 0.97 x 1 | 9.5 | 5.5 | 250 x 250 | 190 |
| PRONIA | Basel | Siemens Verio | 3T | 8 | 12 | 1 x 1 x 1 | 2000 | 3.4 | 256 x 256 | 176 |
| PRONIA | Birmingham | Philips Achieva | 3T | 8 | 32 | 1 x 1 x 1 | 8.4 | 3.8 | 288 x 288 | 175 |
| PRONIA | Turku | Philips Ingenuity | 3T | 7 | 32 | 1 x 1 x 1 | 8.1 | 3.7 | 256 x 256 | 176 |
| PRONIA | Udine | Philips Achieva | 3T | 12 | 8 | 0.93 x 0.93 x 1 | Shortest (8.1) | Shortest (3.7) | 240 x 240 | 170 |
| NAPLS-3^22^ | Sites 1, 5 | Siemens Prisma Fit | 3T | 8 | 32 | 1-mm slices | 2400 | 1.96 | 256 | NR |
| NAPLS-3^22^ | Site 8 | Siemens Prisma Fit | 3T | 8 | 64 | 1-mm slices | 2400 | 1.96 | 256 | NR |
| NAPLS-3^22^ | Site 4 | Siemens Prisma | 3T | 8 | 64 | 1-mm slices | 2400 | 1.96 | 256 | NR |
| NAPLS-3^22^ | Site 9 | Siemens Skyra | 3T | 8 | 32 | 1-mm slices | 2400 | 1.96 | 256 | NR |
| NAPLS-3^22^ | Sites 1, 2, 3, 5, 8 | Siemens Trio Tim | 3T | 8 | 32 | 1-mm slices | 2400 | 1.90 | 256 | NR |
| NAPLS-3^22^ | Site 5 | Siemens Biograph mMR | 3T | NR | 12 | 1-mm slices | NR | NR | 256 | NR |
| NAPLS-3^22^ | Site 4 | GE Signa HDxt | 3T | 11 | 8 | 1-mm slices | 4.27 | 1.76 | 256 | NR |
| NAPLS-3^22^ | Sites 6, 7 | GE Discovery MR750 (first entry) | 3T | 11 | NR | 1-mm slices | 4.00 | 1.33 | 256 | NR |
| NAPLS-3^22^ | Sites 6, 7 | GE Discovery MR750 (second entry) | 3T | 11 | NR | 1-mm slices | 4.64 | 2.00 | 256 | NR |
| MUC-FRUE^2^ | Munich LMU early-detection cohort | Siemens Magnetom Vision | 1.5T | NR | NR | 0.45 x 0.45 x 1.5 | 11.6 | 4.9 | 230 x 230 | 126 |
| ZInEP^37^ | Zurich; two identical scanners | Philips Achieva TX | 3T | 8 | 8 | 1 x 1 x 1 | 8.3 | 3.8 | 240 x 240 | 160 |
| FePsy^36^ | University Hospital Basel | Siemens Magnetom Vision | 1.5T | 12 | NR | 1.28 x 1 x 1 | 9.7 | 4 | 256 x 256 | 176 |

NR, not reported in the cited acquisition description. TR, repetition time; TE, echo time; FOV, field of view. Field strength is in tesla; flip angle is in degrees; coil entries give channel counts. For NAPLS-3, the voxel-size column gives the reported slice thickness. Values describe acquisition, not the subsequent 3-mm analysis resolution.

eTable 5. Model card for the PRONIA-derived structural MRI transition-risk score. This summarizes the evaluated model, its research context, observed performance, limitations and safeguards, not a clinical deployment policy.^38^

| **Model identity** | |
| --- | --- |
| **Field** | **Description** |
| Model name | PRONIA-derived structural MRI psychosis transition risk score. |
| Model type | Baseline structural MRI-based machine-learning risk score. The model uses a linear support-vector machine (SVM) trained on gray-matter volume features after dimensionality reduction. Platt-scaled probabilities were generated by the fixed PRONIA-derived SVM pipeline and used for calibration analyses; additional post hoc probability-updating analyses are reported separately as apparent and held-out estimates. |
| Model origin and version | PRONIA training cohort; pipeline re-estimated after CAT12 r1207 preprocessing rather than the original r1155 fitted pipeline.^1^ |
| Developers / contact | Developed by Nikolaos Koutsouleris and the PRONIA consortium.^1^ |
| training-cohort citation | ^1^ |
| **Intended use and out-of-scope use** | |
| **Field** | **Description** |
| Primary intended use | Research evaluation and clinical-decision-support development in early recognition and clinical high-risk for psychosis (CHR-P) settings, following local validation, calibration assessment, and implementation testing. The score is not intended for immediate standalone clinical deployment. |
| Intended users | Researchers and early-detection clinicians with access to structural MRI preprocessing, model-scoring infrastructure, and expertise in CHR-P assessment. |
| Out-of-scope uses | General-population screening; standalone diagnosis of psychosis; standalone treatment allocation; individual absolute-risk communication without local validation and appropriate updating; application to scans failing MRI quality control; use in substantially different populations, endpoints, scanner environments, or preprocessing pipelines without local validation. |
| **Required inputs, outputs, and thresholds** | |
| **Field** | **Description** |
| Required inputs | Baseline T1-weighted structural MRI acquired with sufficient quality for the fixed preprocessing/scoring pipeline. Structural MRI must pass quality control for scanner artifacts, gross anatomical abnormalities, preprocessing failures, and compatibility with the model input space. |
| Model outputs | Continuous MRI risk score; inherited ensemble class prediction based on majority vote; Platt-scaled probability estimates for calibration analyses only. |
| Decision threshold | The fixed SVM operating point was inherited from the training-cohort PRONIA setting and was not retuned in external validation cohorts. Threshold-dependent metrics should therefore be interpreted as fixed-threshold transportability estimates. |
| **Evaluation data and factors** | |
| **Field** | **Description** |
| training-cohort data | PRONIA: 329 participants, 26 transitions; out-of-training predictions. The PRONIA population included CHR-P and recent-onset depression. |
| External validation data | NAPLS-3, MUC-FRUE, ZInEP and FePsy: 685 participants, 106 transitions. Pooled PRONIA and external sample: 1014 participants, 132 transitions. ZInEP and FePsy were previously used; NAPLS-3 and MUC-FRUE had not been used to evaluate the model. |
| Evaluation factors | Cohort, recorded sex, age, BrainAGE, SIPS-P, SIPS-N, COGDIS and functioning. Availability differs by cohort. The SIPS-N linear interaction is nominal but not adjusted-significant; the GAM tensor meets one within-model family but is penalty-sensitive and not corrected across forms. Clinical-cohort BrainAGE errors and residual age dependence qualify interpretation. |
| **Validation results summarized for interpretation** | |
| **Field** | **Description** |
| Average discrimination | PRONIA AUC 0.742 (95% CI, 0.662-0.822), BACC 0.724. External pooled AUC 0.563 (95% CI, 0.505-0.622), BACC 0.558; PPV 0.187. Random-effects external AUC 0.587 (Hartung-Knapp 95% CI, 0.461-0.702). Pooled and meta-analytic AUCs are distinct summaries. |
| Calibration | Original external Brier 0.245, calibration slope 0.166 and calibration-in-the-large intercept -1.731. The external sample-prevalence Brier reference is 0.131. Apparent pooled intercept/slope updating gives Brier 0.130, not independent improvement. PRONIA probabilities are also poorly calibrated (Brier 0.215 versus reference 0.073). |
| Context-dependent performance | Local AUC estimates vary across contexts, but none of the six primary one-dimensional or six surface-level tests survives its family correction. Minimum one-dimensional q=0.073; SIPS-N × BrainAGE surface p=0.025, q=0.148. Magnitude/support depend on population and window settings; no reproducible subgroup was established. |
| Fairness/subgroup audit | Recorded-sex AUCs in the combined PRONIA and external population are 0.602 (male; 526 participants, 80 transitions) and 0.598 (female; 488 participants, 52 transitions), with overlapping intervals. This is not an external-only fairness analysis or a demonstration of equal performance. |
| **Ethical considerations and cautions** | |
| **Field** | **Description** |
| Clinical consequences | The model targets transition to psychosis in help-seeking CHR-P individuals, a clinically consequential endpoint. Misclassification could affect prognostic counseling, intervention planning, and access to early-intervention services. The score is not a standalone clinical decision rule. |
| Main risks | Weak external discrimination; poor calibration in source and external populations; heterogeneous endpoints/follow-up; scanner/protocol differences; sparse local cells; complete-case selection; overlapping samples; multiple exploratory families; GAM concurvity/penalty sensitivity; unharmonized treatment. |
| Component interpretation | The four components describe partly overlapping structural patterns contributing to the classifier. |
| **Recommended safeguards before reuse** | |
| **Field** | **Description** |
| Minimum validation checks | Before reuse in a new setting, report AUC with confidence intervals; BACC, sensitivity, and specificity at the fixed operating point; Brier score; calibration intercept and slope; subgroup/fairness audits; MRI quality-control failures; and endpoint/follow-up comparability with the validation cohorts. |
| Updating strategy | Distinguish probability recalibration from refitting the neuroanatomical decision function. Probability outputs should not be used for absolute-risk communication without local validation and, where appropriate, local updating or recalibration. Updating should be evaluated prospectively or with internal-external validation before clinical use. |
| Monitoring if implemented | Not applicable to current clinical use: the evaluated score is not ready for deployment. Any future implementation requires independent validation and an explicit monitoring plan. |
| **Availability and documentation** | |
| **Field** | **Description** |
| Data availability | Participant-level clinical and imaging data are subject to cohort-specific consent, ethics approvals and data-use agreements. Cohort-specific access routes and requests for the study-specific derived risk scores and harmonized analysis data are described in the Data Sharing Statement. |
| Code, app, and license | The analysis and figure-generation code are available at https://github.com/claravetter/pronia-mri-transition-performance-landscapes under the MIT license. Source code for the performance-landscape application is available at https://github.com/claravetter/prediction-landscape-explorer |
| Reference | ^1^ |

eTable 6. Moderation regressions without functioning and follow-up analyses. All regressions use 658 participants and 84 transitions, with analysis-set standardization. MRI-score interactions, not main effects, define each regression's BH family. Further sections report the MRI score × SIPS-N interaction within each cohort, the baseline clinical profile by cohort and outcome, premorbid adjustment in PRONIA, discrimination in PRONIA by documented timing of transition, and the separation of participants with and without transition on the 4 structural components by cohort.

| **Clinical regression (Model 1)** | | | | | | | | | | | | | | | | | | | | | | | | | | | | | | | | | | | | |
| --- | --- | --- | --- | --- | --- | --- | --- | --- | --- | --- | --- | --- | --- | --- | --- | --- | --- | --- | --- | --- | --- | --- | --- | --- | --- | --- | --- | --- | --- | --- | --- | --- | --- | --- | --- | --- |
| **Term** | | | | | | | | | **β (95% CI)** | | | | | | | | | | | | | **SE** | | | | **p** | | | | | | | | **Interaction q** | | |
| Intercept | | | | | | | | | -2.375 (-2.723 to -2.028) | | | | | | | | | | | | | 0.177 | | | | <0.001 | | | | | | | | — | | |
| MRI risk score | | | | | | | | | 0.262 (0.008 to 0.516) | | | | | | | | | | | | | 0.130 | | | | 0.044 | | | | | | | | — | | |
| COGDIS | | | | | | | | | -0.057 (-0.322 to 0.208) | | | | | | | | | | | | | 0.135 | | | | 0.674 | | | | | | | | — | | |
| SIPS-P | | | | | | | | | 0.491 (0.203 to 0.778) | | | | | | | | | | | | | 0.147 | | | | <0.001 | | | | | | | | — | | |
| SIPS-N | | | | | | | | | 0.278 (0.007 to 0.548) | | | | | | | | | | | | | 0.138 | | | | 0.045 | | | | | | | | — | | |
| PRONIA (vs NAPLS-3) | | | | | | | | | 0.735 (0.117 to 1.352) | | | | | | | | | | | | | 0.315 | | | | 0.020 | | | | | | | | — | | |
| ZInEP (vs NAPLS-3) | | | | | | | | | 1.060 (0.252 to 1.868) | | | | | | | | | | | | | 0.412 | | | | 0.010 | | | | | | | | — | | |
| MRI risk score × COGDIS | | | | | | | | | 0.028 (-0.217 to 0.273) | | | | | | | | | | | | | 0.125 | | | | 0.822 | | | | | | | | 0.822 | | |
| MRI risk score × SIPS-P | | | | | | | | | -0.063 (-0.310 to 0.183) | | | | | | | | | | | | | 0.126 | | | | 0.615 | | | | | | | | 0.822 | | |
| MRI risk score × SIPS-N | | | | | | | | | 0.251 (0.001 to 0.502) | | | | | | | | | | | | | 0.128 | | | | 0.049 | | | | | | | | 0.147 | | |
| *MRI-score simple slopes at SIPS-N levels* | | | | | | | | | | | | | | | | | | | | | | | | | | | | | | | | | | | | |
| **SIPS-N level** | | | | **β (95% CI)** | | | | | | | | | | | | | | | | | | **SE** | | | | | | | | | **p** | | | | | |
| -1 SD | | | | 0.011 (-0.373 to 0.394) | | | | | | | | | | | | | | | | | | 0.196 | | | | | | | | | 0.957 | | | | | |
| Mean | | | | 0.262 (0.008 to 0.516) | | | | | | | | | | | | | | | | | | 0.130 | | | | | | | | | 0.044 | | | | | |
| +1 SD | | | | 0.513 (0.186 to 0.841) | | | | | | | | | | | | | | | | | | 0.167 | | | | | | | | | 0.002 | | | | | |
| **Demographic and BrainAGE extensions** | | | | | | | | | | | | | | | | | | | | | | | | | | | | | | | | | | | | |
| **Term** | | | | | | | **Model 2 Demographic main** | | | | | | | | | | | **Model 2b Demographic interactions** | | | | | | | | | | | | **Model 3 BrainAGE** | | | | | | |
| Intercept | | | | | | | -2.246 (-2.664 to -1.829) p<0.001 | | | | | | | | | | | -2.274 (-2.697 to -1.851) p<0.001 | | | | | | | | | | | | -2.268 (-2.690 to -1.847) p<0.001 | | | | | | |
| MRI risk score | | | | | | | 0.265 (0.009 to 0.520) p=0.042 | | | | | | | | | | | 0.333 (0.001 to 0.665) p=0.049 | | | | | | | | | | | | 0.261 (0.002 to 0.521) p=0.048 | | | | | | |
| COGDIS | | | | | | | -0.063 (-0.328 to 0.202) p=0.640 | | | | | | | | | | | -0.069 (-0.335 to 0.196) p=0.608 | | | | | | | | | | | | -0.071 (-0.335 to 0.194) p=0.600 | | | | | | |
| SIPS-P | | | | | | | 0.506 (0.216 to 0.797) p<0.001 | | | | | | | | | | | 0.507 (0.214 to 0.799) p<0.001 | | | | | | | | | | | | 0.505 (0.213 to 0.798) p<0.001 | | | | | | |
| SIPS-N | | | | | | | 0.285 (0.012 to 0.558) p=0.041 | | | | | | | | | | | 0.302 (0.027 to 0.576) p=0.032 | | | | | | | | | | | | 0.281 (0.008 to 0.554) p=0.044 | | | | | | |
| Age | | | | | | | 0.155 (-0.106 to 0.415) p=0.244 | | | | | | | | | | | 0.108 (-0.169 to 0.386) p=0.445 | | | | | | | | | | | | 0.128 (-0.138 to 0.394) p=0.346 | | | | | | |
| Female (vs male) | | | | | | | -0.203 (-0.691 to 0.285) p=0.415 | | | | | | | | | | | -0.172 (-0.667 to 0.323) p=0.496 | | | | | | | | | | | | -0.213 (-0.703 to 0.277) p=0.394 | | | | | | |
| PRONIA (vs NAPLS-3) | | | | | | | 0.598 (-0.061 to 1.258) p=0.075 | | | | | | | | | | | 0.624 (-0.041 to 1.290) p=0.066 | | | | | | | | | | | | 0.651 (-0.021 to 1.323) p=0.058 | | | | | | |
| ZInEP (vs NAPLS-3) | | | | | | | 0.967 (0.151 to 1.783) p=0.020 | | | | | | | | | | | 0.951 (0.133 to 1.769) p=0.023 | | | | | | | | | | | | 1.014 (0.187 to 1.842) p=0.016 | | | | | | |
| MRI risk score × COGDIS | | | | | | | 0.025 (-0.219 to 0.270) p=0.839; q=0.839 | | | | | | | | | | | 0.024 (-0.223 to 0.271) p=0.849; q=0.901 | | | | | | | | | | | | 0.024 (-0.222 to 0.269) p=0.850; q=0.850 | | | | | | |
| MRI risk score × SIPS-P | | | | | | | -0.053 (-0.300 to 0.193) p=0.671; q=0.839 | | | | | | | | | | | 0.017 (-0.243 to 0.276) p=0.901; q=0.901 | | | | | | | | | | | | -0.055 (-0.304 to 0.195) p=0.668; q=0.850 | | | | | | |
| MRI risk score × SIPS-N | | | | | | | 0.261 (0.010 to 0.512) p=0.042; q=0.126 | | | | | | | | | | | 0.279 (0.027 to 0.530) p=0.030; q=0.148 | | | | | | | | | | | | 0.275 (0.021 to 0.528) p=0.034; q=0.136 | | | | | | |
| MRI risk score × Age | | | | | | | — | | | | | | | | | | | 0.183 (-0.070 to 0.436) p=0.156; q=0.390 | | | | | | | | | | | | — | | | | | | |
| MRI risk score × Female (vs male) | | | | | | | — | | | | | | | | | | | -0.228 (-0.738 to 0.281) p=0.380; q=0.634 | | | | | | | | | | | | — | | | | | | |
| BrainAGE | | | | | | | — | | | | | | | | | | | — | | | | | | | | | | | | -0.155 (-0.410 to 0.100) p=0.234 | | | | | | |
| MRI risk score × BrainAGE | | | | | | | — | | | | | | | | | | | — | | | | | | | | | | | | 0.168 (-0.093 to 0.428) p=0.207; q=0.413 | | | | | | |
| **Fit and nested comparisons** | | | | | | | | | | | | | | | | | | | | | | | | | | | | | | | | | | | | |
| **Model** | | | **N/events** | | | | | | | **AIC** | | | | | | | **Rank** | | | | | | | **Interaction family** | | | | | | | | | | | | |
| M1 | | | 658/84 | | | | | | | 483.260 | | | | | | | 10 | | | | | | | 3 | | | | | | | | | | | | |
| M2 | | | 658/84 | | | | | | | 485.176 | | | | | | | 12 | | | | | | | 3 | | | | | | | | | | | | |
| M2b | | | 658/84 | | | | | | | 486.235 | | | | | | | 14 | | | | | | | 5 | | | | | | | | | | | | |
| M3 | | | 658/84 | | | | | | | 486.767 | | | | | | | 14 | | | | | | | 4 | | | | | | | | | | | | |
| **Comparison** | | | | | | | | | | | **df** | | | **Likelihood-ratio statistic** | | | | | | | | | | | | | | | | | | **p** | | | | |
| M1 versus M2 | | | | | | | | | | | 2 | | | 2.083 | | | | | | | | | | | | | | | | | | 0.353 | | | | |
| M2 versus M2b | | | | | | | | | | | 2 | | | 2.941 | | | | | | | | | | | | | | | | | | 0.230 | | | | |
| M2 versus M3 | | | | | | | | | | | 2 | | | 2.409 | | | | | | | | | | | | | | | | | | 0.300 | | | | |
| **MRI score × SIPS-N interaction by cohort** | | | | | | | | | | | | | | | | | | | | | | | | | | | | | | | | | | | | |
| **Cohort (N/transitions)** | | | | | **Interaction β (SE)** | | | | | | | **P** | | | | **AUC below median SIPS-N (transitions)** | | | | | | | | | | | | | **AUC at or above median (transitions)** | | | | | | | |
| NAPLS-3 (449/50) | | | | | 0.29 (0.14) | | | | | | | .03 | | | | 0.44 (17) | | | | | | | | | | | | | 0.58 (33) | | | | | | | |
| PRONIA (163/23) | | | | | 0.13 (0.28) | | | | | | | .65 | | | | 0.72 (12) | | | | | | | | | | | | | 0.75 (11) | | | | | | | |
| ZInEP (58/11) | | | | | 0.51 (0.39) | | | | | | | .19 | | | | 0.34 (4) | | | | | | | | | | | | | 0.72 (7) | | | | | | | |
| **Baseline clinical profile by cohort and outcome** | | | | | | | | | | | | | | | | | | | | | | | | | | | | | | | | | | | | |
| **Cohort, outcome (N)** | | | | | **Age, y** | | | | | **Female, No. (%)** | | | | | **SIPS-P** | | | | | | **SIPS-N** | | | | | | **COGDIS** | | | | | | | | | **MRI score** |
| PRONIA, transition (23) | | | | | 24.1 (5.9) | | | | | 9 (39) | | | | | 10.9 (3.8) | | | | | | 11.9 (9.8) | | | | | | 9.5 (10.7) | | | | | | | | | 0.26 (0.31) |
| PRONIA, no transition (139) | | | | | 23.8 (5.3) | | | | | 71 (51) | | | | | 7.6 (4.4) | | | | | | 9.6 (6.4) | | | | | | 8.8 (6.7) | | | | | | | | | −0.02 (0.36) |
| PRONIA, standardized difference | | | | | 0.06 | | | | |  | | | | | 0.76 | | | | | | 0.33 | | | | | | 0.10 | | | | | | | | | 0.80 |
| NAPLS-3, transition (50) | | | | | 19.3 (4.3) | | | | | 24 (48) | | | | | 14.2 (3.4) | | | | | | 15.1 (6.2) | | | | | | 13.5 (8.2) | | | | | | | | | 0.01 (0.36) |
| NAPLS-3, no transition (399) | | | | | 18.5 (3.9) | | | | | 200 (50) | | | | | 13.0 (3.3) | | | | | | 12.1 (6.1) | | | | | | 10.5 (8.6) | | | | | | | | | −0.02 (0.38) |
| NAPLS-3, standardized difference | | | | | 0.21 | | | | |  | | | | | 0.36 | | | | | | 0.49 | | | | | | 0.35 | | | | | | | | | 0.09 |
| ZInEP, transition (11) | | | | | 20.4 (4.7) | | | | | 5 (45) | | | | | 9.5 (7.8) | | | | | | 13.7 (6.9) | | | | | | 10.2 (11.7) | | | | | | | | | 0.10 (0.43) |
| ZInEP, no transition (47) | | | | | 21.0 (5.9) | | | | | 16 (34) | | | | | 8.0 (4.7) | | | | | | 10.9 (6.0) | | | | | | 13.2 (10.1) | | | | | | | | | −0.03 (0.43) |
| ZInEP, standardized difference | | | | | −0.11 | | | | |  | | | | | 0.29 | | | | | | 0.46 | | | | | | −0.29 | | | | | | | | | 0.32 |
| **Premorbid adjustment and MRI score discrimination in PRONIA** | | | | | | | | | | | | | | | | | | | | | | | | | | | | | | | | | | | | |
| **Measure** | | | | | | | | | | | | | | | | | | | | **Estimate** | | | | | | | | | | | | | | | | |
| Premorbid adjustment, transition (23), mean (SD) | | | | | | | | | | | | | | | | | | | | 1.68 (0.83) | | | | | | | | | | | | | | | | |
| Premorbid adjustment, no transition (132), mean (SD) | | | | | | | | | | | | | | | | | | | | 1.65 (0.78); *P* = .73 | | | | | | | | | | | | | | | | |
| AUC at or below median adjustment score (78; 11 transitions) | | | | | | | | | | | | | | | | | | | | 0.70 (0.57 to 0.83) | | | | | | | | | | | | | | | | |
| AUC above median adjustment score (77; 12 transitions) | | | | | | | | | | | | | | | | | | | | 0.80 (0.66 to 0.94) | | | | | | | | | | | | | | | | |
| MRI score × premorbid adjustment, β (SE) | | | | | | | | | | | | | | | | | | | | 0.16 (0.29); *P* = .57 | | | | | | | | | | | | | | | | |
| **MRI score discrimination in PRONIA by documented timing of transition** | | | | | | | | | | | | | | | | | | | | | | | | | | | | | | | | | | | | |
| **Transition definition** | | | | | | | | | | | | | | | | | **Participants (transitions)** | | | | | | | | | | | **AUC (95% CI)** | | | | | | | | |
| All transitions | | | | | | | | | | | | | | | | | 164 (23) | | | | | | | | | | | 0.73 (0.64 to 0.83) | | | | | | | | |
| Onset documented at the 9- or 18-month assessment; other transitions counted as nontransitioning | | | | | | | | | | | | | | | | | 164 (13) | | | | | | | | | | | 0.65 (0.52 to 0.79) | | | | | | | | |
| Onset documented at the 9- or 18-month assessment; other transitions excluded | | | | | | | | | | | | | | | | | 154 (13) | | | | | | | | | | | 0.68 (0.54 to 0.81) | | | | | | | | |
| Later or undocumented transitions vs nontransitions | | | | | | | | | | | | | | | | | 151 (10) | | | | | | | | | | | 0.81 (0.72 to 0.90) | | | | | | | | |
| NAPLS-3, all transitions (reference) | | | | | | | | | | | | | | | | | 458 (51) | | | | | | | | | | | 0.53 (0.45 to 0.61) | | | | | | | | |
| **Separation of participants with and without transition by cohort** | | | | | | | | | | | | | | | | | | | | | | | | | | | | | | | | | | | | |
| **Variable** | **PRONIA (329/26)** | | | | | **NAPLS-3 (458/51)** | | | | | | **ZInEP (157/23)** | | | | | | | **FePsy (37/16)** | | | | | | **MUC-FRUE (33/16)** | | | | | | | | | | **External summary (685/106)** | |
| MRI score | 0.90 (0.62 to 1.21) | | | | | 0.10 (−0.18 to 0.39) | | | | | | 0.51 (0.06 to 1.01) | | | | | | | 0.54 (−0.13 to 1.45) | | | | | | −0.04 (−0.72 to 0.69) | | | | | | | | | | 0.21 (0.00 to 0.45) | |
| Comp 1 | 0.71 (0.39 to 1.08) | | | | | 0.05 (−0.25 to 0.39) | | | | | | 0.27 (−0.18 to 0.71) | | | | | | | 0.15 (−0.51 to 0.87) | | | | | | 0.43 (−0.18 to 1.28) | | | | | | | | | | 0.12 (−0.12 to 0.36) | |
| Comp 2 | 0.91 (0.60 to 1.27) | | | | | 0.19 (−0.11 to 0.51) | | | | | | 0.18 (−0.24 to 0.67) | | | | | | | 0.54 (−0.12 to 1.30) | | | | | | −0.01 (−0.72 to 0.68) | | | | | | | | | | 0.20 (−0.03 to 0.45) | |
| Comp 3 | 0.61 (0.28 to 0.98) | | | | | −0.09 (−0.37 to 0.17) | | | | | | 0.24 (−0.20 to 0.67) | | | | | | | 0.68 (0.02 to 1.59) | | | | | | −0.36 (−1.09 to 0.32) | | | | | | | | | | 0.01 (−0.20 to 0.23) | |
| Comp 4 | 0.56 (0.16 to 0.97) | | | | | −0.05 (−0.35 to 0.25) | | | | | | 0.15 (−0.31 to 0.68) | | | | | | | −0.40 (−1.10 to 0.24) | | | | | | −0.57 (−1.31 to 0.07) | | | | | | | | | | −0.05 (−0.28 to 0.19) | |
| **Loss of separation relative to PRONIA by component** | | | | | | | | | | | | | | | | | | | | | | | | | | | | | | | | | | | | |
| **Variable** | | **Loss, NAPLS-3** | | | | | | **Relative loss, NAPLS-3** | | | | | **Loss, external summary** | | | | | | | | | | **Relative loss, external summary** | | | | | | | | | | **q (NAPLS-3 / external)** | | | |
| MRI score | | 0.80 (0.41 to 1.24) | | | | | | NA | | | | | 0.69 (0.33 to 1.08) | | | | | | | | | | NA | | | | | | | | | | NA | | | |
| Comp 1 | | 0.66 (0.19 to 1.13) | | | | | | −0.01 (−0.39 to 0.38) | | | | | 0.59 (0.18 to 1.02) | | | | | | | | | | −0.04 (−0.38 to 0.31) | | | | | | | | | | .93 / .93 | | | |
| Comp 2 | | 0.72 (0.24 to 1.18) | | | | | | 0.04 (−0.37 to 0.48) | | | | | 0.71 (0.30 to 1.14) | | | | | | | | | | 0.08 (−0.29 to 0.47) | | | | | | | | | | .93 / .93 | | | |
| Comp 3 | | 0.70 (0.26 to 1.15) | | | | | | 0.03 (−0.37 to 0.43) | | | | | 0.59 (0.21 to 1.03) | | | | | | | | | | −0.03 (−0.40 to 0.35) | | | | | | | | | | .93 / .93 | | | |
| Comp 4 | | 0.61 (0.12 to 1.12) | | | | | | −0.06 (−0.51 to 0.40) | | | | | 0.61 (0.16 to 1.07) | | | | | | | | | | −0.02 (−0.43 to 0.40) | | | | | | | | | | .93 / .93 | | | |

Simple slopes are unadjusted descriptive contrasts of the same fitted model; different slope p-values are not themselves an interaction test.

Entries are standardized log-odds coefficients (95% Wald CI), followed by unadjusted p and, for score interactions, within-model BH q. NAPLS-3 is the cohort reference; male is the recorded-sex reference. Model 3 extends Model 2, not Model 2b.

All participants with SIPS-N ratings; in PRONIA, 162 participants with CHR-P and 1 participant with recent-onset depression (no transition). Logistic regression within each cohort with standardized MRI score, standardized SIPS-N and their interaction. Medians are cohort specific. Unadjusted Wald *P* values; descriptive check of direction.

Participants with SIPS ratings; PRONIA participants with CHR-P only. Values are mean (SD) unless otherwise indicated. Standardized difference, transition minus no transition divided by the pooled SD weighted by degrees of freedom. COGDIS was missing for 11 NAPLS-3 participants without transition. MRI score, decision score of the fixed pipeline.

PRONIA participants with CHR-P and complete Premorbid Adjustment Scale items for childhood and early adolescence (155 participants; 23 transitions). Adjustment score, mean item score (range, 0-6; higher scores indicate poorer adjustment); median, 1.56. *P* for the group difference from a Wilcoxon rank sum test. AUC with DeLong 95% CI. Interaction from a logistic regression with standardized MRI score, standardized adjustment score and their product.

PRONIA participants with CHR-P. Onset dates came from the transition criteria recorded at the 9- and 18-month assessments; documented onsets occurred a median of 7.0 months after baseline (maximum, 20.1). Later or undocumented transitions had no onset recorded at either assessment. AUC with DeLong 95% CI.

Values are standardized mean differences (transition minus nontransition, pooled SD) with bootstrap 95% CIs; column headings give participants/transitions. The external summary is the participant-weighted mean of the 4 external cohorts. Component expression is the dot product of the smoothed gray matter map and the component voxel weights (eMethods 13). Spearman correlations between components ranged from −0.07 to 0.17 in PRONIA and from −0.01 to 0.22 in NAPLS-3.

Loss is the separation in PRONIA minus that in the external cohort or summary. Relative loss is the loss of a component minus the mean loss of the 4 components; positive values indicate a larger loss than average. Bootstrap 95% CIs; q values are Benjamini-Hochberg adjusted across the 8 relative-loss contrasts. NA, not applicable.

eTable 7. Functioning-complete regressions and GAM sensitivity analyses. The regression sections compare logistic regressions in 254 participants (38 transitions); the GAM sections concern the 658 symptom-complete participants (84 transitions).

| **Clinical regression including functioning** | | | | | | | | | | | | | | | | | | | | | | | | | | | | |
| --- | --- | --- | --- | --- | --- | --- | --- | --- | --- | --- | --- | --- | --- | --- | --- | --- | --- | --- | --- | --- | --- | --- | --- | --- | --- | --- | --- | --- |
| **Term** | | | **β (95% CI)** | | | | | | | | | | | | | | **SE** | | | | **p** | | | | | | **Interaction q** | |
| Intercept | | | -3.180 (-4.462 to -1.898) | | | | | | | | | | | | | | 0.654 | | | | <0.001 | | | | | | — | |
| MRI risk score | | | 0.707 (0.259 to 1.155) | | | | | | | | | | | | | | 0.229 | | | | 0.002 | | | | | | — | |
| COGDIS | | | -0.551 (-1.050 to -0.052) | | | | | | | | | | | | | | 0.255 | | | | 0.030 | | | | | | — | |
| SIPS-P | | | 0.699 (0.226 to 1.172) | | | | | | | | | | | | | | 0.241 | | | | 0.004 | | | | | | — | |
| SIPS-N | | | 0.264 (-0.264 to 0.791) | | | | | | | | | | | | | | 0.269 | | | | 0.327 | | | | | | — | |
| Functioning | | | 0.161 (-0.341 to 0.663) | | | | | | | | | | | | | | 0.256 | | | | 0.530 | | | | | | — | |
| PRONIA (vs NAPLS-3) | | | 1.102 (-0.227 to 2.431) | | | | | | | | | | | | | | 0.678 | | | | 0.104 | | | | | | — | |
| ZInEP (vs NAPLS-3) | | | 1.466 (0.040 to 2.891) | | | | | | | | | | | | | | 0.727 | | | | 0.044 | | | | | | — | |
| MRI risk score × COGDIS | | | 0.324 (-0.098 to 0.746) | | | | | | | | | | | | | | 0.215 | | | | 0.133 | | | | | | 0.531 | |
| MRI risk score × SIPS-P | | | 0.004 (-0.418 to 0.425) | | | | | | | | | | | | | | 0.215 | | | | 0.987 | | | | | | 0.987 | |
| MRI risk score × SIPS-N | | | 0.040 (-0.446 to 0.526) | | | | | | | | | | | | | | 0.248 | | | | 0.872 | | | | | | 0.987 | |
| MRI risk score × Functioning | | | -0.203 (-0.701 to 0.295) | | | | | | | | | | | | | | 0.254 | | | | 0.425 | | | | | | 0.851 | |
| *Matched clinical regression without functioning* | | | | | | | | | | | | | | | | | | | | | | | | | | | | |
| **Term** | | | **β (95% CI)** | | | | | | | | | | | | | | **SE** | | | | **p** | | | | | | **Interaction q** | |
| Intercept | | | -3.045 (-4.251 to -1.839) | | | | | | | | | | | | | | 0.615 | | | | <0.001 | | | | | | — | |
| MRI risk score | | | 0.690 (0.246 to 1.134) | | | | | | | | | | | | | | 0.226 | | | | 0.002 | | | | | | — | |
| COGDIS | | | -0.560 (-1.062 to -0.059) | | | | | | | | | | | | | | 0.256 | | | | 0.028 | | | | | | — | |
| SIPS-P | | | 0.646 (0.202 to 1.090) | | | | | | | | | | | | | | 0.227 | | | | 0.004 | | | | | | — | |
| SIPS-N | | | 0.165 (-0.282 to 0.613) | | | | | | | | | | | | | | 0.228 | | | | 0.469 | | | | | | — | |
| PRONIA (vs NAPLS-3) | | | 0.968 (-0.282 to 2.219) | | | | | | | | | | | | | | 0.638 | | | | 0.129 | | | | | | — | |
| ZInEP (vs NAPLS-3) | | | 1.364 (-0.005 to 2.732) | | | | | | | | | | | | | | 0.698 | | | | 0.051 | | | | | | — | |
| MRI risk score × COGDIS | | | 0.330 (-0.095 to 0.755) | | | | | | | | | | | | | | 0.217 | | | | 0.129 | | | | | | 0.386 | |
| MRI risk score × SIPS-P | | | 0.045 (-0.369 to 0.460) | | | | | | | | | | | | | | 0.211 | | | | 0.830 | | | | | | 0.830 | |
| MRI risk score × SIPS-N | | | 0.159 (-0.242 to 0.560) | | | | | | | | | | | | | | 0.205 | | | | 0.437 | | | | | | 0.656 | |
| **Functioning-regression extensions** | | | | | | | | | | | | | | | | | | | | | | | | | | | | |
| **Term** | | **F2 Demographic main** | | | | | | | | | | | **F2b Demographic interactions** | | | | | | | | | | **F3 BrainAGE** | | | | | |
| Intercept | | -2.996 (-4.325 to -1.667) p<0.001 | | | | | | | | | | | -3.109 (-4.464 to -1.754) p<0.001 | | | | | | | | | | -3.149 (-4.523 to -1.775) p<0.001 | | | | | |
| MRI risk score | | 0.715 (0.262 to 1.169) p=0.002 | | | | | | | | | | | 0.920 (0.349 to 1.491) p=0.002 | | | | | | | | | | 0.726 (0.258 to 1.194) p=0.002 | | | | | |
| COGDIS | | -0.531 (-1.030 to -0.032) p=0.037 | | | | | | | | | | | -0.566 (-1.076 to -0.055) p=0.030 | | | | | | | | | | -0.577 (-1.074 to -0.081) p=0.023 | | | | | |
| SIPS-P | | 0.693 (0.221 to 1.166) p=0.004 | | | | | | | | | | | 0.745 (0.256 to 1.234) p=0.003 | | | | | | | | | | 0.712 (0.234 to 1.190) p=0.004 | | | | | |
| SIPS-N | | 0.264 (-0.268 to 0.795) p=0.331 | | | | | | | | | | | 0.286 (-0.246 to 0.817) p=0.292 | | | | | | | | | | 0.279 (-0.253 to 0.810) p=0.304 | | | | | |
| Functioning | | 0.183 (-0.324 to 0.690) p=0.480 | | | | | | | | | | | 0.183 (-0.341 to 0.707) p=0.494 | | | | | | | | | | 0.191 (-0.321 to 0.702) p=0.466 | | | | | |
| Age | | 0.071 (-0.345 to 0.486) p=0.740 | | | | | | | | | | | 0.059 (-0.429 to 0.546) p=0.814 | | | | | | | | | | 0.052 (-0.371 to 0.475) p=0.808 | | | | | |
| Female (vs male) | | -0.336 (-1.138 to 0.465) p=0.411 | | | | | | | | | | | -0.115 (-0.970 to 0.741) p=0.793 | | | | | | | | | | -0.367 (-1.181 to 0.447) p=0.377 | | | | | |
| PRONIA (vs NAPLS-3) | | 1.077 (-0.310 to 2.463) p=0.128 | | | | | | | | | | | 1.113 (-0.277 to 2.502) p=0.117 | | | | | | | | | | 1.200 (-0.223 to 2.623) p=0.098 | | | | | |
| ZInEP (vs NAPLS-3) | | 1.397 (-0.040 to 2.834) p=0.057 | | | | | | | | | | | 1.393 (-0.048 to 2.835) p=0.058 | | | | | | | | | | 1.561 (0.087 to 3.035) p=0.038 | | | | | |
| MRI risk score × COGDIS | | 0.316 (-0.103 to 0.734) p=0.139; q=0.557 | | | | | | | | | | | 0.383 (-0.063 to 0.829) p=0.092; q=0.553 | | | | | | | | | | 0.327 (-0.088 to 0.742) p=0.122; q=0.306 | | | | | |
| MRI risk score × SIPS-P | | 0.038 (-0.387 to 0.463) p=0.862; q=0.862 | | | | | | | | | | | 0.024 (-0.418 to 0.466) p=0.916; q=0.978 | | | | | | | | | | 0.017 (-0.416 to 0.450) p=0.939; q=0.939 | | | | | |
| MRI risk score × SIPS-N | | 0.046 (-0.442 to 0.533) p=0.855; q=0.862 | | | | | | | | | | | 0.007 (-0.498 to 0.512) p=0.978; q=0.978 | | | | | | | | | | 0.072 (-0.413 to 0.557) p=0.771; q=0.939 | | | | | |
| MRI risk score × Functioning | | -0.204 (-0.700 to 0.292) p=0.421; q=0.841 | | | | | | | | | | | -0.209 (-0.730 to 0.313) p=0.433; q=0.865 | | | | | | | | | | -0.195 (-0.699 to 0.309) p=0.448; q=0.746 | | | | | |
| MRI risk score × Age | | — | | | | | | | | | | | 0.036 (-0.412 to 0.485) p=0.874; q=0.978 | | | | | | | | | | — | | | | | |
| MRI risk score × Female (vs male) | | — | | | | | | | | | | | -0.590 (-1.484 to 0.304) p=0.196; q=0.588 | | | | | | | | | | — | | | | | |
| BrainAGE | | — | | | | | | | | | | | — | | | | | | | | | | -0.305 (-0.788 to 0.178) p=0.216 | | | | | |
| MRI risk score × BrainAGE | | — | | | | | | | | | | | — | | | | | | | | | | 0.398 (-0.083 to 0.878) p=0.105; q=0.306 | | | | | |
| **Fit and nested comparisons** | | | | | | | | | | | | | | | | | | | | | | | | | | | | |
| **Model** | | **N/events** | | | | | | **AIC** | | | | | | | **Rank** | | | | | **Interaction family** | | | | | | | | |
| F1 | | 254/38 | | | | | | 207.601 | | | | | | | 12 | | | | | 4 | | | | | | | | |
| F2 | | 254/38 | | | | | | 210.699 | | | | | | | 14 | | | | | 4 | | | | | | | | |
| F2b | | 254/38 | | | | | | 212.984 | | | | | | | 16 | | | | | 6 | | | | | | | | |
| F3 | | 254/38 | | | | | | 211.723 | | | | | | | 16 | | | | | 5 | | | | | | | | |
| Matched, no functioning | | 254/38 | | | | | | 204.410 | | | | | | | 10 | | | | | 3 | | | | | | | | |
| **Comparison** | | | | | | **df** | | | | **Likelihood-ratio statistic** | | | | | | | | | | | | | | | **p** | | | |
| F1 versus F2 | | | | | | 2 | | | | 0.903 | | | | | | | | | | | | | | | 0.637 | | | |
| F2 versus F2b | | | | | | 2 | | | | 1.715 | | | | | | | | | | | | | | | 0.424 | | | |
| F2 versus F3 | | | | | | 2 | | | | 2.976 | | | | | | | | | | | | | | | 0.226 | | | |
| **GAM smooth-term tests and bounded specification comparisons** | | | | | | | | | | | | | | | | | | | | | | | | | | | | |
| *Primary: k=10, selection* | | | | | | | | | | | | | | | | | | | | | | | | | | | | |
| **Term** | | | | | | | **edf** | | | | **Reference df** | | | | | **Statistic** | | | | | | **p** | | | | | | **Tensor q** |
| Smooth: MRI risk score | | | | | | | 0.79 | | | | 9.00 | | | | | 3.642 | | | | | | 0.032 | | | | | | — |
| Smooth: COGDIS | | | | | | | 0.00 | | | | 9.00 | | | | | 0.000 | | | | | | 0.974 | | | | | | — |
| Tensor: MRI risk score × COGDIS | | | | | | | 0.00 | | | | 81.00 | | | | | 0.000 | | | | | | 0.962 | | | | | | 0.962 |
| Smooth: SIPS-P | | | | | | | 0.92 | | | | 9.00 | | | | | 10.565 | | | | | | <0.001 | | | | | | — |
| Tensor: MRI risk score × SIPS-P | | | | | | | 0.00 | | | | 81.00 | | | | | 0.000 | | | | | | 0.851 | | | | | | 0.962 |
| Smooth: SIPS-N | | | | | | | 0.78 | | | | 9.00 | | | | | 3.363 | | | | | | 0.034 | | | | | | — |
| Tensor: MRI risk score × SIPS-N | | | | | | | 1.41 | | | | 59.00 | | | | | 5.730 | | | | | | 0.016 | | | | | | 0.047 |
| *Diagnostic: k=6, selection* | | | | | | | | | | | | | | | | | | | | | | | | | | | | |
| **Term** | | | | | | | **edf** | | | | **Reference df** | | | | | **Statistic** | | | | | | **p** | | | | | | **Tensor q** |
| Smooth: MRI risk score | | | | | | | 0.79 | | | | 5.00 | | | | | 3.654 | | | | | | 0.031 | | | | | | — |
| Smooth: COGDIS | | | | | | | 0.00 | | | | 5.00 | | | | | 0.000 | | | | | | 0.967 | | | | | | — |
| Tensor: MRI risk score × COGDIS | | | | | | | 0.00 | | | | 25.00 | | | | | 0.000 | | | | | | 0.904 | | | | | | 0.904 |
| Smooth: SIPS-P | | | | | | | 1.78 | | | | 5.00 | | | | | 11.420 | | | | | | <0.001 | | | | | | — |
| Tensor: MRI risk score × SIPS-P | | | | | | | 0.00 | | | | 25.00 | | | | | 0.000 | | | | | | 0.858 | | | | | | 0.904 |
| Smooth: SIPS-N | | | | | | | 0.78 | | | | 5.00 | | | | | 3.446 | | | | | | 0.032 | | | | | | — |
| Tensor: MRI risk score × SIPS-N | | | | | | | 1.32 | | | | 25.00 | | | | | 5.603 | | | | | | 0.016 | | | | | | 0.047 |
| *Diagnostic: k=10, no extra selection* | | | | | | | | | | | | | | | | | | | | | | | | | | | | |
| **Term** | | | | | | | **edf** | | | | **Reference df** | | | | | **Statistic** | | | | | | **p** | | | | | | **Tensor q** |
| Smooth: MRI risk score | | | | | | | 1.00 | | | | 1.00 | | | | | 4.141 | | | | | | 0.042 | | | | | | — |
| Smooth: COGDIS | | | | | | | 1.00 | | | | 1.00 | | | | | 0.202 | | | | | | 0.653 | | | | | | — |
| Tensor: MRI risk score × COGDIS | | | | | | | 1.00 | | | | 1.00 | | | | | 0.073 | | | | | | 0.787 | | | | | | 0.787 |
| Smooth: SIPS-P | | | | | | | 1.00 | | | | 1.00 | | | | | 11.234 | | | | | | <0.001 | | | | | | — |
| Tensor: MRI risk score × SIPS-P | | | | | | | 1.00 | | | | 1.00 | | | | | 0.187 | | | | | | 0.666 | | | | | | 0.787 |
| Smooth: SIPS-N | | | | | | | 1.00 | | | | 1.00 | | | | | 3.077 | | | | | | 0.079 | | | | | | — |
| Tensor: MRI risk score × SIPS-N | | | | | | | 1.73 | | | | 2.18 | | | | | 4.840 | | | | | | 0.101 | | | | | | 0.302 |
| *Diagnostic: cubic regression, k=10, selection* | | | | | | | | | | | | | | | | | | | | | | | | | | | | |
| **Term** | | | | | | | **edf** | | | | **Reference df** | | | | | **Statistic** | | | | | | **p** | | | | | | **Tensor q** |
| Smooth: MRI risk score | | | | | | | 0.79 | | | | 8.00 | | | | | 3.642 | | | | | | 0.032 | | | | | | — |
| Smooth: COGDIS | | | | | | | 0.00 | | | | 9.00 | | | | | 0.000 | | | | | | 0.973 | | | | | | — |
| Tensor: MRI risk score × COGDIS | | | | | | | 0.00 | | | | 81.00 | | | | | 0.000 | | | | | | 0.915 | | | | | | 0.915 |
| Smooth: SIPS-P | | | | | | | 0.92 | | | | 9.00 | | | | | 10.565 | | | | | | <0.001 | | | | | | — |
| Tensor: MRI risk score × SIPS-P | | | | | | | 0.00 | | | | 81.00 | | | | | 0.000 | | | | | | 0.854 | | | | | | 0.915 |
| Smooth: SIPS-N | | | | | | | 0.78 | | | | 9.00 | | | | | 3.363 | | | | | | 0.034 | | | | | | — |
| Tensor: MRI risk score × SIPS-N | | | | | | | 1.41 | | | | 73.00 | | | | | 5.730 | | | | | | 0.016 | | | | | | 0.047 |
| **GAM diagnostics** | | | | | | | | | | | | | | | | | | | | | | | | | | | | |
| **Specification** | | | | | **Converged** | | | | **Rank / coefficients** | | | | | | | | | **Log likelihood** | | | | | | | **Deviance explained** | | | |
| Primary: k=10, selection | | | | | True | | | | 282/282 | | | | | | | | | -231.275 | | | | | | | 0.080 | | | |
| Diagnostic: k=6, selection | | | | | True | | | | 98/98 | | | | | | | | | -230.639 | | | | | | | 0.082 | | | |
| Diagnostic: k=10, no extra selection | | | | | True | | | | 282/282 | | | | | | | | | -230.648 | | | | | | | 0.082 | | | |
| Diagnostic: cubic regression, k=10, selection | | | | | True | | | | 282/282 | | | | | | | | | -231.275 | | | | | | | 0.080 | | | |
| *Primary-model k-index diagnostics and row-order sensitivity* | | | | | | | | | | | | | | | | | | | | | | | | | | | | |
| **Term** | | | | **Original order** | | | | | | | | | | **Reversed order** | | | | | | | | | | **Shuffled order** | | | | |
| Smooth: MRI risk score | | | | 0.985; p=0.812 | | | | | | | | | | 0.985; p=0.800 | | | | | | | | | | 0.985; p=0.787 | | | | |
| Smooth: COGDIS | | | | 0.577; 0/400 exceedances | | | | | | | | | | 0.557; 0/400 exceedances | | | | | | | | | | 0.947; p=0.415 | | | | |
| Tensor: MRI risk score × COGDIS | | | | 0.973; p=0.750 | | | | | | | | | | 0.973; p=0.695 | | | | | | | | | | 0.973; p=0.725 | | | | |
| Smooth: SIPS-P | | | | 0.479; 0/400 exceedances | | | | | | | | | | 0.487; 0/400 exceedances | | | | | | | | | | 0.944; p=0.412 | | | | |
| Tensor: MRI risk score × SIPS-P | | | | 0.978; p=0.777 | | | | | | | | | | 0.978; p=0.787 | | | | | | | | | | 0.978; p=0.790 | | | | |
| Smooth: SIPS-N | | | | 0.624; 0/400 exceedances | | | | | | | | | | 0.622; 0/400 exceedances | | | | | | | | | | 0.922; p=0.225 | | | | |
| Tensor: MRI risk score × SIPS-N | | | | 0.926; p=0.245 | | | | | | | | | | 0.926; p=0.247 | | | | | | | | | | 0.926; p=0.207 | | | | |
| *Concurvity* | | | | | | | | | | | | | | | | | | | | | | | | | | | | |
| **Term** | | | | | | | | | | | | **Worst** | | | | | | | **Observed** | | | | | | | **Estimate** | | |
| Parametric | | | | | | | | | | | | 0.999 | | | | | | | 0.999 | | | | | | | 0.999 | | |
| Smooth: MRI risk score | | | | | | | | | | | | 1.000 | | | | | | | 0.987 | | | | | | | 0.983 | | |
| Smooth: COGDIS | | | | | | | | | | | | 1.000 | | | | | | | 0.719 | | | | | | | 0.819 | | |
| Tensor: MRI risk score × COGDIS | | | | | | | | | | | | 1.000 | | | | | | | 0.478 | | | | | | | 0.401 | | |
| Smooth: SIPS-P | | | | | | | | | | | | 1.000 | | | | | | | 0.722 | | | | | | | 0.709 | | |
| Tensor: MRI risk score × SIPS-P | | | | | | | | | | | | 1.000 | | | | | | | 0.377 | | | | | | | 0.379 | | |
| Smooth: SIPS-N | | | | | | | | | | | | 1.000 | | | | | | | 0.775 | | | | | | | 0.760 | | |
| Tensor: MRI risk score × SIPS-N | | | | | | | | | | | | 1.000 | | | | | | | 0.671 | | | | | | | 0.423 | | |
| *Bernoulli simulation diagnostics* | | | | | | | | | | | | | | | | | | | | | | | | | | | | |
| **Summary** | **Observed** | | | | **Simulation mean** | | | | | | | | | **Simulation 2.5%-97.5%** | | | | | | | | | | | | | **Simulation p** | |
| events | 84.000 | | | | 83.975 | | | | | | | | | 69.000 to 99.000 | | | | | | | | | | | | | 1.000 | |
| Brier | 0.103 | | | | 0.104 | | | | | | | | | 0.087 to 0.121 | | | | | | | | | | | | | 0.886 | |
| Pearson | 623.487 | | | | 657.806 | | | | | | | | | 525.873 to 794.614 | | | | | | | | | | | | | 0.604 | |

These two fits use the same participants and shared-variable standardization. NAPLS-3, the cohort reference, contributes 37 participants (4 transitions), rated with the SOFAS rather than the GAF; cohort terms adjust for cohort differences and are not interpreted. The interaction families contain four and three score interactions, respectively.

Entries are standardized log-odds coefficients (95% Wald CI), followed by unadjusted p and, for score interactions, within-model BH q. NAPLS-3 is the cohort reference; male is the recorded-sex reference. F3 extends F2, not F2b.

All fits use 658 participants, 84 transitions, NAPLS-3 reference, and the same analysis-set scaling. p-values are approximate smooth-term tests; q-values correct the three tensor interactions within each fit, not the search across fits or linear/GAM forms.

k-index p-values use 400 resamples. For zero exceedances, the count 0/400 is shown; the Monte Carlo resolution is 1/400=0.0025. This count is neither an exact p=0 nor a confidence bound on the underlying p-value. These neighbor-based diagnostics are sensitive to the ordering of tied symptom scores. Row-order likelihood differences were negligible; the change in diagnostic values is not independent evidence that the fit is adequate.

1000 simulated Bernoulli outcomes conditional on the fitted probabilities, without refitting. These summaries do not establish transportability, correct the cross-form search, or remove concurvity concerns.

eTable 8. Pairwise associations among age, recorded sex, BrainAGE, COGDIS, SIPS-P, SIPS-N, and functioning. Pearson correlations are the primary descriptive summaries; Spearman correlations are a separately corrected sensitivity analysis. N is pairwise complete. Recorded sex is coded male=1, female=2.

| **Pearson correlations** | | | | | |
| --- | --- | --- | --- | --- | --- |
| **Variable 1** | **Variable 2** | **N** | **r** | **p** | **BH q** |
| Age | Recorded sex | 1014 | -0.036 | 0.251 | 0.405 |
| Age | COGDIS | 663 | 0.032 | 0.415 | 0.544 |
| Age | SIPS-P | 676 | -0.275 | <0.001 | <0.001 |
| Age | SIPS-N | 670 | -0.058 | 0.132 | 0.230 |
| Age | Functioning | 422 | -0.043 | 0.381 | 0.533 |
| Age | BrainAGE | 1014 | -0.164 | <0.001 | <0.001 |
| Recorded sex | COGDIS | 663 | 0.036 | 0.348 | 0.523 |
| Recorded sex | SIPS-P | 676 | 0.017 | 0.664 | 0.821 |
| Recorded sex | SIPS-N | 670 | 0.059 | 0.125 | 0.230 |
| Recorded sex | Functioning | 422 | 0.132 | 0.006 | 0.015 |
| Recorded sex | BrainAGE | 1014 | -0.006 | 0.855 | 0.919 |
| COGDIS | SIPS-P | 663 | 0.226 | <0.001 | <0.001 |
| COGDIS | SIPS-N | 658 | 0.400 | <0.001 | <0.001 |
| COGDIS | Functioning | 257 | -0.242 | <0.001 | <0.001 |
| COGDIS | BrainAGE | 663 | 0.002 | 0.966 | 0.966 |
| SIPS-P | SIPS-N | 670 | 0.253 | <0.001 | <0.001 |
| SIPS-P | Functioning | 259 | -0.190 | 0.002 | 0.006 |
| SIPS-P | BrainAGE | 676 | -0.007 | 0.855 | 0.919 |
| SIPS-N | Functioning | 256 | -0.554 | <0.001 | <0.001 |
| SIPS-N | BrainAGE | 670 | 0.006 | 0.875 | 0.919 |
| Functioning | BrainAGE | 422 | -0.106 | 0.029 | 0.061 |
| **Spearman correlations** | | | | | |
| **Variable 1** | **Variable 2** | **N** | **ρ** | **p** | **BH q** |
| Age | Recorded sex | 1014 | -0.033 | 0.300 | 0.421 |
| Age | COGDIS | 663 | 0.029 | 0.454 | 0.596 |
| Age | SIPS-P | 676 | -0.262 | <0.001 | <0.001 |
| Age | SIPS-N | 670 | -0.054 | 0.161 | 0.261 |
| Age | Functioning | 422 | -0.074 | 0.131 | 0.230 |
| Age | BrainAGE | 1014 | -0.162 | <0.001 | <0.001 |
| Recorded sex | COGDIS | 663 | 0.041 | 0.294 | 0.421 |
| Recorded sex | SIPS-P | 676 | 0.020 | 0.599 | 0.699 |
| Recorded sex | SIPS-N | 670 | 0.061 | 0.116 | 0.221 |
| Recorded sex | Functioning | 422 | 0.136 | 0.005 | 0.013 |
| Recorded sex | BrainAGE | 1014 | -0.017 | 0.584 | 0.699 |
| COGDIS | SIPS-P | 663 | 0.243 | <0.001 | <0.001 |
| COGDIS | SIPS-N | 658 | 0.397 | <0.001 | <0.001 |
| COGDIS | Functioning | 257 | -0.225 | <0.001 | <0.001 |
| COGDIS | BrainAGE | 663 | 0.002 | 0.953 | 0.953 |
| SIPS-P | SIPS-N | 670 | 0.268 | <0.001 | <0.001 |
| SIPS-P | Functioning | 259 | -0.171 | 0.006 | 0.013 |
| SIPS-P | BrainAGE | 676 | 0.003 | 0.931 | 0.953 |
| SIPS-N | Functioning | 256 | -0.535 | <0.001 | <0.001 |
| SIPS-N | BrainAGE | 670 | -0.003 | 0.938 | 0.953 |
| Functioning | BrainAGE | 422 | -0.093 | 0.057 | 0.120 |

BH correction is across 21 comparisons for this method, not across both correlation methods together.

eTable 9. MRI-score associations with the seven moderators. Pearson correlations are the primary descriptive summaries; Spearman correlations are a separately corrected sensitivity analysis. N is pairwise complete. Recorded sex is coded male=1, female=2.

| **Pearson correlations** | | | | | |
| --- | --- | --- | --- | --- | --- |
| **Variable 1** | **Variable 2** | **N** | **r** | **p** | **BH q** |
| MRI risk score | Age | 1014 | 0.024 | 0.439 | 0.615 |
| MRI risk score | Recorded sex | 1014 | 0.068 | 0.030 | 0.105 |
| MRI risk score | BrainAGE | 1014 | -0.070 | 0.026 | 0.105 |
| MRI risk score | COGDIS | 663 | -0.016 | 0.675 | 0.675 |
| MRI risk score | SIPS-P | 676 | -0.016 | 0.673 | 0.675 |
| MRI risk score | SIPS-N | 670 | 0.045 | 0.241 | 0.422 |
| MRI risk score | Functioning | 422 | -0.088 | 0.072 | 0.167 |
| **Spearman correlations** | | | | | |
| **Variable 1** | **Variable 2** | **N** | **ρ** | **p** | **BH q** |
| MRI risk score | Age | 1014 | 0.027 | 0.391 | 0.683 |
| MRI risk score | Recorded sex | 1014 | 0.069 | 0.027 | 0.140 |
| MRI risk score | BrainAGE | 1014 | -0.055 | 0.082 | 0.192 |
| MRI risk score | COGDIS | 663 | -0.027 | 0.490 | 0.686 |
| MRI risk score | SIPS-P | 676 | -0.003 | 0.933 | 0.933 |
| MRI risk score | SIPS-N | 670 | 0.021 | 0.593 | 0.692 |
| MRI risk score | Functioning | 422 | -0.100 | 0.040 | 0.140 |

BH correction is across 7 comparisons for this method, not across both correlation methods together.

eTable 10. Normative brain-age training sample and clinical-cohort application diagnostics.

| **Normative training sample** | | | | | | | | | | | |
| --- | --- | --- | --- | --- | --- | --- | --- | --- | --- | --- | --- |
| **Characteristic** | | | | | | **Value** | | | | | |
| Healthy controls, N | | | | | | 1105 | | | | | |
| Sites, N | | | | | | 22 | | | | | |
| Age range, years | | | | | | 12-65 | | | | | |
| Mean (SD) age, years | | | | | | 27.70 (9.3) | | | | | |
| Female participants, % | | | | | | 46.43 | | | | | |
| Outer-fold cross-validation R² | | | | | | 0.69 | | | | | |
| **Clinical-cohort prediction error** | | | | | | | | | | | |
| **Cohort** | **N** | **Age range, y** | | | **MAE, y** | | **RMSE, y** | | **Signed error, y** | | **Predictive R²** |
| FePsy | 37 | 18.0 to 42.0 | | | 6.87 | | 9.64 | | 0.31 | | -1.38 |
| MUC-FRUE | 33 | 17.7 to 36.8 | | | 6.41 | | 7.88 | | -0.57 | | -1.49 |
| NAPLS-3 | 458 | 12.0 to 30.0 | | | 5.80 | | 7.74 | | 0.27 | | -2.95 |
| PRONIA | 329 | 15.1 to 41.4 | | | 6.33 | | 7.92 | | 1.01 | | -0.88 |
| ZInEP | 157 | 13.0 to 35.0 | | | 5.93 | | 7.71 | | 1.42 | | -0.71 |
| **Age association and calibration** | | | | | | | | | | | |
| **Cohort** | **Pearson r** | | **Spearman ρ** | **Age-calibration intercept, y** | | | | **Age-calibration slope** | | **Residual-age r** | |
| FePsy | 0.346 | | 0.364 | 11.58 | | | | 0.544 | | -0.295 | |
| MUC-FRUE | 0.345 | | 0.389 | 10.15 | | | | 0.556 | | -0.282 | |
| NAPLS-3 | 0.291 | | 0.291 | 7.87 | | | | 0.591 | | -0.206 | |
| PRONIA | 0.493 | | 0.496 | 6.96 | | | | 0.760 | | -0.176 | |
| ZInEP | 0.472 | | 0.495 | 8.81 | | | | 0.665 | | -0.260 | |

Training characteristics and healthy-control outer-cross-validation performance refer to the normative model. They do not establish prediction accuracy in the clinical cohorts.

Signed error is corrected predicted age minus chronological age (BrainAGE). Predictive R² uses the cohort mean-age benchmark and is not squared correlation. Calibration is regression of corrected predicted age on chronological age; residual-age r correlates signed error with age. Restricted age ranges and clinical/measurement differences affect these summaries. Negative R² does not alone establish biological invalidity or disease-related aging.

eTable 11. Subject-level associations with structural components. Spearman correlations use pairwise-complete observations. The component labels correspond to aligned identifiers 01, 02, 04 and 06.

| **MRI score versus component expression** | | | | | | | | | |
| --- | --- | --- | --- | --- | --- | --- | --- | --- | --- |
| **Component** | | **N** | | **ρ** | | **p** | | **BH q (4 tests)** | |
| Comp 1 | | 1014 | | 0.444 | | <0.001 | | <0.001 | |
| Comp 2 | | 1014 | | 0.444 | | <0.001 | | <0.001 | |
| Comp 3 | | 1014 | | 0.458 | | <0.001 | | <0.001 | |
| Comp 4 | | 1014 | | 0.457 | | <0.001 | | <0.001 | |
| **Continuous moderators versus component expression** | | | | | | | | | |
| **Moderator** | **Component** | | **N** | | **ρ** | | **p** | | **BH q (24 tests)** |
| Age | Comp 1 | | 1014 | | 0.071 | | 0.023 | | 0.397 |
| Age | Comp 2 | | 1014 | | -0.007 | | 0.813 | | 0.959 |
| Age | Comp 3 | | 1014 | | -0.028 | | 0.375 | | 0.926 |
| Age | Comp 4 | | 1014 | | -0.021 | | 0.514 | | 0.926 |
| BrainAGE | Comp 1 | | 1014 | | 0.025 | | 0.434 | | 0.926 |
| BrainAGE | Comp 2 | | 1014 | | -0.025 | | 0.425 | | 0.926 |
| BrainAGE | Comp 3 | | 1014 | | -0.067 | | 0.033 | | 0.397 |
| BrainAGE | Comp 4 | | 1014 | | -0.053 | | 0.093 | | 0.557 |
| COGDIS | Comp 1 | | 663 | | -0.059 | | 0.127 | | 0.611 |
| COGDIS | Comp 2 | | 663 | | -0.031 | | 0.432 | | 0.926 |
| COGDIS | Comp 3 | | 663 | | -0.050 | | 0.197 | | 0.787 |
| COGDIS | Comp 4 | | 663 | | -0.018 | | 0.651 | | 0.938 |
| SIPS-P | Comp 1 | | 676 | | -0.002 | | 0.959 | | 0.959 |
| SIPS-P | Comp 2 | | 676 | | 0.003 | | 0.936 | | 0.959 |
| SIPS-P | Comp 3 | | 676 | | -0.031 | | 0.421 | | 0.926 |
| SIPS-P | Comp 4 | | 676 | | -0.014 | | 0.722 | | 0.938 |
| SIPS-N | Comp 1 | | 670 | | -0.014 | | 0.709 | | 0.938 |
| SIPS-N | Comp 2 | | 670 | | 0.017 | | 0.667 | | 0.938 |
| SIPS-N | Comp 3 | | 670 | | -0.024 | | 0.540 | | 0.926 |
| SIPS-N | Comp 4 | | 670 | | 0.026 | | 0.494 | | 0.926 |
| Functioning | Comp 1 | | 422 | | 0.016 | | 0.743 | | 0.938 |
| Functioning | Comp 2 | | 422 | | -0.085 | | 0.082 | | 0.557 |
| Functioning | Comp 3 | | 422 | | -0.009 | | 0.860 | | 0.959 |
| Functioning | Comp 4 | | 422 | | 0.004 | | 0.934 | | 0.959 |

These continuous-moderator/component correlations use a 24-test family. The analysis does not establish independent biological validation, because the component representation contributes to the classifier.

eTable 12. Clinical availability, cohort composition and omnibus comparisons. Complete-case populations depend on all variables required by a particular model; they are not obtained by adding marginal availability counts.

| **Available clinical ratings by cohort** | | | | | | | | | | | | | | | | | |
| --- | --- | --- | --- | --- | --- | --- | --- | --- | --- | --- | --- | --- | --- | --- | --- | --- | --- |
| **Cohort** | **Total N** | | | | **COGDIS** | | | | **SIPS-P** | | | **SIPS-N** | | | | **Functioning** | |
| PRONIA | 329 | | | | 164 | | | | 165 | | | 163 | | | | 325 | |
| NAPLS-3 | 458 | | | | 441 | | | | 453 | | | 449 | | | | 39 | |
| ZInEP | 157 | | | | 58 | | | | 58 | | | 58 | | | | 58 | |
| FePsy | 37 | | | | 0 | | | | 0 | | | 0 | | | | 0 | |
| MUC-FRUE | 33 | | | | 0 | | | | 0 | | | 0 | | | | 0 | |
| **Moderation population composition** | | | | | | | | | | | | | | | | | |
| **Population** | | | | | | **Cohort** | | | | **N** | | | **Transitions** | | | | **Nontransitions** |
| Symptom-complete | | | | | | NAPLS-3 | | | | 438 | | | 50 | | | | 388 |
| Symptom-complete | | | | | | PRONIA | | | | 162 | | | 23 | | | | 139 |
| Symptom-complete | | | | | | ZInEP | | | | 58 | | | 11 | | | | 47 |
| Symptom-complete | | | | | | ALL | | | | 658 | | | 84 | | | | 574 |
| Functioning-complete / matched | | | | | | NAPLS-3 | | | | 37 | | | 4 | | | | 33 |
| Functioning-complete / matched | | | | | | PRONIA | | | | 159 | | | 23 | | | | 136 |
| Functioning-complete / matched | | | | | | ZInEP | | | | 58 | | | 11 | | | | 47 |
| Functioning-complete / matched | | | | | | ALL | | | | 254 | | | 38 | | | | 216 |
| **Continuous-variable comparisons across cohorts** | | | | | | | | | | | | | | | | | |
| **Variable** | | **N** | | **df** | | | **Kruskal-Wallis H** | | | | **p** | | | | **BH q** | | |
| Age | | 1014 | | 4 | | | 263.446 | | | | <0.001 | | | | <0.001 | | |
| MRI risk score | | 1014 | | 4 | | | 1.134 | | | | 0.889 | | | | 0.889 | | |
| COGDIS | | 663 | | 2 | | | 8.344 | | | | 0.015 | | | | 0.027 | | |
| SIPS-P | | 676 | | 2 | | | 162.094 | | | | <0.001 | | | | <0.001 | | |
| SIPS-N | | 670 | | 2 | | | 22.366 | | | | <0.001 | | | | <0.001 | | |
| Functioning | | 422 | | 2 | | | 3.557 | | | | 0.169 | | | | 0.236 | | |
| BrainAGE | | 1014 | | 4 | | | 3.348 | | | | 0.501 | | | | 0.585 | | |
| **Categorical comparisons** | | | | | | | | | | | | | | | | | |
| **Variable** | | | **N** | | | | | **Simulated Fisher p** | | | | | | **BH q (2 tests)** | | | |
| Recorded sex | | | 1014 | | | | | 0.053 | | | | | | 0.053 | | | |
| Transition | | | 1014 | | | | | <0.001 | | | | | | <0.001 | | | |

Age, outcome, MRI score, original probabilities and the four component scores were available for the 1014 participants; BrainAGE was also available for all 1014. FePsy and MUC-FRUE did not contribute to the symptom moderation regressions because the required ratings were unavailable.

BH correction covers seven continuous-variable omnibus tests. An omnibus difference does not identify which cohort pairs differ. No individual cohort-pair differences are inferred from these omnibus tests.

20,000 simulated tables per test, with deterministic random-number settings. The p-values are from simulated Fisher tests, with BH correction across the two categorical comparisons.

eTable 13. Statistical families and uncertainty methods. The table lists the test families used for multiple-testing correction, followed by confidence intervals and resampling. Adjustment is within each stated family, not across the entire analytical program. Unadjusted results are identified as such; exploratory hypotheses and diagnostic models are not independent replication.

| **Analysis / quantity** | **Test family or estimation method** | **Adjustment, scope and interpretation** |
| --- | --- | --- |
| **A. Multiple-testing correction** | | |
| Continuous cohort comparisons | 7 omnibus tests | BH across seven continuous variables; no post-hoc cohort-pair claims |
| Categorical cohort comparisons | 2 tests | BH across recorded sex and transition; Fisher simulation 20,000 tables |
| Moderator correlations (eTable 8) | 21 per method | Pearson and Spearman each have separate 21-test families |
| MRI-score/moderator correlations (eTable 9) | 7 per method | Separate seven-test Pearson and Spearman families |
| MRI-score/component correlations | 4 | BH across four components |
| Continuous-moderator/component correlations | 24 | BH across six moderators and four components |
| Primary 1D global landscapes | 6 | BH across six moderator Dmax tests |
| Primary 2D global surfaces | 6 | BH across six surface Dmax tests |
| 1D joint width/step sensitivities | 6 per setting | Four separate six-moderator families |
| Selected surface settings | 4 | BH across four settings of selected pair; no correction for surface selection |
| Symptom GLM hierarchy | M1/M2:3; M2b:5; M3:4 | Only score-interaction terms; each regression is a separate family |
| Functioning GLM hierarchy | F1/F2:4; F2b:6; F3:5 | Only score-interaction terms; matched no-functioning regression has 3 |
| GAM interactions | 3 per specification | Within-fit tensor family; does not correct searching across forms/penalties |
| Main effects, slopes, nested comparisons | Unadjusted | Descriptive/model-specific inference; not independent confirmation |
| Population/support sensitivity summaries | Descriptive | No new significance inferred from Dmax or eligible counts |
| Placement-value regressions (eTable 21) | 5 per regression; 2 in the external regressions | BH within each of 6 regressions; cohort indicators not counted; exploratory |
| Component separation losses (eTable 6) | 8 relative-loss contrasts (4 components in NAPLS-3 and in the external summary) | BH across the 8 contrasts; bootstrap CIs and P values |
| **B. Confidence intervals and resampling** | | |
| Cohort/pooled AUC | DeLong 95% CI; 5000 label permutations | Fixed-score evaluation |
| Random-effects AUC | Logit AUC/DeLong sampling variances; REML | Back-transformed summary |
| Local 1D AUC | 2000 outcome-stratified bootstrap percentile intervals | Pointwise, not simultaneous |
| Global landscapes | 5000 valid within-cohort moderator permutations | Add-one p-values |
| Pooled external calibration | 2000 participant-bootstrap intervals | Apparent update refitted in each resample |
| Source/cohort/held-out calibration | 500-replicate percentile intervals | Separately reported from the pooled external intervals |
| Logistic coefficients/slopes | Wald/model-covariance 95% intervals | Analysis-set scaling |
| GAM curves | Pointwise model-based 95% intervals | Diagnostics do not establish external prediction |
| Paired clinical-regression comparisons | 2000 paired within-cohort resamples of held-out predictions | No model refit |
| GAM diagnostic simulations | 400 k-diagnostic resamples; 1000 conditional Bernoulli simulations | Conditional diagnostic comparisons |
| Placement values, separation synthesis and component membership (eMethods 12) | 2000 bootstrap replicates within cohort and outcome; REML with Hartung-Knapp; 1000 label permutations | Percentile intervals; add-one permutation P values |

BH, Benjamini-Hochberg; CI, confidence interval; AUC, area under the receiver operating characteristic curve; GLM, generalized linear model; GAM, generalized additive model; REML, restricted maximum likelihood. Dmax is the range of eligible local AUC estimates.

eTable 14. Discrimination of alternative classifiers. All classifiers used the same images, partitions, and preprocessing (eMethods 3). Values are AUCs with DeLong 95% CIs; PRONIA AUCs are out of training under leave-one-site-out cross-validation; 328 of the 329 PRONIA participants had image data in the NeuroMiner structure.

| **Classifier** | **PRONIA (328/26)** | **NAPLS-3 (458/51)** | **ZInEP (157/23)** | **FePsy (37/16)** | **MUC-FRUE (33/16)** | **External, pooled (685/106)** | **External, random effects** |
| --- | --- | --- | --- | --- | --- | --- | --- |
| Original model (NeuroMiner) | 0.74 (0.66 to 0.82) | 0.53 (0.45 to 0.61) | 0.65 (0.52 to 0.78) | 0.69 (0.51 to 0.87) | 0.56 (0.34 to 0.77) | 0.56 (0.51 to 0.62) | 0.59 (0.46 to 0.70) |
| Linear reference (PCA, linear SVM) | 0.65 (0.54 to 0.75) | 0.55 (0.47 to 0.63) | 0.67 (0.54 to 0.79) | 0.61 (0.42 to 0.80) | 0.43 (0.22 to 0.63) | 0.54 (0.48 to 0.60) | 0.57 (0.43 to 0.70) |
| PCA, RBF SVM | 0.67 (0.56 to 0.77) | 0.55 (0.47 to 0.63) | 0.67 (0.54 to 0.79) | 0.63 (0.45 to 0.82) | 0.44 (0.23 to 0.65) | 0.56 (0.50 to 0.62) | 0.58 (0.44 to 0.70) |
| PCA, gradient boosting | 0.63 (0.53 to 0.73) | 0.54 (0.46 to 0.62) | 0.69 (0.57 to 0.82) | 0.60 (0.41 to 0.79) | 0.47 (0.26 to 0.67) | 0.54 (0.48 to 0.60) | 0.58 (0.43 to 0.71) |
| Voxels, linear SVM | 0.58 (0.46 to 0.70) | 0.53 (0.45 to 0.61) | 0.51 (0.36 to 0.65) | 0.45 (0.25 to 0.65) | 0.44 (0.23 to 0.65) | 0.48 (0.42 to 0.54) | 0.51 (0.45 to 0.57) |
| Voxels, RBF SVM | 0.52 (0.41 to 0.64) | 0.54 (0.46 to 0.63) | 0.58 (0.45 to 0.72) | 0.48 (0.28 to 0.68) | 0.44 (0.23 to 0.66) | 0.52 (0.47 to 0.58) | 0.54 (0.47 to 0.61) |
| PLS, RBF SVM | 0.53 (0.41 to 0.66) | 0.51 (0.42 to 0.59) | 0.60 (0.47 to 0.72) | 0.59 (0.40 to 0.78) | 0.43 (0.23 to 0.64) | 0.53 (0.47 to 0.59) | 0.53 (0.44 to 0.62) |

Column headers give N/transitions. External, pooled is the AUC across all 685 external participants, as reported in the main text; External, random effects is the REML summary of the 4 cohort logit AUCs with Hartung-Knapp CIs. Paired differences are reported in eResults. PCA, principal component analysis; PLS, partial least squares; RBF, radial basis function; SVM, support vector machine.

eTable 15. Calibration and additional validation metrics. Original probabilities, apparent same-sample updates and held-out updates are kept separate. Assessment intercepts/slopes describe the evaluated probabilities; they are not automatically the coefficients of the updating transformation. Further sections add threshold metrics at thresholds chosen in hindsight, the PRONIA reference by diagnostic group, the decision-curve summary, and the random-effects synthesis.

| **Pooled external calibration (N=685; 106 transitions)** | | | | | | | | | | | | | | | | | | | | | | | | | | |
| --- | --- | --- | --- | --- | --- | --- | --- | --- | --- | --- | --- | --- | --- | --- | --- | --- | --- | --- | --- | --- | --- | --- | --- | --- | --- | --- |
| **Probability set** | | | | | **Assessment quantity** | | | | | | | | | | | | **Estimate (95% CI)** | | | | | | | | | |
| Original | | | | | Calibration-in-the-large intercept | | | | | | | | | | | | -1.731 (-2.005 to -1.483) | | | | | | | | | |
| Original | | | | | Joint intercept | | | | | | | | | | | | -1.641 (-1.874 to -1.448) | | | | | | | | | |
| Original | | | | | Calibration slope | | | | | | | | | | | | 0.166 (0.008 to 0.335) | | | | | | | | | |
| Original | | | | | Brier score | | | | | | | | | | | | 0.245 (0.228 to 0.262) | | | | | | | | | |
| Apparent intercept-only | | | | | Calibration-in-the-large intercept | | | | | | | | | | | | 0.000 (0.000 to 0.000) | | | | | | | | | |
| Apparent intercept-only | | | | | Joint intercept | | | | | | | | | | | | -1.355 (-1.770 to -1.000) | | | | | | | | | |
| Apparent intercept-only | | | | | Calibration slope | | | | | | | | | | | | 0.166 (0.001 to 0.337) | | | | | | | | | |
| Apparent intercept-only | | | | | Brier score | | | | | | | | | | | | 0.145 (0.123 to 0.169) | | | | | | | | | |
| Apparent intercept+slope | | | | | Calibration-in-the-large intercept | | | | | | | | | | | | 0.000 (0.000 to 0.000) | | | | | | | | | |
| Apparent intercept+slope | | | | | Joint intercept | | | | | | | | | | | | 0.000 (0.000 to 0.000) | | | | | | | | | |
| Apparent intercept+slope | | | | | Calibration slope | | | | | | | | | | | | 1.000 (1.000 to 1.000) | | | | | | | | | |
| Apparent intercept+slope | | | | | Brier score | | | | | | | | | | | | 0.130 (0.111 to 0.149) | | | | | | | | | |
| **PRONIA original-probability assessment (N=329; 26 transitions)** | | | | | | | | | | | | | | | | | | | | | | | | | | |
| **Quantity** | | | | | | | | | | | | | **Estimate (95% CI)** | | | | | | | | | | | | | |
| Calibration-in-the-large intercept | | | | | | | | | | | | | -2.533 (-3.066 to -2.145) | | | | | | | | | | | | | |
| Joint intercept | | | | | | | | | | | | | -2.435 (-2.946 to -2.069) | | | | | | | | | | | | | |
| Calibration slope | | | | | | | | | | | | | 0.739 (0.407 to 1.087) | | | | | | | | | | | | | |
| Brier score | | | | | | | | | | | | | 0.215 (0.196 to 0.235) | | | | | | | | | | | | | |
| **Cohort-specific and held-out calibration** | | | | | | | | | | | | | | | | | | | | | | | | | | |
| **Cohort / evaluation** | | | | **N/events** | | | | **Offset intercept** | | | **Slope** | | | | **Brier (95% CI)** | | | | | | | | | | **Evaluation reference** | |
| FePsy / Original | | | | 37/16 | | | | 0.008 | | | 0.465 | | | | 0.225 (0.158 to 0.301) | | | | | | | | | | 0.245 | |
| FePsy / Apparent cohort update | | | | 37/16 | | | | 0.000 | | | 1.000 | | | | 0.224 (0.182 to 0.270) | | | | | | | | | | 0.245 | |
| FePsy / Held-out update | | | | 37/16 | | | | 1.559 | | | 3.400 | | | | 0.326 (0.209 to 0.434) | | | | | | | | | | 0.245 | |
| MUC-FRUE / Original | | | | 33/16 | | | | 0.515 | | | -0.030 | | | | 0.325 (0.241 to 0.416) | | | | | | | | | | 0.250 | |
| MUC-FRUE / Apparent cohort update | | | | 33/16 | | | | 0.000 | | | 1.000 | | | | 0.250 (0.243 to 0.256) | | | | | | | | | | 0.250 | |
| MUC-FRUE / Held-out update | | | | 33/16 | | | | 1.817 | | | -0.147 | | | | 0.374 (0.244 to 0.498) | | | | | | | | | | 0.250 | |
| NAPLS-3 / Original | | | | 458/51 | | | | -2.174 | | | 0.078 | | | | 0.247 (0.226 to 0.268) | | | | | | | | | | 0.099 | |
| NAPLS-3 / Apparent cohort update | | | | 458/51 | | | | 0.000 | | | 1.000 | | | | 0.099 (0.077 to 0.126) | | | | | | | | | | 0.099 | |
| NAPLS-3 / Held-out update | | | | 458/51 | | | | -0.958 | | | 0.281 | | | | 0.119 (0.104 to 0.137) | | | | | | | | | | 0.099 | |
| ZInEP / Original | | | | 157/23 | | | | -1.825 | | | 0.424 | | | | 0.227 (0.195 to 0.265) | | | | | | | | | | 0.125 | |
| ZInEP / Apparent cohort update | | | | 157/23 | | | | 0.000 | | | 1.000 | | | | 0.121 (0.087 to 0.158) | | | | | | | | | | 0.125 | |
| ZInEP / Held-out update | | | | 157/23 | | | | -0.082 | | | 4.488 | | | | 0.123 (0.090 to 0.161) | | | | | | | | | | 0.125 | |
| *Training-cohort constant comparators for held-out updating* | | | | | | | | | | | | | | | | | | | | | | | | | | |
| **Held-out cohort** | | | | | | | **Probability from training cohorts** | | | | | | | | | | | | | **Held-out Brier** | | | | | | |
| FePsy | | | | | | | 0.1389 | | | | | | | | | | | | | 0.3316 | | | | | | |
| MUC-FRUE | | | | | | | 0.1380 | | | | | | | | | | | | | 0.3700 | | | | | | |
| NAPLS-3 | | | | | | | 0.2423 | | | | | | | | | | | | | 0.1161 | | | | | | |
| ZInEP | | | | | | | 0.1572 | | | | | | | | | | | | | 0.1251 | | | | | | |
| **Threshold metrics and recorded-sex strata** | | | | | | | | | | | | | | | | | | | | | | | | | | |
| **Population** | | **N/events** | | | | **TP/FP/TN/FN** | | | | | | | | **PPV** | | | | **NPV** | | | **LR+** | | | | | **LR-** |
| PRONIA training cohort | | 329/26 | | | | 24/144/159/2 | | | | | | | | 0.143 | | | | 0.988 | | | 1.942 | | | | | 0.147 |
| External pooled | | 685/106 | | | | 60/261/318/46 | | | | | | | | 0.187 | | | | 0.874 | | | 1.256 | | | | | 0.790 |
| FePsy | | 37/16 | | | | 10/8/13/6 | | | | | | | | 0.556 | | | | 0.684 | | | 1.641 | | | | | 0.606 |
| MUC-FRUE | | 33/16 | | | | 6/6/11/10 | | | | | | | | 0.500 | | | | 0.524 | | | 1.063 | | | | | 0.966 |
| NAPLS-3 | | 458/51 | | | | 30/190/217/21 | | | | | | | | 0.136 | | | | 0.912 | | | 1.260 | | | | | 0.772 |
| ZInEP | | 157/23 | | | | 14/57/77/9 | | | | | | | | 0.197 | | | | 0.895 | | | 1.431 | | | | | 0.681 |
| **Recorded sex** | **N/events** | | | | | | **AUC (95% CI)** | | | | | | | | | | | | **BACC** | | | | | **Brier** | | |
| Male | 526/80 | | | | | | 0.602 (0.534 to 0.670) | | | | | | | | | | | | 0.585 | | | | | 0.228 | | |
| Female | 488/52 | | | | | | 0.598 (0.524 to 0.672) | | | | | | | | | | | | 0.602 | | | | | 0.243 | | |
| **Sensitivity, specificity and balanced accuracy at the original threshold and at thresholds chosen in hindsight** | | | | | | | | | | | | | | | | | | | | | | | | | | |
| **Population** | **N/events** | | | **Threshold** | | | **Sensitivity (95% CI)** | | | | | **Specificity (95% CI)** | | | | | | | | **BACC (95% CI)** | | | | | | **Permuted-outcome maximum, 95th percentile (P)** |
| *Original threshold (decision value 0)* | | | | | | | | | | | | | | | | | | | | | | | | | | |
| PRONIA training cohort | 329/26 | | | 0.000 | | | 0.923 (0.749 to 0.991) | | | | | 0.525 (0.467 to 0.582) | | | | | | | | 0.724 (0.664 to 0.776) | | | | | | — |
| PRONIA, CHR-P only | 164/23 | | | 0.000 | | | 0.913 (0.720 to 0.989) | | | | | 0.525 (0.439 to 0.609) | | | | | | | | 0.719 (0.639 to 0.784) | | | | | | — |
| External pooled | 685/106 | | | 0.000 | | | 0.566 (0.466 to 0.662) | | | | | 0.549 (0.508 to 0.590) | | | | | | | | 0.558 (0.507 to 0.611) | | | | | | — |
| FePsy | 37/16 | | | 0.000 | | | 0.625 (0.354 to 0.848) | | | | | 0.619 (0.384 to 0.819) | | | | | | | | 0.622 (0.457 to 0.772) | | | | | | — |
| MUC-FRUE | 33/16 | | | 0.000 | | | 0.375 (0.152 to 0.646) | | | | | 0.647 (0.383 to 0.858) | | | | | | | | 0.511 (0.333 to 0.665) | | | | | | — |
| NAPLS-3 | 458/51 | | | 0.000 | | | 0.588 (0.442 to 0.724) | | | | | 0.533 (0.483 to 0.582) | | | | | | | | 0.561 (0.488 to 0.631) | | | | | | — |
| ZInEP | 157/23 | | | 0.000 | | | 0.609 (0.385 to 0.803) | | | | | 0.575 (0.486 to 0.660) | | | | | | | | 0.592 (0.478 to 0.694) | | | | | | — |
| *Threshold maximizing BACC in each cohort, chosen in hindsight* | | | | | | | | | | | | | | | | | | | | | | | | | | |
| PRONIA training cohort | 329/26 | | | 0.027 | | | 0.923 (0.749 to 0.991) | | | | | 0.545 (0.487 to 0.602) | | | | | | | | 0.734 (0.668 to 0.784) | | | | | | 0.621 (<.001) |
| PRONIA, CHR-P only | 164/23 | | | 0.061 | | | 0.870 (0.664 to 0.972) | | | | | 0.596 (0.510 to 0.677) | | | | | | | | 0.733 (0.647 to 0.804) | | | | | | 0.632 (<.001) |
| External pooled | 685/106 | | | -0.059 | | | 0.632 (0.533 to 0.724) | | | | | 0.499 (0.458 to 0.541) | | | | | | | | 0.566 (0.513 to 0.617) | | | | | | 0.558 (.02) |
| FePsy | 37/16 | | | -0.037 | | | 0.750 (0.476 to 0.927) | | | | | 0.619 (0.384 to 0.819) | | | | | | | | 0.685 (0.527 to 0.826) | | | | | | 0.693 (.07) |
| MUC-FRUE | 33/16 | | | -0.189 | | | 0.812 (0.544 to 0.960) | | | | | 0.529 (0.278 to 0.770) | | | | | | | | 0.671 (0.518 to 0.822) | | | | | | 0.699 (.10) |
| NAPLS-3 | 458/51 | | | 0.056 | | | 0.569 (0.422 to 0.707) | | | | | 0.577 (0.528 to 0.626) | | | | | | | | 0.573 (0.506 to 0.643) | | | | | | 0.589 (.14) |
| ZInEP | 157/23 | | | 0.254 | | | 0.522 (0.306 to 0.732) | | | | | 0.776 (0.696 to 0.844) | | | | | | | | 0.649 (0.537 to 0.748) | | | | | | 0.632 (.02) |
| **Training-cohort reference by diagnostic group** | | | | | | | | | | | | | | | | | | | | | | | | | | |
| **PRONIA group** | | | **N/transitions** | | | | | | **AUC (95% CI)** | | | | | | | **Balanced accuracy** | | | | | | **Calibration slope (95% CI)** | | | | |
| All | | | 329/26 | | | | | | 0.742 (0.662 to 0.822) | | | | | | | 0.724 | | | | | | 0.74 (0.41 to 1.09) | | | | |
| CHR-P only | | | 164/23 | | | | | | 0.734 (0.639 to 0.829) | | | | | | | 0.719 | | | | | | 0.69 (0.37 to 1.21) | | | | |
| Recent-onset depression | | | 165/3 | | | | | | 0.776 | | | | | | | 0.762 | | | | | | — | | | | |
| **Net benefit relative to the better default strategy** | | | | | | | | | | | | | | | | | | | | | | | | | | |
| **Model** | | | **10%** | | | | | | | **15%** | | | | | | **20%** | | | | | | | **30%** | | | |
| Original probabilities | | | -0.002 (-0.010 to 0.006) | | | | | | | 0.009 (-0.001 to 0.018) | | | | | | -0.034 (-0.047 to -0.021) | | | | | | | -0.114 (-0.133 to -0.095) | | | |
| Held-out recalibration | | | -0.003 (-0.007 to 0.000) | | | | | | | -0.015 (-0.026 to -0.005) | | | | | | -0.052 (-0.063 to -0.041) | | | | | | | -0.034 (-0.047 to -0.022) | | | |
| Clinical only | | | -0.005 (-0.014 to 0.005) | | | | | | | -0.011 (-0.022 to -0.000) | | | | | | -0.027 (-0.040 to -0.013) | | | | | | | -0.025 (-0.040 to -0.011) | | | |
| Clinical + MRI | | | 0.002 (-0.009 to 0.013) | | | | | | | 0.000 (-0.014 to 0.014) | | | | | | -0.016 (-0.031 to -0.001) | | | | | | | -0.019 (-0.036 to -0.003) | | | |
| **Random-effects synthesis of external AUCs** | | | | | | | | | | | | | | | | | | | | | | | | | | |
| **Analysis** | | | | | | | | | | | | **Estimate (95% CI)** | | | | | | | | | | | | | **P** | |
| Random effects, REML (Wald CI) | | | | | | | | | | | | 0.59 (0.50 to 0.67) | | | | | | | | | | | | | — | |
| Random effects, REML (Hartung-Knapp CI) | | | | | | | | | | | | 0.59 (0.46 to 0.70) | | | | | | | | | | | | | — | |
| 95% prediction interval (Hartung-Knapp) | | | | | | | | | | | | 0.38 to 0.76 | | | | | | | | | | | | | — | |
| Never-used cohorts, inverse-variance pooled AUC | | | | | | | | | | | | 0.53 (0.46 to 0.61) | | | | | | | | | | | | | — | |
| Previously used cohorts, inverse-variance pooled AUC | | | | | | | | | | | | 0.66 (0.55 to 0.76) | | | | | | | | | | | | | — | |
| Difference, logit scale (z test) | | | | | | | | | | | | 0.55 (-0.01 to 1.11) | | | | | | | | | | | | | .06 | |
| Difference, logit scale (meta-regression, Hartung-Knapp) | | | | | | | | | | | | 0.55 (0.18 to 0.92) | | | | | | | | | | | | | .02 | |

Intervals use 2000 participant-bootstrap replicates. Apparent updates were refitted in each replicate. The Brier reference assigning the pooled evaluation prevalence to every participant is 0.1308. Near-zero intercepts and unit slopes after an apparent two-parameter update follow from fitting the same observations and do not establish out-of-sample calibration.

These separate training-cohort intervals use 500 percentile-bootstrap replicates. Evaluation-prevalence Brier reference: 0.0728. The original probability field is evaluated here without refitting its historical calibrator.

Cohort-specific updating was fitted and evaluated in that cohort. Held-out (IECV) updating was learned in the other three external cohorts. These separately reported 500-replicate percentile intervals are not the 2000-replicate pooled intervals reported above. Updated-probability assessment parameters are shown, not the transformation coefficients. The evaluation-prevalence reference is descriptive.

TP, true positive; FP, false positive; TN, true negative; FN, false negative; PPV, positive predictive value; NPV, negative predictive value; LR+/LR-, positive/negative likelihood ratios. Classifications are the inherited model labels, not a retuned threshold.

Recorded sex follows the harmonized male/female coding. These estimates pool PRONIA and external cohorts and are not an external-only fairness analysis. AUC, area under the receiver operating characteristic curve; BACC, balanced accuracy; CI, confidence interval. Intervals are DeLong; no cross-sex equivalence test or demographic-invariance claim is made.

Sensitivity and specificity have exact binomial (Clopper-Pearson) 95% CIs, and BACC has percentile 95% CIs from 2000 bootstrap replicates resampling within cohort and outcome. Thresholds are on the scale of the MRI score, the support vector machine decision value. At 0, the threshold reproduces the inherited model labels of every participant. In the PRONIA training cohort, the threshold maximizing BACC (0.027) was close to the original threshold. Hindsight thresholds maximized BACC in the cohort in which they were evaluated and are therefore optimistic. Each hindsight maximum is compared with the maximum BACC reached in 5000 permutations of outcome, within cohort for the pooled external cohorts, with add-one P values. BACC, balanced accuracy; CHR-P, clinical high risk for psychosis.

The recent-onset depression group was coded “ROP” in the analysis export and verified as recent-onset depression against the PRONIA database. With the database’s group labels, the CHR-P-only AUC was 0.733. AUC intervals are DeLong; calibration-slope intervals use 500 percentile-bootstrap replicates. No interval is given for 3 transitions.

Net benefit minus that of the better of treat all and treat none at each threshold probability (95% CI from 2000 bootstrap replicates resampling within cohort and outcome). Original and held-out recalibrated probabilities were evaluated in the external cohorts (685/106); the clinical regressions in leave-one-cohort-out predictions for PRONIA, NAPLS-3 and ZInEP (658/84). Curves are in eFigure 2.

Cohort AUCs were synthesized on the logit scale with DeLong variances; *I*^2^ = 31%. The meta-regression estimated a between-cohort variance of 0, so its interval is narrower than that of the *z* test; with 4 cohorts, neither is a precise test. The never-used estimate rests mainly on NAPLS-3 (51 of 67 transitions). REML, restricted maximum likelihood.

eTable 16. Added value of the MRI score beyond symptoms. Clinical-only and clinical-plus-MRI regressions were evaluated on identical held-out participants under leave-one-cohort-out evaluation. Standardization and second-stage coefficients were learned in the training cohorts. The MRI classifier was not retrained; these are not independent validations of the entire MRI training procedure. Intervals resample within cohort and outcome; the lower panel reports each held-out cohort.

| **Population** | | | **N/events** | **Metric** | | | **Clinical only** | | | **Clinical + MRI** | | | **Difference (95% CI)** | | |
| --- | --- | --- | --- | --- | --- | --- | --- | --- | --- | --- | --- | --- | --- | --- | --- |
| With functioning | | | 254/38 | AUC | | | 0.552 | | | 0.610 | | | 0.058 (0.008 to 0.105) | | |
|  | | |  | Brier | | | 0.1322 | | | 0.1312 | | | -0.0010 (-0.0088 to 0.0075) | | |
| Without functioning | | | 658/84 | AUC | | | 0.568 | | | 0.582 | | | 0.014 (-0.024 to 0.053) | | |
|  | | |  | Brier | | | 0.1190 | | | 0.1202 | | | 0.0013 (-0.0035 to 0.0060) | | |
| **Pooled held-out prediction metrics** | | | | | | | | | | | | | | | |
| **Variant** | **Model** | | | | **N** | | | **Events** | | | | **AUC** | | **Brier** | |
| With functioning | Clinical | | | | 254 | | | 38 | | | | 0.552 | | 0.132 | |
| With functioning | Clinical + MRI | | | | 254 | | | 38 | | | | 0.610 | | 0.131 | |
| Without functioning | Clinical | | | | 658 | | | 84 | | | | 0.568 | | 0.119 | |
| Without functioning | Clinical + MRI | | | | 658 | | | 84 | | | | 0.582 | | 0.120 | |
| **Per held-out cohort** | | | | | | | | | | | | | | | |
| **Variant** | | **Held-out cohort (N/transitions)** | | | | **Clinical AUC** | | | **Clinical + MRI AUC** | | **Difference (95% CI)** | | | | **Paired DeLong P** |
| Symptoms, age, sex | | NAPLS-3 (438/50) | | | | 0.631 | | | 0.598 | | -0.033 (-0.104 to 0.038) | | | | .36 |
|  | | ZInEP (58/11) | | | | 0.580 | | | 0.619 | | 0.039 (-0.048 to 0.130) | | | | .40 |
|  | | PRONIA (162/23) | | | | 0.622 | | | 0.674 | | 0.052 (0.026 to 0.082) | | | | <.001 |
|  | | Pooled (658/84) | | | | 0.568 | | | 0.582 | | 0.014 (-0.024 to 0.053) | | | | — |
| Plus functioning | | NAPLS-3 (37/4) | | | | 0.629 | | | 0.742 | | 0.114 (-0.114 to 0.348) | | | | .37 |
|  | | ZInEP (58/11) | | | | 0.542 | | | 0.582 | | 0.041 (-0.095 to 0.172) | | | | .55 |
|  | | PRONIA (159/23) | | | | 0.581 | | | 0.643 | | 0.062 (0.013 to 0.114) | | | | .02 |
|  | | Pooled (254/38) | | | | 0.552 | | | 0.610 | | 0.058 (0.008 to 0.105) | | | | — |

Difference is clinical-plus-MRI minus clinical-only. Intervals use 2000 paired bootstrap resamples of held-out predictions within cohort and outcome, without refitting either model. Positive AUC and negative Brier differences favor adding the score. The comparison is conditional on the fixed MRI outputs and small event counts, especially the functioning-complete set (38 transitions). It does not establish useful absolute-risk prediction or clinical benefit.

Each cohort’s predictions come from regressions trained in the other 2 cohorts. Intervals use 2000 paired bootstrap replicates resampling within cohort and outcome. Only 37 NAPLS-3 participants had complete ratings including functioning (SOFAS; GAF in PRONIA and ZInEP).

eTable 17. Window definitions, global tests and local estimates. Widths and steps are percentages of each observed analysis-set range. The primary support rule is at least seven transitions and seven nontransitions. The final candidate interval includes the observed maximum; it is displayed only if it meets the support rule.

| **Moderator** | | **Units** | | | | | | **Window width** | | | | | | **Step** | | | | | **Supported / candidates** | | |
| --- | --- | --- | --- | --- | --- | --- | --- | --- | --- | --- | --- | --- | --- | --- | --- | --- | --- | --- | --- | --- | --- |
| Age | | years | | | | | | 6.000 | | | | | | 1.500 | | | | | 13/17 | | |
| BrainAGE | | years | | | | | | 15.646 | | | | | | 3.911 | | | | | 11/17 | | |
| COGDIS | | score points | | | | | | 7.680 | | | | | | 1.920 | | | | | 14/17 | | |
| SIPS-P | | score points | | | | | | 4.600 | | | | | | 1.150 | | | | | 14/17 | | |
| SIPS-N | | score points | | | | | | 6.600 | | | | | | 1.650 | | | | | 15/17 | | |
| Functioning | | GAF-equivalent points | | | | | | 17.000 | | | | | | 4.250 | | | | | 10/17 | | |
| **Primary one-dimensional global tests** | | | | | | | | | | | | | | | | | | | | | |
| **Moderator** | **Analysis N/events** | | | | **Represented N/events** | | | | **Windows** | | | | **Dmax** | | | **p** | | | | **BH q** | |
| Age | 1014/132 | | | | 988/128 | | | | 13 | | | | 0.328 | | | 0.045 | | | | 0.073 | |
| BrainAGE | 1014/132 | | | | 1013/132 | | | | 11 | | | | 0.275 | | | 0.034 | | | | 0.073 | |
| COGDIS | 663/84 | | | | 655/81 | | | | 14 | | | | 0.339 | | | 0.048 | | | | 0.073 | |
| SIPS-P | 676/84 | | | | 636/80 | | | | 14 | | | | 0.311 | | | 0.188 | | | | 0.226 | |
| SIPS-N | 670/84 | | | | 668/82 | | | | 15 | | | | 0.320 | | | 0.043 | | | | 0.073 | |
| Functioning | 422/41 | | | | 402/38 | | | | 10 | | | | 0.164 | | | 0.596 | | | | 0.596 | |
| **Primary two-dimensional global tests** | | | | | | | | | | | | | | | | | | | | | |
| **Surface** | | | | **Analysis N/events** | | | **Represented N/events** | | | | | **Cells** | | | **Dmax** | | | **p** | | | **BH q** |
| Age × BrainAGE | | | | 1014/132 | | | 966/122 | | | | | 80 | | | 0.412 | | | 0.325 | | | 0.650 |
| Age × COGDIS | | | | 663/84 | | | 547/63 | | | | | 62 | | | 0.380 | | | 0.487 | | | 0.698 |
| Age × SIPS-N | | | | 670/84 | | | 558/69 | | | | | 73 | | | 0.441 | | | 0.295 | | | 0.650 |
| BrainAGE × COGDIS | | | | 663/84 | | | 570/72 | | | | | 66 | | | 0.341 | | | 0.698 | | | 0.698 |
| BrainAGE × SIPS-N | | | | 670/84 | | | 652/79 | | | | | 76 | | | 0.541 | | | 0.025 | | | 0.148 |
| COGDIS × SIPS-N | | | | 658/84 | | | 524/61 | | | | | 46 | | | 0.361 | | | 0.633 | | | 0.698 |
| **Unrestricted-permutation comparison** | | | | | | | | | | | | | | | | | | | | | |
| **Trajectory / surface** | | | | | | | | | | **Unrestricted p** | | | | | | | **Unrestricted BH q** | | | | |
| Age | | | | | | | | | | 0.035 | | | | | | | 0.056 | | | | |
| BrainAGE | | | | | | | | | | 0.037 | | | | | | | 0.056 | | | | |
| COGDIS | | | | | | | | | | 0.038 | | | | | | | 0.056 | | | | |
| SIPS-P | | | | | | | | | | 0.098 | | | | | | | 0.117 | | | | |
| SIPS-N | | | | | | | | | | 0.036 | | | | | | | 0.056 | | | | |
| Functioning | | | | | | | | | | 0.544 | | | | | | | 0.544 | | | | |
| Age × BrainAGE | | | | | | | | | | 0.293 | | | | | | | 0.585 | | | | |
| Age × COGDIS | | | | | | | | | | 0.436 | | | | | | | 0.654 | | | | |
| Age × SIPS-N | | | | | | | | | | 0.219 | | | | | | | 0.585 | | | | |
| BrainAGE × COGDIS | | | | | | | | | | 0.688 | | | | | | | 0.688 | | | | |
| BrainAGE × SIPS-N | | | | | | | | | | 0.017 | | | | | | | 0.104 | | | | |
| COGDIS × SIPS-N | | | | | | | | | | 0.614 | | | | | | | 0.688 | | | | |
| **Supported local windows: Age** | | | | | | | | | | | | | | | | | | | | | |
| **Interval** | | | **Midpoint** | | | **Participant mean** | | | | | **N/events** | | | | **AUC (95% bootstrap CI)** | | | | | | |
| 12.00 to 18.00 | | | 15.00 | | | 15.45 | | | | | 277/31 | | | | 0.539 (0.441 to 0.640) | | | | | | |
| 13.50 to 19.50 | | | 16.50 | | | 16.84 | | | | | 388/46 | | | | 0.553 (0.467 to 0.633) | | | | | | |
| 15.00 to 21.00 | | | 18.00 | | | 17.80 | | | | | 453/56 | | | | 0.584 (0.512 to 0.655) | | | | | | |
| 16.50 to 22.50 | | | 19.50 | | | 19.60 | | | | | 463/68 | | | | 0.618 (0.549 to 0.686) | | | | | | |
| 18.00 to 24.00 | | | 21.00 | | | 20.59 | | | | | 462/67 | | | | 0.617 (0.545 to 0.687) | | | | | | |
| 19.50 to 25.50 | | | 22.50 | | | 21.90 | | | | | 376/59 | | | | 0.622 (0.551 to 0.694) | | | | | | |
| 21.00 to 27.00 | | | 24.00 | | | 23.03 | | | | | 314/48 | | | | 0.614 (0.521 to 0.698) | | | | | | |
| 22.50 to 28.50 | | | 25.50 | | | 24.99 | | | | | 217/31 | | | | 0.571 (0.463 to 0.675) | | | | | | |
| 24.00 to 30.00 | | | 27.00 | | | 26.53 | | | | | 172/22 | | | | 0.510 (0.378 to 0.637) | | | | | | |
| 25.50 to 31.50 | | | 28.50 | | | 28.01 | | | | | 141/18 | | | | 0.587 (0.444 to 0.735) | | | | | | |
| 27.00 to 33.00 | | | 30.00 | | | 29.46 | | | | | 126/16 | | | | 0.635 (0.480 to 0.786) | | | | | | |
| 28.50 to 34.50 | | | 31.50 | | | 31.03 | | | | | 96/11 | | | | 0.658 (0.461 to 0.841) | | | | | | |
| 30.00 to 36.00 | | | 33.00 | | | 32.38 | | | | | 77/8 | | | | 0.839 (0.676 to 0.962) | | | | | | |
| **Supported local windows: BrainAGE** | | | | | | | | | | | | | | | | | | | | | |
| **Interval** | | | **Midpoint** | | | **Participant mean** | | | | | **N/events** | | | | **AUC (95% bootstrap CI)** | | | | | | |
| -22.78 to -7.14 | | | -14.96 | | | -10.80 | | | | | 153/26 | | | | 0.601 (0.476 to 0.723) | | | | | | |
| -18.87 to -3.23 | | | -11.05 | | | -7.76 | | | | | 305/42 | | | | 0.568 (0.465 to 0.658) | | | | | | |
| -14.96 to 0.69 | | | -7.14 | | | -4.80 | | | | | 505/71 | | | | 0.585 (0.513 to 0.655) | | | | | | |
| -11.05 to 4.60 | | | -3.23 | | | -2.15 | | | | | 664/86 | | | | 0.571 (0.507 to 0.634) | | | | | | |
| -7.14 to 8.51 | | | 0.69 | | | 0.60 | | | | | 723/89 | | | | 0.570 (0.508 to 0.630) | | | | | | |
| -3.23 to 12.42 | | | 4.60 | | | 3.09 | | | | | 641/78 | | | | 0.585 (0.520 to 0.649) | | | | | | |
| 0.69 to 16.33 | | | 8.51 | | | 5.96 | | | | | 464/51 | | | | 0.587 (0.507 to 0.665) | | | | | | |
| 4.60 to 20.24 | | | 12.42 | | | 9.04 | | | | | 281/30 | | | | 0.649 (0.541 to 0.748) | | | | | | |
| 8.51 to 24.15 | | | 16.33 | | | 12.61 | | | | | 129/13 | | | | 0.700 (0.556 to 0.825) | | | | | | |
| 12.42 to 28.07 | | | 20.24 | | | 16.99 | | | | | 62/9 | | | | 0.782 (0.652 to 0.891) | | | | | | |
| 16.33 to 31.98 | | | 24.15 | | | 21.05 | | | | | 27/7 | | | | 0.843 (0.671 to 0.979) | | | | | | |
| **Supported local windows: COGDIS** | | | | | | | | | | | | | | | | | | | | | |
| **Interval** | | | **Midpoint** | | | **Participant mean** | | | | | **N/events** | | | | **AUC (95% bootstrap CI)** | | | | | | |
| 0.00 to 7.68 | | | 3.84 | | | 2.63 | | | | | 279/32 | | | | 0.545 (0.446 to 0.643) | | | | | | |
| 1.92 to 9.60 | | | 5.76 | | | 5.88 | | | | | 212/25 | | | | 0.541 (0.428 to 0.651) | | | | | | |
| 3.84 to 11.52 | | | 7.68 | | | 7.76 | | | | | 215/21 | | | | 0.567 (0.446 to 0.690) | | | | | | |
| 5.76 to 13.44 | | | 9.60 | | | 9.18 | | | | | 226/24 | | | | 0.626 (0.519 to 0.729) | | | | | | |
| 7.68 to 15.36 | | | 11.52 | | | 11.27 | | | | | 205/25 | | | | 0.633 (0.512 to 0.746) | | | | | | |
| 9.60 to 17.28 | | | 13.44 | | | 13.24 | | | | | 183/24 | | | | 0.643 (0.521 to 0.764) | | | | | | |
| 11.52 to 19.20 | | | 15.36 | | | 15.26 | | | | | 161/22 | | | | 0.645 (0.509 to 0.767) | | | | | | |
| 13.44 to 21.12 | | | 17.28 | | | 17.22 | | | | | 150/20 | | | | 0.517 (0.377 to 0.661) | | | | | | |
| 15.36 to 23.04 | | | 19.20 | | | 18.94 | | | | | 122/16 | | | | 0.495 (0.346 to 0.644) | | | | | | |
| 17.28 to 24.96 | | | 21.12 | | | 20.40 | | | | | 93/13 | | | | 0.462 (0.306 to 0.621) | | | | | | |
| 19.20 to 26.88 | | | 23.04 | | | 22.30 | | | | | 75/11 | | | | 0.547 (0.347 to 0.729) | | | | | | |
| 21.12 to 28.80 | | | 24.96 | | | 24.52 | | | | | 53/8 | | | | 0.675 (0.444 to 0.886) | | | | | | |
| 23.04 to 30.72 | | | 26.88 | | | 26.74 | | | | | 47/8 | | | | 0.779 (0.625 to 0.913) | | | | | | |
| 24.96 to 32.64 | | | 28.80 | | | 27.62 | | | | | 40/7 | | | | 0.801 (0.662 to 0.926) | | | | | | |
| **Supported local windows: SIPS-P** | | | | | | | | | | | | | | | | | | | | | |
| **Interval** | | | **Midpoint** | | | **Participant mean** | | | | | **N/events** | | | | **AUC (95% bootstrap CI)** | | | | | | |
| 3.45 to 8.05 | | | 5.75 | | | 6.15 | | | | | 117/10 | | | | 0.785 (0.681 to 0.879) | | | | | | |
| 4.60 to 9.20 | | | 6.90 | | | 7.34 | | | | | 140/16 | | | | 0.653 (0.502 to 0.790) | | | | | | |
| 5.75 to 10.35 | | | 8.05 | | | 8.50 | | | | | 169/20 | | | | 0.601 (0.466 to 0.729) | | | | | | |
| 6.90 to 11.50 | | | 9.20 | | | 9.48 | | | | | 199/23 | | | | 0.588 (0.474 to 0.699) | | | | | | |
| 8.05 to 12.65 | | | 10.35 | | | 10.72 | | | | | 225/25 | | | | 0.534 (0.421 to 0.649) | | | | | | |
| 9.20 to 13.80 | | | 11.50 | | | 11.57 | | | | | 239/24 | | | | 0.597 (0.489 to 0.699) | | | | | | |
| 10.35 to 14.95 | | | 12.65 | | | 12.48 | | | | | 243/27 | | | | 0.583 (0.473 to 0.688) | | | | | | |
| 11.50 to 16.10 | | | 13.80 | | | 13.79 | | | | | 294/34 | | | | 0.588 (0.491 to 0.689) | | | | | | |
| 12.65 to 17.25 | | | 14.95 | | | 14.73 | | | | | 250/34 | | | | 0.628 (0.526 to 0.725) | | | | | | |
| 13.80 to 18.40 | | | 16.10 | | | 15.40 | | | | | 205/30 | | | | 0.586 (0.469 to 0.698) | | | | | | |
| 14.95 to 19.55 | | | 17.25 | | | 16.25 | | | | | 165/30 | | | | 0.588 (0.478 to 0.696) | | | | | | |
| 16.10 to 20.70 | | | 18.40 | | | 17.94 | | | | | 65/19 | | | | 0.513 (0.350 to 0.665) | | | | | | |
| 17.25 to 21.85 | | | 19.55 | | | 19.17 | | | | | 41/10 | | | | 0.474 (0.287 to 0.661) | | | | | | |
| 18.40 to 23.00 (closed) | | | 20.70 | | | 19.91 | | | | | 32/9 | | | | 0.536 (0.338 to 0.734) | | | | | | |
| **Supported local windows: SIPS-N** | | | | | | | | | | | | | | | | | | | | | |
| **Interval** | | | **Midpoint** | | | **Participant mean** | | | | | **N/events** | | | | **AUC (95% bootstrap CI)** | | | | | | |
| 0.00 to 6.60 | | | 3.30 | | | 3.52 | | | | | 158/16 | | | | 0.608 (0.464 to 0.742) | | | | | | |
| 1.65 to 8.25 | | | 4.95 | | | 5.33 | | | | | 196/19 | | | | 0.588 (0.462 to 0.705) | | | | | | |
| 3.30 to 9.90 | | | 6.60 | | | 6.76 | | | | | 189/20 | | | | 0.530 (0.405 to 0.656) | | | | | | |
| 4.95 to 11.55 | | | 8.25 | | | 8.14 | | | | | 233/25 | | | | 0.501 (0.381 to 0.623) | | | | | | |
| 6.60 to 13.20 | | | 9.90 | | | 10.11 | | | | | 254/21 | | | | 0.451 (0.336 to 0.570) | | | | | | |
| 8.25 to 14.85 | | | 11.55 | | | 11.60 | | | | | 230/24 | | | | 0.535 (0.419 to 0.651) | | | | | | |
| 9.90 to 16.50 | | | 13.20 | | | 12.96 | | | | | 252/30 | | | | 0.558 (0.457 to 0.661) | | | | | | |
| 11.55 to 18.15 | | | 14.85 | | | 14.63 | | | | | 234/27 | | | | 0.600 (0.483 to 0.717) | | | | | | |
| 13.20 to 19.80 | | | 16.50 | | | 16.24 | | | | | 182/29 | | | | 0.618 (0.505 to 0.724) | | | | | | |
| 14.85 to 21.45 | | | 18.15 | | | 17.48 | | | | | 164/23 | | | | 0.588 (0.464 to 0.706) | | | | | | |
| 16.50 to 23.10 | | | 19.80 | | | 19.32 | | | | | 126/18 | | | | 0.593 (0.459 to 0.720) | | | | | | |
| 18.15 to 24.75 | | | 21.45 | | | 20.88 | | | | | 84/16 | | | | 0.610 (0.482 to 0.741) | | | | | | |
| 19.80 to 26.40 | | | 23.10 | | | 22.43 | | | | | 67/15 | | | | 0.682 (0.541 to 0.818) | | | | | | |
| 21.45 to 28.05 | | | 24.75 | | | 23.98 | | | | | 47/12 | | | | 0.650 (0.474 to 0.817) | | | | | | |
| 23.10 to 29.70 | | | 26.40 | | | 25.69 | | | | | 26/9 | | | | 0.771 (0.575 to 0.928) | | | | | | |
| **Supported local windows: Functioning** | | | | | | | | | | | | | | | | | | | | | |
| **Interval** | | | **Midpoint** | | | **Participant mean** | | | | | **N/events** | | | | **AUC (95% bootstrap CI)** | | | | | | |
| 26.25 to 43.25 | | | 34.75 | | | 37.79 | | | | | 65/12 | | | | 0.744 (0.569 to 0.879) | | | | | | |
| 30.50 to 47.50 | | | 39.00 | | | 41.71 | | | | | 95/13 | | | | 0.775 (0.640 to 0.895) | | | | | | |
| 34.75 to 51.75 | | | 43.25 | | | 45.27 | | | | | 144/15 | | | | 0.755 (0.629 to 0.864) | | | | | | |
| 39.00 to 56.00 | | | 47.50 | | | 48.31 | | | | | 181/19 | | | | 0.717 (0.613 to 0.816) | | | | | | |
| 43.25 to 60.25 | | | 51.75 | | | 52.74 | | | | | 209/15 | | | | 0.667 (0.548 to 0.772) | | | | | | |
| 47.50 to 64.50 | | | 56.00 | | | 55.26 | | | | | 191/14 | | | | 0.661 (0.530 to 0.789) | | | | | | |
| 51.75 to 68.75 | | | 60.25 | | | 59.32 | | | | | 176/15 | | | | 0.677 (0.554 to 0.793) | | | | | | |
| 56.00 to 73.00 | | | 64.50 | | | 63.54 | | | | | 164/12 | | | | 0.696 (0.549 to 0.824) | | | | | | |
| 60.25 to 77.25 | | | 68.75 | | | 68.17 | | | | | 118/9 | | | | 0.662 (0.490 to 0.810) | | | | | | |
| 64.50 to 81.50 | | | 73.00 | | | 70.48 | | | | | 107/9 | | | | 0.611 (0.427 to 0.785) | | | | | | |

p-values use within-cohort moderator permutations (5000 valid replicates). q-values correct across the six one-dimensional global tests. The represented population is the union of supported windows, not the sum of window counts.

Paired moderator coordinates were permuted together within cohort. BH q-values correct across all six surfaces, not settings of a preselected surface. No primary 1D or 2D global test survives its six-test family.

These secondary tests shuffle moderators without cohort blocks; the six trajectories and six surfaces form separate correction families. They answer a different exchangeability question from the primary within-cohort tests.

Intervals include the lower endpoint and exclude the upper endpoint. Only the final candidate interval is closed at the observed maximum; it is not displayed when it fails the support rule. The displayed x-coordinate in eFigure 9 is the midpoint, not the participant mean. Confidence intervals are pointwise percentiles from 2000 outcome-stratified resamples with 2000 valid statistics. Windows overlap and their participants must not be added across rows.

Intervals are left-closed and right-open except where marked closed. The displayed x-coordinate in eFigure 9 is the midpoint, not the participant mean. Confidence intervals are pointwise percentiles from 2000 outcome-stratified resamples with 2000 valid statistics. Windows overlap and their participants must not be added across rows.

eTable 18. Sensitivity of performance landscapes. These comparisons vary the support, interval settings or analysis population; they do not independently validate a selected subgroup. N/events refer to available observations unless otherwise stated. The last section reports the design-preserving simulation of the window test (eMethods 8).

| **Joint one-dimensional width/step sensitivity** | | | | | | | | | | | | | | | | | | | | | | |
| --- | --- | --- | --- | --- | --- | --- | --- | --- | --- | --- | --- | --- | --- | --- | --- | --- | --- | --- | --- | --- | --- | --- |
| **Moderator** | **Width / step** | | | | **Windows** | | | **Represented N/events** | | | | | **Dmax** | | | | | **p** | | | | **BH q** |
| Age | 15% / 3.75% | | | | 17 | | | 977/127 | | | | | 0.356 | | | | | 0.066 | | | | 0.133 |
| Age | 20% / 5% | | | | 13 | | | 988/128 | | | | | 0.328 | | | | | 0.045 | | | | 0.073 |
| Age | 25% / 6.25% | | | | 11 | | | 1005/131 | | | | | 0.254 | | | | | 0.116 | | | | 0.139 |
| Age | 30% / 7.5% | | | | 9 | | | 1006/131 | | | | | 0.234 | | | | | 0.124 | | | | 0.186 |
| BrainAGE | 15% / 3.75% | | | | 12 | | | 1002/128 | | | | | 0.299 | | | | | 0.067 | | | | 0.133 |
| BrainAGE | 20% / 5% | | | | 11 | | | 1013/132 | | | | | 0.275 | | | | | 0.034 | | | | 0.073 |
| BrainAGE | 25% / 6.25% | | | | 9 | | | 1013/132 | | | | | 0.275 | | | | | 0.015 | | | | 0.092 |
| BrainAGE | 30% / 7.5% | | | | 7 | | | 1013/132 | | | | | 0.223 | | | | | 0.037 | | | | 0.186 |
| COGDIS | 15% / 3.75% | | | | 17 | | | 652/81 | | | | | 0.420 | | | | | 0.018 | | | | 0.108 |
| COGDIS | 20% / 5% | | | | 14 | | | 655/81 | | | | | 0.339 | | | | | 0.048 | | | | 0.073 |
| COGDIS | 25% / 6.25% | | | | 11 | | | 656/82 | | | | | 0.280 | | | | | 0.081 | | | | 0.139 |
| COGDIS | 30% / 7.5% | | | | 11 | | | 663/84 | | | | | 0.246 | | | | | 0.118 | | | | 0.186 |
| SIPS-P | 15% / 3.75% | | | | 17 | | | 591/79 | | | | | 0.269 | | | | | 0.563 | | | | 0.676 |
| SIPS-P | 20% / 5% | | | | 14 | | | 636/80 | | | | | 0.311 | | | | | 0.188 | | | | 0.226 |
| SIPS-P | 25% / 6.25% | | | | 11 | | | 650/80 | | | | | 0.330 | | | | | 0.060 | | | | 0.139 |
| SIPS-P | 30% / 7.5% | | | | 11 | | | 676/84 | | | | | 0.206 | | | | | 0.414 | | | | 0.496 |
| SIPS-N | 15% / 3.75% | | | | 20 | | | 666/81 | | | | | 0.309 | | | | | 0.143 | | | | 0.214 |
| SIPS-N | 20% / 5% | | | | 15 | | | 668/82 | | | | | 0.320 | | | | | 0.043 | | | | 0.073 |
| SIPS-N | 25% / 6.25% | | | | 13 | | | 670/84 | | | | | 0.251 | | | | | 0.102 | | | | 0.139 |
| SIPS-N | 30% / 7.5% | | | | 11 | | | 670/84 | | | | | 0.244 | | | | | 0.076 | | | | 0.186 |
| Functioning | 15% / 3.75% | | | | 11 | | | 379/34 | | | | | 0.162 | | | | | 0.707 | | | | 0.707 |
| Functioning | 20% / 5% | | | | 10 | | | 402/38 | | | | | 0.164 | | | | | 0.596 | | | | 0.596 |
| Functioning | 25% / 6.25% | | | | 9 | | | 412/39 | | | | | 0.171 | | | | | 0.475 | | | | 0.475 |
| Functioning | 30% / 7.5% | | | | 9 | | | 420/41 | | | | | 0.146 | | | | | 0.513 | | | | 0.513 |
| **Selected surface width/step sensitivity** | | | | | | | | | | | | | | | | | | | | | | |
| **Width / step** | | | | | **Dmax** | | | | | | **p** | | | | | | **BH q (4 settings)** | | | | | |
| 15% / 3.75% | | | | | 0.564 | | | | | | 0.025 | | | | | | 0.039 | | | | | |
| 20% / 5% | | | | | 0.541 | | | | | | 0.024 | | | | | | 0.039 | | | | | |
| 25% / 6.25% | | | | | 0.484 | | | | | | 0.029 | | | | | | 0.039 | | | | | |
| 30% / 7.5% | | | | | 0.386 | | | | | | 0.082 | | | | | | 0.082 | | | | | |
| **Minimum-event support in one-dimensional analyses** | | | | | | | | | | | | | | | | | | | | | | |
| **Moderator** | | | | **Min T / NT** | | | **Analysis N/events** | | | | | | | | **Windows** | | | | | | **Dmax** | |
| Age | | | | 7/7 | | | 1014/132 | | | | | | | | 13 | | | | | | 0.328 | |
| Age | | | | 10/10 | | | 1014/132 | | | | | | | | 12 | | | | | | 0.147 | |
| BrainAGE | | | | 7/7 | | | 1014/132 | | | | | | | | 11 | | | | | | 0.275 | |
| BrainAGE | | | | 10/10 | | | 1014/132 | | | | | | | | 9 | | | | | | 0.132 | |
| COGDIS | | | | 7/7 | | | 663/84 | | | | | | | | 14 | | | | | | 0.339 | |
| COGDIS | | | | 10/10 | | | 663/84 | | | | | | | | 11 | | | | | | 0.184 | |
| SIPS-P | | | | 7/7 | | | 676/84 | | | | | | | | 14 | | | | | | 0.311 | |
| SIPS-P | | | | 10/10 | | | 676/84 | | | | | | | | 13 | | | | | | 0.311 | |
| SIPS-N | | | | 7/7 | | | 670/84 | | | | | | | | 15 | | | | | | 0.320 | |
| SIPS-N | | | | 10/10 | | | 670/84 | | | | | | | | 14 | | | | | | 0.231 | |
| Functioning | | | | 7/7 | | | 422/41 | | | | | | | | 10 | | | | | | 0.164 | |
| Functioning | | | | 10/10 | | | 422/41 | | | | | | | | 8 | | | | | | 0.114 | |
| **Selected surface population/support comparisons** | | | | | | | | | | | | | | | | | | | | | | |
| **Population** | | | **Min T / NT** | | | **Analysis N/events** | | | | | | | | | | **Cells** | | | | **Dmax** | | |
| Pooled | | | 7/7 | | | 670/84 | | | | | | | | | | 76 | | | | 0.541 | | |
| pooled | | | 10/10 | | | 670/84 | | | | | | | | | | 57 | | | | 0.401 | | |
| external only | | | 7/7 | | | 507/61 | | | | | | | | | | 62 | | | | 0.535 | | |
| external only | | | 10/10 | | | 507/61 | | | | | | | | | | 33 | | | | 0.307 | | |
| **Population composition sensitivity** | | | | | | | | | | | | | | | | | | | | | | |
| *Pooled PRONIA and external* | | | | | | | | | | | | | | | | | | | | | | |
| **Moderator** | | **Available N/events** | | | | | | | | **Windows** | | **Dmax** | | | | | | | **AUC range** | | | |
| Age | | 1014/132 | | | | | | | | 13 | | 0.328 | | | | | | | 0.510 to 0.839 | | | |
| BrainAGE | | 1014/132 | | | | | | | | 11 | | 0.275 | | | | | | | 0.568 to 0.843 | | | |
| COGDIS | | 663/84 | | | | | | | | 14 | | 0.339 | | | | | | | 0.462 to 0.801 | | | |
| SIPS-P | | 676/84 | | | | | | | | 14 | | 0.311 | | | | | | | 0.474 to 0.785 | | | |
| SIPS-N | | 670/84 | | | | | | | | 15 | | 0.320 | | | | | | | 0.451 to 0.771 | | | |
| Functioning | | 422/41 | | | | | | | | 10 | | 0.164 | | | | | | | 0.611 to 0.775 | | | |
| *External only* | | | | | | | | | | | | | | | | | | | | | | |
| **Moderator** | | **Available N/events** | | | | | | | | **Windows** | | **Dmax** | | | | | | | **AUC range** | | | |
| Age | | 685/106 | | | | | | | | 12 | | 0.245 | | | | | | | 0.449 to 0.694 | | | |
| BrainAGE | | 685/106 | | | | | | | | 9 | | 0.139 | | | | | | | 0.519 to 0.658 | | | |
| COGDIS | | 499/61 | | | | | | | | 13 | | 0.324 | | | | | | | 0.440 to 0.764 | | | |
| SIPS-P | | 511/61 | | | | | | | | 13 | | 0.163 | | | | | | | 0.442 to 0.604 | | | |
| SIPS-N | | 507/61 | | | | | | | | 16 | | 0.409 | | | | | | | 0.382 to 0.791 | | | |
| Functioning | | 97/15 | | | | | | | | 0 | | — | | | | | | | — to — | | | |
| *CHR-P only (excluding PRONIA depression)* | | | | | | | | | | | | | | | | | | | | | | |
| **Moderator** | | **Available N/events** | | | | | | | | **Windows** | | **Dmax** | | | | | | | **AUC range** | | | |
| Age | | 849/129 | | | | | | | | 13 | | 0.334 | | | | | | | 0.488 to 0.821 | | | |
| BrainAGE | | 849/129 | | | | | | | | 10 | | 0.199 | | | | | | | 0.563 to 0.762 | | | |
| COGDIS | | 663/84 | | | | | | | | 14 | | 0.339 | | | | | | | 0.462 to 0.801 | | | |
| SIPS-P | | 675/84 | | | | | | | | 14 | | 0.309 | | | | | | | 0.474 to 0.783 | | | |
| SIPS-N | | 669/84 | | | | | | | | 15 | | 0.319 | | | | | | | 0.452 to 0.771 | | | |
| Functioning | | 258/38 | | | | | | | | 10 | | 0.145 | | | | | | | 0.612 to 0.757 | | | |
| **Population composition sensitivity** | | | | | | | | | | | | | | | | | | | | | | |
| *Never-used external cohorts* | | | | | | | | | | | | | | | | | | | | | | |
| **Moderator** | | **Available N/events** | | | | | | | | **Windows** | | **Dmax** | | | | | | | **AUC range** | | | |
| Age | | 491/67 | | | | | | | | 12 | | 0.111 | | | | | | | 0.471 to 0.581 | | | |
| BrainAGE | | 491/67 | | | | | | | | 8 | | 0.068 | | | | | | | 0.477 to 0.546 | | | |
| COGDIS | | 441/50 | | | | | | | | 13 | | 0.386 | | | | | | | 0.381 to 0.767 | | | |
| SIPS-P | | 453/50 | | | | | | | | 15 | | 0.223 | | | | | | | 0.424 to 0.647 | | | |
| SIPS-N | | 449/50 | | | | | | | | 13 | | 0.312 | | | | | | | 0.399 to 0.711 | | | |
| Functioning | | 39/4 | | | | | | | | 0 | | — | | | | | | | — to — | | | |
| *Omit FePsy* | | | | | | | | | | | | | | | | | | | | | | |
| **Moderator** | | **Available N/events** | | | | | | | | **Windows** | | **Dmax** | | | | | | | **AUC range** | | | |
| Age | | 977/116 | | | | | | | | 12 | | 0.148 | | | | | | | 0.498 to 0.646 | | | |
| BrainAGE | | 977/116 | | | | | | | | 9 | | 0.257 | | | | | | | 0.555 to 0.812 | | | |
| COGDIS | | 663/84 | | | | | | | | 14 | | 0.339 | | | | | | | 0.462 to 0.801 | | | |
| SIPS-P | | 676/84 | | | | | | | | 14 | | 0.311 | | | | | | | 0.474 to 0.785 | | | |
| SIPS-N | | 670/84 | | | | | | | | 15 | | 0.320 | | | | | | | 0.451 to 0.771 | | | |
| Functioning | | 422/41 | | | | | | | | 10 | | 0.164 | | | | | | | 0.611 to 0.775 | | | |
| *Omit MUC-FRUE* | | | | | | | | | | | | | | | | | | | | | | |
| **Moderator** | | **Available N/events** | | | | | | | | **Windows** | | **Dmax** | | | | | | | **AUC range** | | | |
| Age | | 981/116 | | | | | | | | 13 | | 0.308 | | | | | | | 0.525 to 0.833 | | | |
| BrainAGE | | 981/116 | | | | | | | | 11 | | 0.270 | | | | | | | 0.573 to 0.843 | | | |
| COGDIS | | 663/84 | | | | | | | | 14 | | 0.339 | | | | | | | 0.462 to 0.801 | | | |
| SIPS-P | | 676/84 | | | | | | | | 14 | | 0.311 | | | | | | | 0.474 to 0.785 | | | |
| SIPS-N | | 670/84 | | | | | | | | 15 | | 0.320 | | | | | | | 0.451 to 0.771 | | | |
| Functioning | | 422/41 | | | | | | | | 10 | | 0.164 | | | | | | | 0.611 to 0.775 | | | |
| **Population composition sensitivity** | | | | | | | | | | | | | | | | | | | | | | |
| *Omit NAPLS-3* | | | | | | | | | | | | | | | | | | | | | | |
| **Moderator** | | **Available N/events** | | | | | | | | **Windows** | | **Dmax** | | | | | | | **AUC range** | | | |
| Age | | 556/81 | | | | | | | | 12 | | 0.179 | | | | | | | 0.505 to 0.684 | | | |
| BrainAGE | | 556/81 | | | | | | | | 13 | | 0.340 | | | | | | | 0.520 to 0.860 | | | |
| COGDIS | | 222/34 | | | | | | | | 5 | | 0.151 | | | | | | | 0.638 to 0.789 | | | |
| SIPS-P | | 223/34 | | | | | | | | 9 | | 0.154 | | | | | | | 0.659 to 0.813 | | | |
| SIPS-N | | 221/34 | | | | | | | | 9 | | 0.295 | | | | | | | 0.527 to 0.822 | | | |
| Functioning | | 383/37 | | | | | | | | 11 | | 0.122 | | | | | | | 0.663 to 0.785 | | | |
| *Omit PRONIA* | | | | | | | | | | | | | | | | | | | | | | |
| **Moderator** | | **Available N/events** | | | | | | | | **Windows** | | **Dmax** | | | | | | | **AUC range** | | | |
| Age | | 685/106 | | | | | | | | 12 | | 0.245 | | | | | | | 0.449 to 0.694 | | | |
| BrainAGE | | 685/106 | | | | | | | | 9 | | 0.139 | | | | | | | 0.519 to 0.658 | | | |
| COGDIS | | 499/61 | | | | | | | | 13 | | 0.324 | | | | | | | 0.440 to 0.764 | | | |
| SIPS-P | | 511/61 | | | | | | | | 13 | | 0.163 | | | | | | | 0.442 to 0.604 | | | |
| SIPS-N | | 507/61 | | | | | | | | 16 | | 0.409 | | | | | | | 0.382 to 0.791 | | | |
| Functioning | | 97/15 | | | | | | | | 0 | | — | | | | | | | — to — | | | |
| *Omit ZInEP* | | | | | | | | | | | | | | | | | | | | | | |
| **Moderator** | | **Available N/events** | | | | | | | | **Windows** | | **Dmax** | | | | | | | **AUC range** | | | |
| Age | | 857/109 | | | | | | | | 13 | | 0.318 | | | | | | | 0.509 to 0.827 | | | |
| BrainAGE | | 857/109 | | | | | | | | 9 | | 0.146 | | | | | | | 0.556 to 0.702 | | | |
| COGDIS | | 605/73 | | | | | | | | 15 | | 0.361 | | | | | | | 0.437 to 0.799 | | | |
| SIPS-P | | 618/73 | | | | | | | | 14 | | 0.293 | | | | | | | 0.477 to 0.770 | | | |
| SIPS-N | | 612/73 | | | | | | | | 15 | | 0.278 | | | | | | | 0.484 to 0.762 | | | |
| Functioning | | 364/30 | | | | | | | | 8 | | 0.163 | | | | | | | 0.620 to 0.783 | | | |
| **Design-preserving simulation of the window test** | | | | | | | | | | | | | | | | | | | | | | |
| **Moderator (N/transitions)** | | **True local AUC** | | | | | | | **Window test rejects** | | | | | **Interaction test rejects** | | | | | | | **Mean range (95th percentile)** | |
| SIPS-N (670/84) | | Constant 0.60 | | | | | | | 8.9% | | | | | 6.2% | | | | | | | 0.22 (0.34) | |
| SIPS-N | | 0.50 to 0.70 | | | | | | | 17.9% | | | | | 28.0% | | | | | | | 0.25 (0.38) | |
| SIPS-N | | 0.45 to 0.80 | | | | | | | 41.0% | | | | | 70.1% | | | | | | | 0.31 (0.46) | |
| SIPS-N | | Step 0.55 to 0.75 | | | | | | | 53.5% | | | | | 68.0% | | | | | | | 0.33 (0.47) | |
| SIPS-N | | Observed trajectory | | | | | | | 28.3% | | | | | 23.4% | | | | | | | 0.27 (0.43) | |
| Age (1014/132) | | Constant 0.60 | | | | | | | 3.1% | | | | | 3.4% | | | | | | | 0.18 (0.30) | |
| Age | | 0.50 to 0.70 | | | | | | | 4.7% | | | | | 19.4% | | | | | | | 0.20 (0.33) | |
| Age | | 0.45 to 0.80 | | | | | | | 15.4% | | | | | 58.4% | | | | | | | 0.25 (0.39) | |
| Age | | Step 0.55 to 0.75 | | | | | | | 27.4% | | | | | 50.1% | | | | | | | 0.28 (0.41) | |
| Age | | Observed trajectory | | | | | | | 10.4% | | | | | 27.2% | | | | | | | 0.23 (0.37) | |

Each setting has its own six-moderator BH family. Width and step change together; this is not a width-only comparison at a fixed step. Range endpoints follow the same closed-final-interval rule as the primary analysis.

The surface is SIPS-N × BrainAGE. These q-values correct only four settings of this selected surface, not the original selection among six surfaces. The primary six-surface q is 0.148 (eTable 17). The primary-setting p in this sensitivity family uses a separately indexed deterministic permutation stream and can differ slightly from the primary-family p through Monte Carlo variation; the observed Dmax is identical.

T, transition; NT, nontransition. These support comparisons are descriptive and do not have a new multiplicity-adjusted global inference.

The available external population for this clinical surface is 507 participants/61 transitions, not the full external validation population of 685/106. Ranges and supported cell sets change between populations; a similar Dmax does not demonstrate that the same high-performing region is preserved.

Moderator range recalculated in this population. Dashes indicate insufficient supported windows; no global p-value is inferred from these descriptive comparisons.

1000 replicates per scenario. The window test is the within-cohort permutation test of Dmax (199 permutations); the interaction test is the MRI score × moderator term in a logistic regression with cohort effects. Under a constant AUC, rejection rates estimate the type I error; otherwise, power. Range is Dmax, the largest minus the smallest supported local AUC.

eTable 19. Reweighting for participant characteristics. The upper panel reweights PRONIA participants with CHR-P to the measured characteristics of each external cohort within outcome groups, so that the standardized AUC is the discrimination expected in the training cohort under those characteristics. The lower panel, shown in Figure 2B, reweights each external cohort to the measured characteristics of PRONIA participants with CHR-P, so that its AUC is the external discrimination expected under the characteristics of PRONIA. A gap to the PRONIA AUC that persists after reweighting is not explained by the measured covariates (eMethods 10).

| **External cohort** | **Covariates** | **Membership AUC** | **Effective reference N (transitions)** | **Standardized PRONIA AUC (95% CI)** | **Observed external AUC** |
| --- | --- | --- | --- | --- | --- |
| NAPLS-3 | Age, sex | 0.81 | 97 (14.0) | 0.77 (0.67 to 0.86) | 0.53 |
| NAPLS-3 | Age, sex, SIPS-P, SIPS-N, COGDIS | 0.88 | 29 (5.6) | 0.76 (0.53 to 0.91) | 0.53 |
| ZInEP | Age, sex | 0.61 | 145 (17.5) | 0.77 (0.68 to 0.86) | 0.65 |
| ZInEP | Age, sex, SIPS-P, SIPS-N, COGDIS | 0.73 | 89 (16.2) | 0.78 (0.62 to 0.88) | 0.59 |
| FePsy | Age, sex | 0.66 | 159 (20.1) | 0.72 (0.58 to 0.83) | 0.69 |
| MUC-FRUE | Age, sex | 0.71 | 154 (21.1) | 0.74 (0.60 to 0.84) | 0.56 |

Lower panel. External cohorts reweighted to the measured characteristics of PRONIA participants with CHR-P.

| **External cohort** | **Covariates** | **Membership AUC** | **Effective external N (transitions)** | **External AUC reweighted to PRONIA (95% CI)** | **Unweighted external AUC** |
| --- | --- | --- | --- | --- | --- |
| NAPLS-3 | Age, sex | 0.81 | 135 (22.9) | 0.59 (0.44 to 0.73) | 0.53 |
| NAPLS-3 | Age, sex, SIPS-P, SIPS-N, COGDIS | 0.88 | 94 (12.9) | 0.59 (0.32 to 0.77) | 0.53 |
| ZInEP | Age, sex | 0.61 | 130 (15.7) | 0.66 (0.50 to 0.81) | 0.65 |
| ZInEP | Age, sex, SIPS-P, SIPS-N, COGDIS | 0.73 | 21 (4.4) | 0.66 (0.19 to 0.90) | 0.59 |
| FePsy | Age, sex | 0.66 | 34 (13.6) | 0.67 (0.46 to 0.84) | 0.69 |
| MUC-FRUE | Age, sex | 0.71 | 31 (14.5) | 0.56 (0.33 to 0.78) | 0.56 |

Membership AUC, discrimination of the membership regression between the external cohort and PRONIA participants with CHR-P (higher values indicate less overlap). Effective reference N is the Kish effective sample size of the weighted PRONIA reference, overall and among transitions (23 unweighted); the standardized AUC inherits any optimism of the training estimate. In the lower panel, effective external N is the Kish effective sample size of the weighted external cohort, and external participants outside the covariate range of PRONIA receive weights near zero. The symptom-adjusted ZInEP analysis used the 58 participants (11 transitions) with ratings. Age overlap: 102 of 458 NAPLS-3 participants (22.3%) and 23 of 157 ZInEP participants were younger than the youngest PRONIA participant with CHR-P (15.2 years); AUC was 0.45 outside and 0.55 inside the overlapping range in NAPLS-3 (11 and 40 transitions) and 0.43 and 0.67 in ZInEP (2 and 21 transitions). COGDIS, cognitive disturbances; SIPS-P and SIPS-N, SIPS positive and negative scores.

eTable 20. Technical-shift analyses. Score distributions and discrimination were compared between cohorts and, within NAPLS-3, between acquisition sites. Because the pipeline is linear from gray-matter features to score, a site-specific additive feature offset moves all scores at that site by a constant and leaves within-site discrimination unchanged (eMethods 11).

| **Score distributions by cohort** | | | | | | | | | |
| --- | --- | --- | --- | --- | --- | --- | --- | --- | --- |
| **Cohort** | **Nontransition shift vs PRONIA (SMD)** | | **SD ratio** | **Separation (SMD)** | | | **Observed transition rate** | | **Mean predicted probability** |
| PRONIA | 0 | | 1.00 | 0.90 | | | 7.9% | | 0.42 |
| NAPLS-3 | 0.06 | | 1.01 | 0.10 | | | 11.1% | | 0.42 |
| ZInEP | -0.02 | | 1.01 | 0.51 | | | 14.6% | | 0.42 |
| FePsy | -0.17 | | 0.92 | 0.54 | | | 43.2% | | 0.43 |
| MUC-FRUE | 0.00 | | 1.13 | -0.04 | | | 48.5% | | 0.39 |
| **Acquisition sites within NAPLS-3 (9 sites; 458 participants; 51 transitions)** | | | | | | | | | |
| **Analysis** | | | | | **AUC (95% CI)** | | | | |
| NAPLS-3, cohort AUC | | | | | 0.528 (0.458 to 0.597) | | | | |
| NAPLS-3, within-site AUC (same-site pairs) | | | | | 0.537 (0.461 to 0.609) | | | | |
| NAPLS-3, per-site median centering | | | | | 0.520 | | | | |
| NAPLS-3, per-site z-scoring | | | | | 0.528 | | | | |
| PRONIA (7 sites), cohort AUC | | | | | 0.742 | | | | |
| PRONIA (7 sites), within-site AUC | | | | | 0.734 | | | | |
| **Image quality and global gray-matter volume (NAPLS-3)** | | | | | | | | | |
| **Tertile** | **N/transitions** | **AUC (95% CI), CAT12 rating** | | | | **N/transitions** | | **AUC (95% CI), global gray-matter volume** | |
| Lowest third (best quality; smallest volume) | 153/19 | 0.56 (0.42 to 0.70) | | | | 153/20 | | 0.59 (0.45 to 0.72) | |
| Middle third | 152/16 | 0.53 (0.39 to 0.67) | | | | 152/17 | | 0.41 (0.28 to 0.54) | |
| Highest third (worst quality; largest volume) | 153/16 | 0.49 (0.35 to 0.63) | | | | 153/14 | | 0.58 (0.44 to 0.73) | |

SMD, standardized mean difference (pooled SD). Shift and SD ratio compare participants without transition with those in PRONIA; separation is the SMD between participants with and without transition within the cohort. After outcome-blind per-cohort median centering or z-scoring of the score, the pooled external AUC was 0.573 and 0.566, compared with 0.563 for the original score.

Nontransition score offsets ranged from -0.08 to 0.15 SD across NAPLS-3 sites (Kruskal-Wallis *P* = .99), with SD ratios of 0.80 to 1.11. Intervals here use 2000 bootstrap replicates resampling within site and outcome; Table 2 gives the DeLong interval for the NAPLS-3 AUC (0.45 to 0.61).

The CAT12 image-quality rating uses CAT12’s mark scale, on which lower values indicate better quality (1 = excellent); NAPLS-3 ratings ranged from 1.45 to 2.95 (median, 1.97). Score × image-quality interaction *P* = .59; score × global gray-matter volume interaction *P* = .36.

eTable 21. Placement-value regressions. Placement values rank each participant against PRONIA participants of the other outcome group (eMethods 12), and a positive estimate means ranking more as in PRONIA. Estimates are per SD for continuous covariates and for female vs male sex, from linear regressions fitted separately by outcome group. Intervals and P values are from 2000 bootstrap replicates within cohort and outcome, and q values are Benjamini-Hochberg adjusted within each regression. The regression across the external cohorts included cohort indicators, which are not shown or counted. These analyses are exploratory.

| **Sample** | **Participants** | **Covariate** | **n** | **Estimate (95% CI)** | **P** | **Adjusted q** | **Tests in family** |
| --- | --- | --- | --- | --- | --- | --- | --- |
| NAPLS-3 | With transition | Age, per SD | 50 | 0.05 (−0.06 to 0.13) | .38 | .64 | 5 |
| NAPLS-3 | With transition | Female sex | 50 | 0.01 (−0.14 to 0.16) | .86 | .86 | 5 |
| NAPLS-3 | With transition | SIPS positive, per SD | 50 | 0.03 (−0.04 to 0.10) | .38 | .64 | 5 |
| NAPLS-3 | With transition | SIPS negative, per SD | 50 | 0.10 (0.02 to 0.17) | .02 | .08 | 5 |
| NAPLS-3 | With transition | COGDIS, per SD | 50 | 0.02 (−0.05 to 0.10) | .53 | .67 | 5 |
| NAPLS-3 | Without transition | Age, per SD | 388 | 0.01 (−0.02 to 0.04) | .54 | .54 | 5 |
| NAPLS-3 | Without transition | Female sex | 388 | −0.09 (−0.16 to −0.02) | .007 | .04 | 5 |
| NAPLS-3 | Without transition | SIPS positive, per SD | 388 | 0.01 (−0.02 to 0.05) | .54 | .54 | 5 |
| NAPLS-3 | Without transition | SIPS negative, per SD | 388 | 0.02 (−0.02 to 0.05) | .34 | .54 | 5 |
| NAPLS-3 | Without transition | COGDIS, per SD | 388 | 0.01 (−0.02 to 0.05) | .49 | .54 | 5 |
| PRONIA CHR-P | With transition | Age, per SD | 23 | 0.01 (−0.15 to 0.10) | .96 | .96 | 5 |
| PRONIA CHR-P | With transition | Female sex | 23 | 0.07 (−0.11 to 0.25) | .43 | .76 | 5 |
| PRONIA CHR-P | With transition | SIPS positive, per SD | 23 | −0.07 (−0.18 to 0.07) | .28 | .76 | 5 |
| PRONIA CHR-P | With transition | SIPS negative, per SD | 23 | 0.05 (−0.09 to 0.18) | .46 | .76 | 5 |
| PRONIA CHR-P | With transition | COGDIS, per SD | 23 | 0.01 (−0.16 to 0.12) | .80 | .96 | 5 |
| PRONIA CHR-P | Without transition | Age, per SD | 139 | −0.07 (−0.13 to −0.00) | .05 | .12 | 5 |
| PRONIA CHR-P | Without transition | Female sex | 139 | −0.14 (−0.25 to −0.03) | .02 | .08 | 5 |
| PRONIA CHR-P | Without transition | SIPS positive, per SD | 139 | 0.00 (−0.05 to 0.05) | .90 | .90 | 5 |
| PRONIA CHR-P | Without transition | SIPS negative, per SD | 139 | −0.05 (−0.10 to 0.01) | .09 | .15 | 5 |
| PRONIA CHR-P | Without transition | COGDIS, per SD | 139 | 0.01 (−0.05 to 0.06) | .85 | .90 | 5 |
| All external cohorts | With transition | Age, per SD | 106 | 0.07 (0.01 to 0.13) | .03 | .052 | 2 |
| All external cohorts | With transition | Female sex | 106 | 0.02 (−0.08 to 0.12) | .68 | .68 | 2 |
| All external cohorts | Without transition | Age, per SD | 579 | −0.00 (−0.03 to 0.03) | .81 | .81 | 2 |
| All external cohorts | Without transition | Female sex | 579 | −0.07 (−0.12 to −0.01) | .02 | .046 | 2 |
